# Simpler is not always better: Phylodynamic misspecification and deep-learning corrections

**DOI:** 10.64898/2026.05.07.26352661

**Authors:** Ruopeng Xie, Olivier Gascuel, Anna Zhukova

## Abstract

Phylodynamics bridges the gap between epidemiology and pathogen genetic data. The birth-death (BD) model and its extensions serve to infer the average number of secondary infections *R* and the infection duration *d* from time-scaled pathogen phylogenies. Moreover, more complex models add extra parameters, such as the average length of the incubation period, the proportion of superspreaders in the infected population, or the speed-up in detection after contact-tracing. However, these additional parameters come at an important computational cost: While the simplest, BD, model, has a closed-form solution, its extensions do not and require numerical methods for their likelihood computation. This leads to increased computational times and potential numerical errors. Therefore, the BD model remains the favorite researchers’ choice for real dataset analyses, and is often applied even in cases where more complex epidemiological aspects are present. We investigated, using simulations, how model misspecification influences inference of *R* and *d* in the phylo-dynamic framework. We showed that the use of models not accounting for various epidemiological aspects leads to bias. In particular the simplest, BD, estimator tends to underestimate *R* in the presence of super-spreading, incubation or contact-tracing, which might be dangerous from the public health prospective. In contrast, deep-learning-based estimators for complex models, accounting for multiple epidemiological factors, performed well in our simulations both on the data where those factors were present and where they were absent. This motivates the use of complex epidemiologically realistic estimators, whose design has recently become possible thanks to deep learning.

## 1 Introduction

Many pathogens, especially viruses, evolve rapidly and accumulate mutations in their genomes between transmissions. Phylodynamics [5, 35] leverages these mutations to estimate epidemiological parameters. Phylodynamic models bridge the gap between traditional epidemiology and sequence data, and permit estimation of such parameters as the effective reproduction number, *R*_*e*_, from the topology and time-measured branch lengths of pathogen transmission trees. These trees can be approximated by pathogen phylogenies (i.e., genealogies of the pathogen population) [12, 17], time-scaled [31] using the sampling dates of pathogen genomes collected over time. The likelihood of a given transmission tree under phylodynamic model parameters can be represented with differential equations. Model parameter values can then be found by, for instance, maximizing the tree likelihood. The use of the transmission structure empowers the phylodynamic approach with respect to classical epidemiological methods. The latter make inferences from incidence curves and in the presence of imported cases might give appearance of elevated local transmission, while the former can separate local clusters from imported cases using phylogeographic methods [15, 10, 32].

In the era of rapidly growing pathogen genomic sequence data, the phylodynamic approach is especially promising. However its power to provide valuable insights for preventing epidemic spread comes at a computational cost. A closed form solution of the differential equations exists only for the most simple Birth-Death (BD) model with incomplete sampling [24], and its Skyline version BDSKY [27], where parameter values can change in a piecewise-constant manner. The BD model is similar to the SIR (Susceptible-Infectious-Removed [8]) model in classical epidemiology and models pathogen spread in a homogeneous population. However, unlike the SIR model, it only applies to epidemics in the exponential phase, with an unlimited number of susceptible individuals. Under this model, infected individuals *I* can transmit their pathogen further and eventually get removed.

Removal corresponds to the moment when the individual cannot transmit anymore, for example because they healed, became aware of the infection and took action to avoid transmission, or died. For a fraction of infected individuals *ρ* their pathogen is sampled and sequenced at removal (and becomes available for phylodynamic analyses). Note that sampling therefore occurs through time [24]. The BD model permits the estimation of the *effective reproduction number R*_*e*_ (i.e., expected number of secondary infections) and the average *infection duration d* (i.e., the time between the moment the individual got infected and the moment they stopped transmitting further). Note that this definition of *d* does not strictly correspond to the infection duration in the clinical sense (until the pathogen gets undetectable), but rather corresponds to the public health sense (until the pathogen cannot be transmitted further, e.g., because the person decided to self-isolate). The BDSKY model with its piecewise-constant rates further approximates the classical SIR model, as reduced transmission rates at later time-points can model decrease in the susceptible population. More complex versions of the BD model where transmission and removal rates can vary arbitrarily over time can further bridge the gap between the BD and the SIR models. However, not only no closed form solution might exist for their differential equations, but also as highlighted by Louca *et al*. [16] under such BD models for any true epidemiological scenario there exists a myriad of alternative “congruent” scenarios that cannot be distinguished using phylogenetic data alone. Identifiability is restored when the number of hidden transmission events along branches is known, the information which can in some cases be inferred from the pathogen sequences [3]. In the case of (piecewise-)constant BD(SKY) models, giving one of the model parameters as an input also restores identifiability [26, 27]. This is often the sampling fraction, estimated as the proportion of sampled sequences among the total declared cases. Due to the relative simplicity and speed, the (piecewise)-constant version of the BD model has become widely adopted and remains the first choice for many pathogen analyses. The original BD and BDSKY publications [24, 27] have been cited more than a thousand times^1^.

The BD model was extended to incorporate complex epidemiological aspects, for instance accounting for an incubation period, super-spreading events or contact-tracing of potentially infected recent partners [25, 28, 13, 40]. For these models the differential equations need to be solved numerically. Numerical solution is challenging, time consuming and may lead to underflow errors [23, 36]. This results in extensive use of simpler models and ignoring potentially important aspects of real-life epidemics (e.g., incubation). For instance for the phylodynamic analyses of SARS-CoV-2 pandemic in Europe [9], New Zealand [4], and Brazil [22], the implementation of BDSKY model in a Bayesian framework was used instead of the BD Exposed-Infectious (SKY) model, accounting for incubation (which takes 2-5 days [37], i.e., an important time in the total infection duration). In the cases when the epidemiological question is simple (e.g., estimating *R*_*e*_ to assess whether the epidemic is contained (*R <* 1) and adjust the lockdown measures), a simpler model choice is indeed tempting (and for very large datasets until recently has been the only one possible).

To our knowledge, the consequences of choosing a simpler homogeneous phylodynamic model in a complex heterogeneous epidemiological setting has only been investigated for the incubation period [19] and showed that not accounting for it leads to underestimation of *R*_*e*_. Until recently, due to computational limitations, investigating model misspecification essentially meant identifying potential biases in estimates with the simple BD model on complex epidemic data. However, the recent arrival of deep learning (DL) to the phylodynamic domain [36, 30, 14, 20] enabled replacing complex likelihood computation with much simpler transmission tree simulation (for training of a DL estimator). Once trained, DL estimators permit almost instantaneous parameter inference. DL hence enabled parameter estimation under more complex models, and on larger datasets [36]. However, its practical applications have not been extensive so far and require ad-hoc implementations and time- and memory-consuming training phase. For instance, only few studies used DL-based estimators for SARS-CoV-2 analyses under more complex and epidemiologically relevant scenarios (e.g., with super-spreading [39]). This raises additional questions that we investigate in this study: Do we need DL estimators for complex models? In which settings do they perform better than simple ones?

In the rest of this article we describe a simulation study that addresses these questions. We generated transmission trees under a wide epidemiological parameter range, covering such epidemics as Ebola, SARS-CoV-2, flu and HIV. These trees varied in size from 200 to 5 000 tips, covering a variety of sequence dataset sizes typically available for such epidemics. The trees featured different combinations of presence or absence of epidemiological aspects potentially relevant to these epidemics (incubation, super-spreading and contact tracing). We estimated the epidemiological parameters on these trees using a simple BD estimator and more complex ones (based on deep-learning). We then compared the accuracy of different estimators for common parameters (such as the average number of secondary infections *R*_*e*_ and the infection time *d*), and, when applicable, the model-specific ones (such as the fraction of the total infection duration in the incubation, the fraction of the population who are superspreaders, or the probability to contact-trace). We reported the identified biases and discussed the new opportunities that deep learning opens for phylodynamic analyses of complex epidemics. Finally, to show practical use, we applied our new estimators to Hong Kong’s third COVID-19 epidemic wave.

## 2 Materials and Methods

### 2.1 Birth-Death Exposed-Infectious model with Super-Spreading and Contact Tracing (BDEISS-CT)

The BDEISS-CT model (Fig. 1, Table 1) combines the Birth-Death Exposed Infectious (BDEI), Birth-Death with Super-Spreading (BDSS) and Birth-Death with Contact-Tracing (BD-CT) models [25, 13, 40]. It models a heterogeneous host population of superspreaders and regular spreaders, infected with a pathogen that features an incubation period. Moreover, at the moment of removal (e.g., due to becoming aware of the infection and self-isolating), the infected individual might notify their contacts (person who infected or people who were infected by the individual in question). Such notification leads to reduced time untill removal of the contacts, and hence reduced reproduction number *R*_*e*_.

**Table 1:**
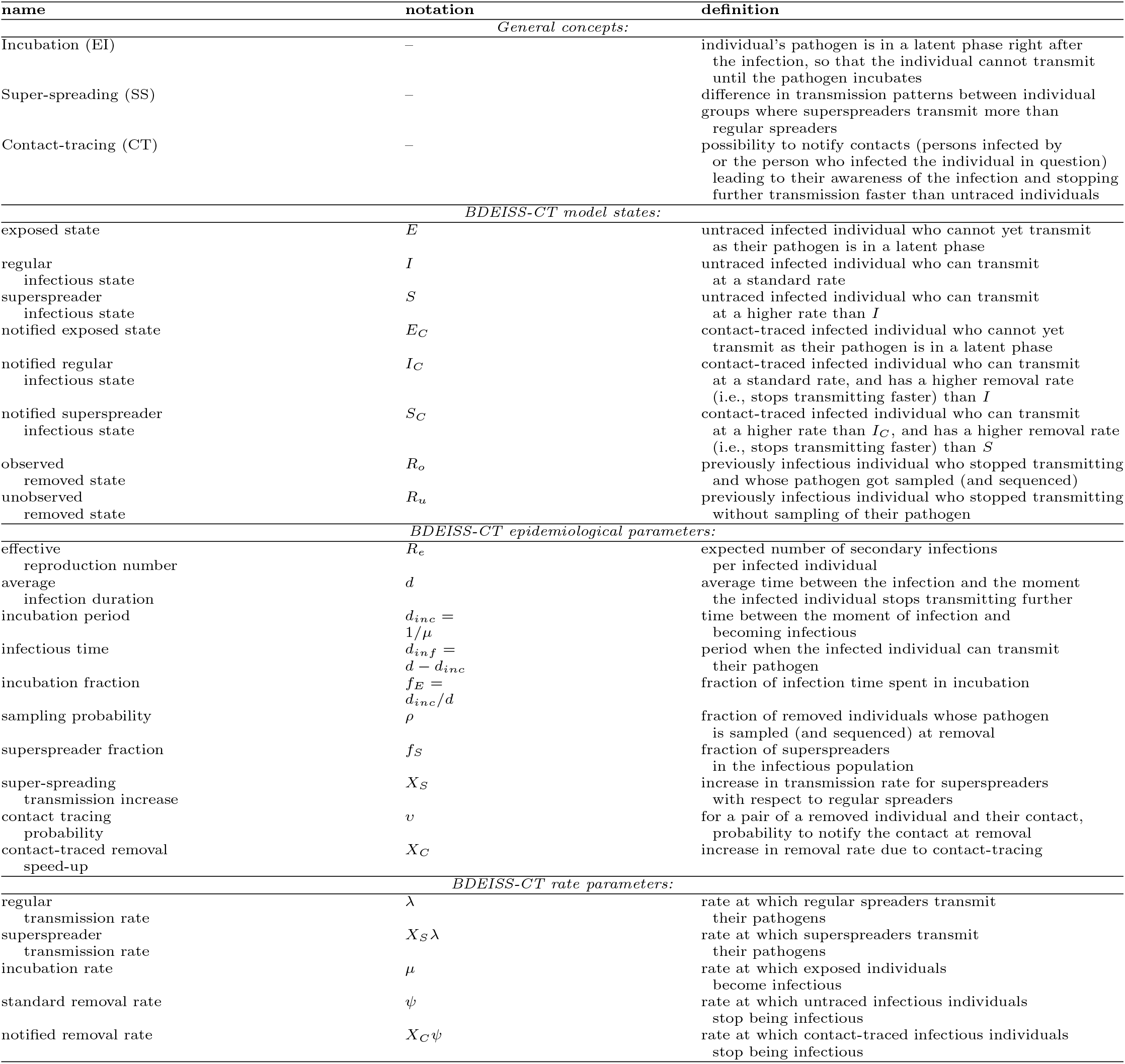
Definitions and notations.

**Figure 1:**
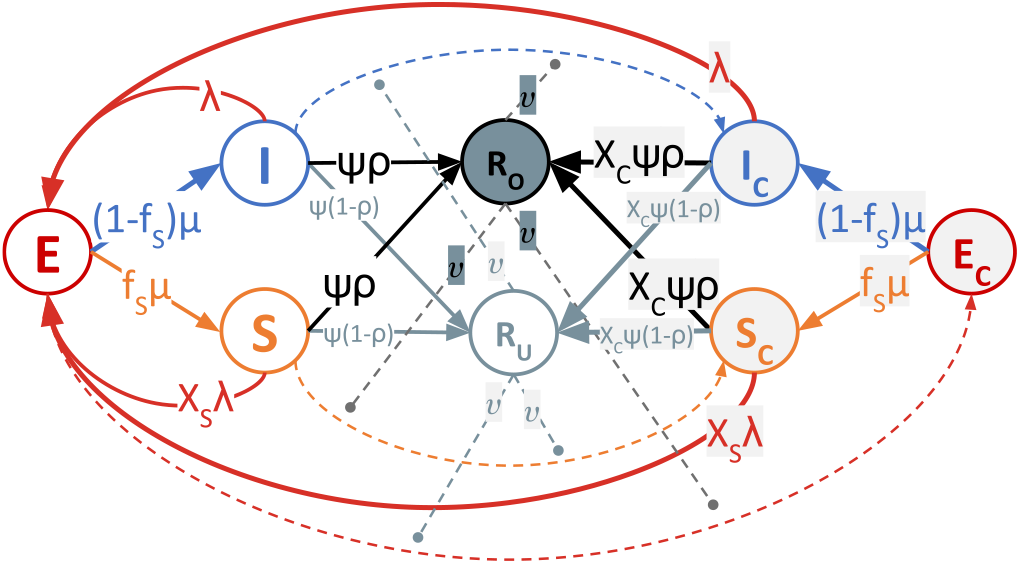
BDEISS-CT model. The model includes three main states, represented as circles – Exposed (E, red), Infectious regular spreader (I, blue) and infectious Super-spreader (S, orange) – as well as their contact-traced versions: *E*_*C*_, *I*_*C*_ and *S*_*C*_ (with gray filling). It also includes two Removed, non-infectious, states – observed (*R*_*o*_, black) and unobserved (*R*_*u*_, gray). Straight solid lines represent state changes, either due to incubation (e.g., the blue line from *E* to *I*), or due to removal (e.g., the black horizontal line from *I* to *R*_*o*_ due to sampling). Curved dashed lines represent state changes due to contact-tracing (e.g., from *I* to *I*_*C*_), they are “catalyzed” (solid dashed lines) by the removal events. Curved solid lines represent transmissions (e.g., the red line for *I* to *E*). The rates at which different events happen are shown on the corresponding lines (apart from contact-tracing, for which the mechanism is more complex, and only the probability to notify is shown).

Mathematically speaking, this model has six possible states: exposed *E* (i.e., infected but not yet infectious), infectious regular spreader *I*, infectious super-spreader *S*, as well as their contact-traced versions: *E*_*C*_, *I*_*C*_, and *S*_*C*_. The regular spreaders (contact-traced or not, *I*_*C*_ or *I*) transmit the epidemic at a constant *transmission rate λ*, while the superspreaders (*S* or *S*_*C*_) transmit it at a higher rate *X*_*S*_*λ*, where *X*_*S*_ *>* 1 represents the *super-spreading transmission increase*. Note, that the model assumes an exponential epidemic phase, with no depletion of susceptible hosts (and hence no decrease in transmission rate over time). The recipient is initially in the exposed state *E*, and their pathogen needs to go through the incubation before being able to be transmitted further. The incubation is modeled via an *incubation rate μ*. The duration of the *incubation period* can then be calculated as *d*_*inc*_ = 1*/μ*. It can also be expressed as a *fraction of incubation* in the total infection time: *f*_*E*_ = *d*_*inc*_*/d*. The average infection duration *d* for this model is a sum of the incubation period *d*_*inc*_ and the *infectious time d*_*inf*_, corresponding to the time interval between the moment the individual got infected and the moment they stop transmitting their pathogen further: *d* = *d*_*inc*_ + *d*_*inf*_. Upon incubation a fraction *f*_*S*_ of individuals becomes superspreaders, while the rest become regular spreaders. If an individual gets contact-traced during the incubation period, their incubation rate does not change (is *μ*), but upon the incubation such an individual becomes either a contact-traced superspreader (*S*_*C*_, with a probability *f*_*S*_) or a contact-traced regular spreader (*I*_*C*_, with a probability 1 − *f*_*S*_). Moreover, infected individuals may also get contact-traced after incubation, in which case their state (*I* or *S*) changes to the corresponding contact-traced one (*I*_*C*_ or *S*_*C*_). The untraced infectious individuals (*I* and *S*) are removed (i.e., stop transmitting) at a *removal rate ψ*, while the contact-traced infectious individuals (*I*_*C*_ and *S*_*C*_) are removed at a higher rate *X*_*C*_*ψ*, where an *X*_*C*_ *>* 1 represents the *contact-traced removal speed-up*. Upon removal the individual’s pathogen may be sampled and sequenced with a *sampling probability ρ*. The exposed individuals (*E* and *E*_*C*_) need to incubate first for their pathogen to be detectable, hence their removal rate is zero. Moreover, upon removal (independently of sampling) an individual (in state *I, S, I*_*C*_ or *S*_*C*_) may notify (all) their contacts with a *contact tracing probability υ* (drawn independently for each contact). Here by contacts of an individual *i* we mean the person who infected *i* and people infected by *i*.

The BDEISS-CT model can be described with the following 8 parameters: {*R, d, ρ*; *f*_*E*_; *f*_*S*_, *X*_*S*_; *υ, X*_*C*_} (see Table 1 for definitions). Setting *f*_*E*_ = 0 removes incubation (*EI*); setting *f*_*S*_ = 0 or *X*_*S*_ = 1 removes super-spreading (*SS*); setting *υ* = 0 or *X*_*C*_ = 1 removes contact tracing (−CT). Fig. 2 shows how the BD, BDEI, BDSS, BDEISS models and their contact-tracing versions (−CT) are nested within each other and the BDEISS-CT model.

**Figure 2:**
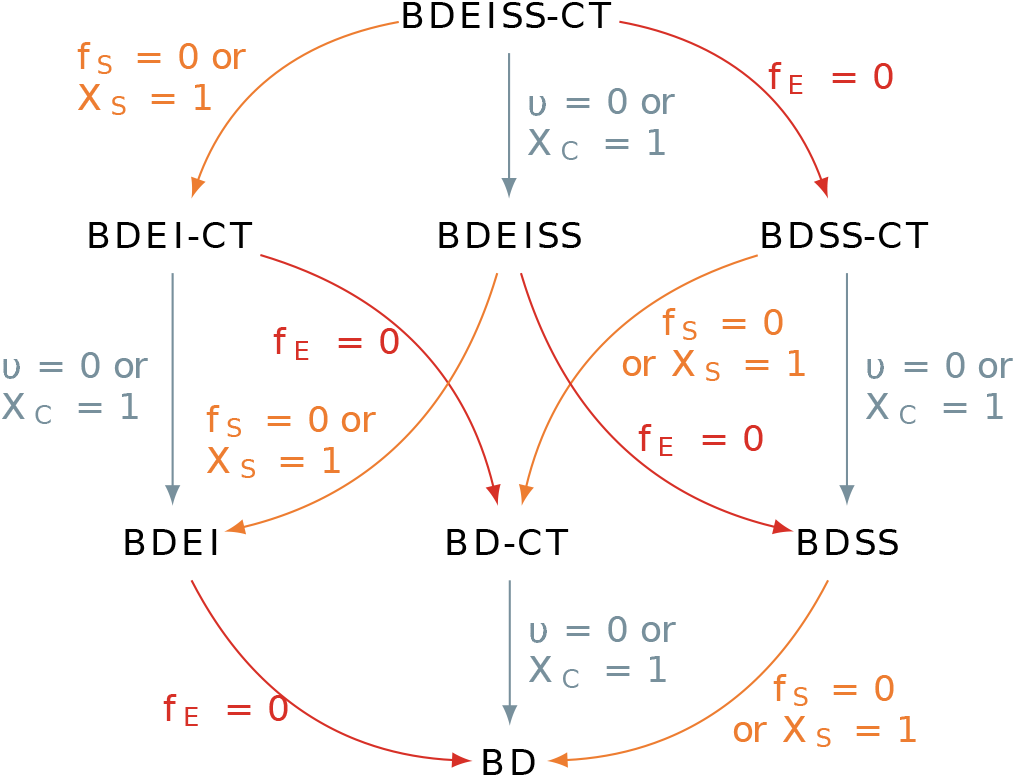
Nesting of BD(EI)(SS)(−CT) models. The arrows link more general models to their nested versions, with the fixed parameter shown along the arrow. The most general, BDEISS-CT, model has 8 parameters: Θ_*BDEISS−CT*_ = {*R, d, ρ, f*_*E*_, *f*_*S*_, *X*_*S*_, *υ, X*_*C*_}, while the most basic, BD, model has only 3 parameters: Θ_*BD*_ = {*R, d, ρ*}.

Note that the BD(EI)(SS) models are compartmental and belong to a broader Multi-Type Birth-Death model family [25, 13]. The epidemiological process under these models is memory-less, where the future events depend only on the current infected individual state, and the transmission tree branches are independent. In the Supplementary Material we explain how *R*_*e*_ and *d* can be calculated from the rate parameters for a general Multi-Type Birth-Death [25, 13] model. In the special case of the BDEISS model, these formulas transform to:

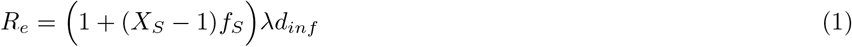

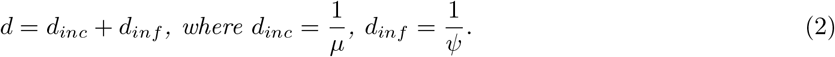

The CT extensions of these models add the dependency between the notifier branch and the contact branches, and keep the memory of who infected whom. This complicates the calculations, and makes the formula for *d*_*inf*_ – and as a consequence the formulas for *d* and *R*_*e*_ – not easily derivable. We know only that the upper bound for *d*_*inf*_ is 1*/ψ* (no CT) and its lower bound is 1*/*(*X*_*C*_*ψ*) (notification systematically during the incubation period). Nevertheless, we can extract the values of these parameters from the transmission tree simulations (see Supplementary Material for details).

#### Simulated datasets

We used treesimulator [40] to generate transmission trees under the BDEISS-CT and its seven nested models shown in Fig. 2. For each model, we generated 1 000 trees for the test set and 2^20^ ≈ 1 000 000 trees for the training+validation set. For each tree the parameter values were drawn uniformly within the bounds listed in Table 2. Note that for model-specific parameters (such as the incubation fraction *f*_*E*_, which is only applicable to models with incubation), the range for the training set was wider than for the test set (e.g., starting at *f*_*E*_ = 0 vs *f*_*E*_ = 0.01). This was done to produce model-specific trees in the test data (e.g., with distinguishable incubation period), while allowing for boundary cases in the training set (e.g., trees without incubation by setting *f*_*E*_ ≈ 0) and accommodating nesting of the models. The number of tips *s*^*\**^ was drawn uniformly between *s*^(0)^ = 2 000 and *S*^(0)^ = 5 000 (inclusive). Additionally, to analyze the impact of tree size on estimations, we generated 3 datasets with smaller tree sizes: [*s*^(1)^ = 1 000, *S*^(1)^ = 2 000], [*s*^(2)^ = 500, *S*^(2)^ = 1 000], [*s*^(3)^ = 200, *S*^(3)^ = 500], where *s*^(*i*)^ and *S*^(*i*)^ correspond to the minimum and maximum size for the trees in the *i*-th dataset. The tree 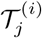 in the dataset *i >* 0 was generated from the corresponding tree 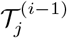 by (1) drawing the target number of tips 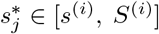, and (2) pruning the tips sampled after the 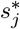-th sampling date from the tree 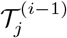.

**Table 2:** Ranges for parameter values for the trees in the training and test data sets. The BD(EI)(SS)(−CT) models in which each parameter is used are listed in the last column (see Fig. 2 for model nesting).

| parameter | training set range | test set range | models |
| --- | --- | --- | --- |
| $R_e$ | [1, 10) | [1, 10) | all |
| $d$ | [1, 31) | [1, 31) | all |
| $\rho$ | [0.05, 0.75) | [0.05, 0.75) | all |
| $f_E$ | [0, 1) | [0.01, 1) | BDEI and its extensions |
| $f_S$ | [0, 0.5) | [0.05, 0.5) | BDSS and its extensions |
| $X_S$ | [1, 25) | [5, 25) | BDSS and its extensions |
| $v$ | [0, 0.75) | [0.05, 0.75) | BD-CT and its extensions |
| $X_C$ | [1, 100) | [10, 100) | BD-CT and its extensions |

We chose wide parameter ranges to cover a variety of epidemics. For instance, *R* ∈ [1, 10) covers epidemics such as HIV-1 (with *R*_*e*_ between 1 and 2 [21, 36]), influenza (with *R*_*e*_ also between 1 and 2 [2]), SARS-CoV-2 (with a potentially higher *R*_*e*_, e.g., between 2 and 4 for its Omicron variant [34]), and Ebola (with estimates between 1 and 8 [18]). The infection time *d* ∈ [1, 31) covers, on a yearly scale, such epidemics as HIV-1 B (whose average time of progression to AIDS in the absence of treatment is 10 years, and the time before treatment start is shorter). On a daily scale, it corresponds to [1 day, 1 month] and covers influenza (with several days of incubation and infectious time of around 1 week), SARS-CoV-2 (2-5 days of incubation and up to 10 days since the symptom onset [37]) and Ebola (2-21 days of incubation plus 1-16 days of infectious time [33]).

#### Parameter estimators

We used our maximum-likelihood parameter estimator, bd [40] for the BD model, to have a reference point for the best achievable performance (as the tree likelihood under the BD model has an analytical formula). This estimator used three different starting points (and kept the result with the highest likelihood), to avoid local maxima. We trained DL-based parameter estimators for all the models shown in Fig. 2, adapting the procedure described in Voznica *et al*. [36]. The estimator for the BD model was trained on a dataset of *N* ≈ 900 000 training BD trees and *n* ≈ 100 000 validation BD trees. For the other models we tried two approaches: the “pure” approach, where, like for BD, the estimator for a model *X* was trained on *N* + *n* (training+validation) trees generated under the same model *X*; and the “mixed” approach, where the estimator for a model *X* was trained on a data set containing trees generated under the model *X* and all its nested models (the same numbers *N* + *n* of total trees equally distributed between different models). For instance, the mixed estimator for the BDEISS model was trained on a dataset of *N/*4+*n/*4 BDEISS, *N/*4+*n/*4 BDEI, *N/*4+*n/*4 BDSS, and *N/*4+*n/*4 BD trees. We tried the mixed approach to put more weight on boundary cases in complex models. For instance *<* 1% of the pure BDEISS-CT training trees were BD-like (with *f*_*E*_, *f*_*S*_ and *υ* close to zero) vs ≥12.5% for the mixed case. As the result, pure estimators specialized less well to their nested models (see for example Table S2). For each model we trained one pure and one mixed DL estimator per simulated tree size range (see “Simulated datasets”). In the following, we report the results for mixed estimators on the largest trees (2 000-5 000 tips), unless otherwise stated.

As in [36], the trees were rescaled so that their average branch length became 1, and the target values of the infection duration *d* were rescaled accordingly. The predicted values of *d* were converted back to the original scale. We encoded the rescaled trees as vectors of 218 summary statistics, most of which were adapted from [36] and some were added to account for peculiarities of the new models, in particular contact-tracing (described in Supplementary Material). These statistics always included the value of the sampling probability *ρ* (needed for model identifiability). Their values were standardized (by removing the mean and scaling to unit variance, independently for each statistic, based on the training dataset for each estimator). The output values were standardized in the same way. Input features (tree summary statistics) were processed through a three-layer fully-connected network (64, 64, and 32 units) incorporating Layer Normalization, ELU activations, and L2 weight regularization. The output was represented as a Dense layer without activation of the size corresponding to the number of parameters pertinent to the estimator’s epidemiological model. For instance for the BDEISS-CT estimator it contained 7×3 neurons: for the three quantiles of *R*_*e*_, *d, f*_*E*_, *f*_*S*_, *X*_*S*_, *υ*, and *X*_*C*_; while for the BD estimator it contained only 2 × 3 neurons: for the three quantiles of *R*_*e*_ and *d*. We used the Adam optimizer with the pinball loss function [29], allowing for estimation of the 0.025^*th*^, 0.5^*th*^ and 0.975^*th*^ quantiles (i.e., the median and a 95%-confidence interval, CI) for each model parameter. To prevent optimization divergence, global L2 gradient norm clipping (clipnorm=0.1) was applied to the Adam optimizer, rescaling any gradient vector whose total L2 norm exceeded 0.1 without altering update directions. We set up an adaptive learning rate schedule that monitored the validation loss and decayed the learning rate by 50% after 5 epochs without improvement, starting at 0.0003 and down to a floor of 10^*−*7^. To enhance early training stability, a custom callback linearly scaled the optimizer’s learning rate from near zero to its target value (0.0003) across the first five epochs. We set the maximal number of epochs to 1 000 (this value was never reached) and used early stopping (with a patience of 15 epochs, the warm-up epochs ignored) to avoid overfitting. The batch size was set to 2^12^ = 8 192.

To enhance model stability against stochastic initialization, for each setting, five independent deep neural network instances were trained under identical architectural configurations, hyperparameter settings and on the same training data sets, differing solely by their initial random seed. A median ensemble estimator was constructed by computing the point-wise median prediction across the five base models for each sample and output quantile. Our evaluations showed that good cross-seed reproducibility was achieved between these estimators and that the median ensemble consistently filtered out single-seed optimization drift, keeping predictions centered within the optimal seed cluster (see the Supplementary Material and Table S2 for details).

#### Real pathogen data

We applied our estimators to the transmission trees reconstructed from real pathogen data, described below. We also checked whether these trees resembled the training data for each model by comparing the values of different summary statistics described in the Supplementary Material on the real data to their distributions on the training data. If they were more than 5 standard deviations away, we considered this to be an incompatibility between the real and training data, and an indication that the estimator’s results should be treated with caution.

#### Hong Kong SARS-CoV-2 wave 3 tree

We applied our estimators to the time-scaled phylogenetic tree of wave 3 of SARS-CoV-2 epidemic in Hong Kong from Xie *et al*. [39], representing its exponential phase from May 13 to August 1 2020. This tree contained 460 samples, corresponding to the sampling proportion *ρ* = 0.238. It was resolved with contact-tracing data and previously analyzed with the BDSS model (see more details on these data in [6, 39]).

#### 2.1.1 Code and data availability

The Snakemake [11] pipelines and source code for training and test dataset generation, tree encoding into summary statistics, DL-based estimator training, and epidemiological parameter inference with DL- and maximum-likelihood-based estimators are available on GitHub at github.com/modpath/bdeissct. The repository also contains the newick files with transmission trees used for tests, as well as the pretrained DL models. The DL-based estimators are also available as a command-line program and a Python 3 package via PyPi (bdext), and via Docker/Singularity (evolbioinfo/bdext).

## 3 Results

The main question that we sought to answer in this study was: If one is only interested in a particular aspect of an epidemic, can one use a simpler model that does not take into account other epidemic aspects? For instance, can one reliably estimate the effective reproduction number *R*_*e*_ with the BD model, even if super-spreading is present in the epidemic? We were also interested in the opposite question: How does a complex estimator perform in a simpler setting? For instance, will a BDSS estimator (which accounts for super-spreading) correctly estimate the absence of super-spreading in a BD epidemic? Do we need to explicitly perform model selection?

To answer these questions we simulated transmission trees under eight epidemic settings representing all the combinations of presence and absence of super-spreading, incubation, and contact-tracing (see “Simulated datasets” for more details). We estimated the epidemiological parameters on these trees with the estimators that ranged in complexity from BDEISS-CT (the most complex model, accounting for all of the above aspects) to BD (the most basic model, which only estimates the reproduction number *R*_*e*_ and the infection duration *d*). Finally we compared the errors and biases for common parameters obtained with different estimators. The results obtained on the 2 000-5 000-tip trees dataset and with estimators trained on a mix of the corresponding and its nested models are shown here: for *R*_*e*_ and *d* in Fig. 3, for the other parameters in Tables 3–5. The results for the smaller tree size datasets and estimators trained on matched models are shown in Tables S3-S16.

**Table 3:**
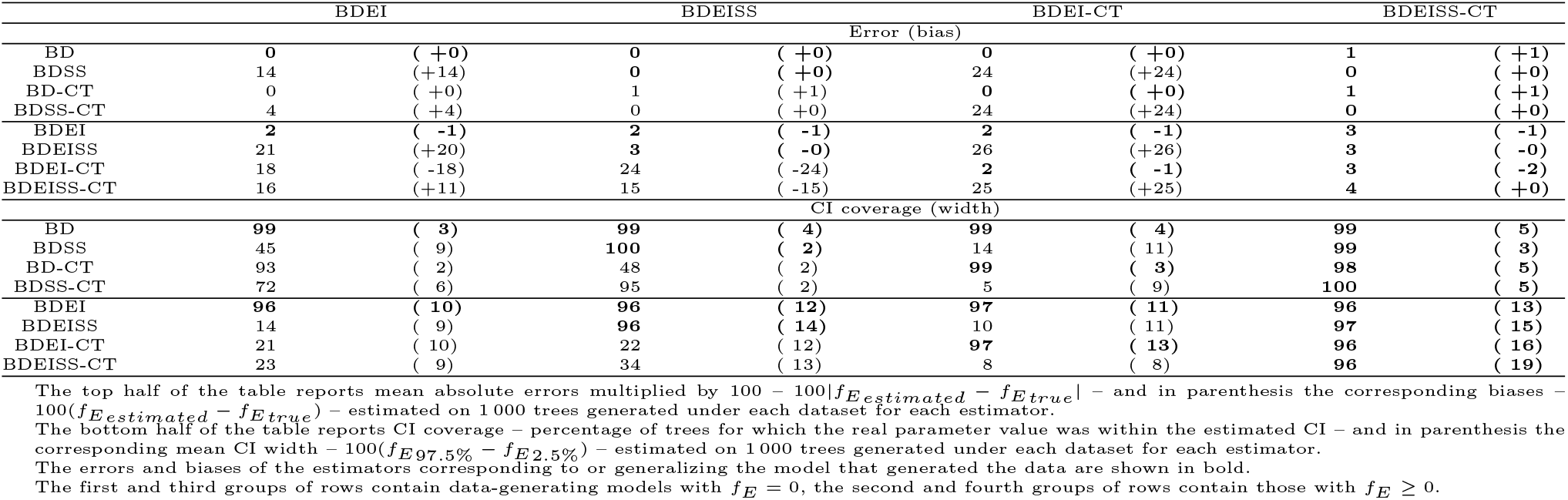
Estimation metrics (errors, biases, CI coverage and width) for the incubation fraction *f*_*E*_ for 2 000-5 000-tip transmission trees generated under different models (rows) and different estimators (columns).

**Figure 3:**
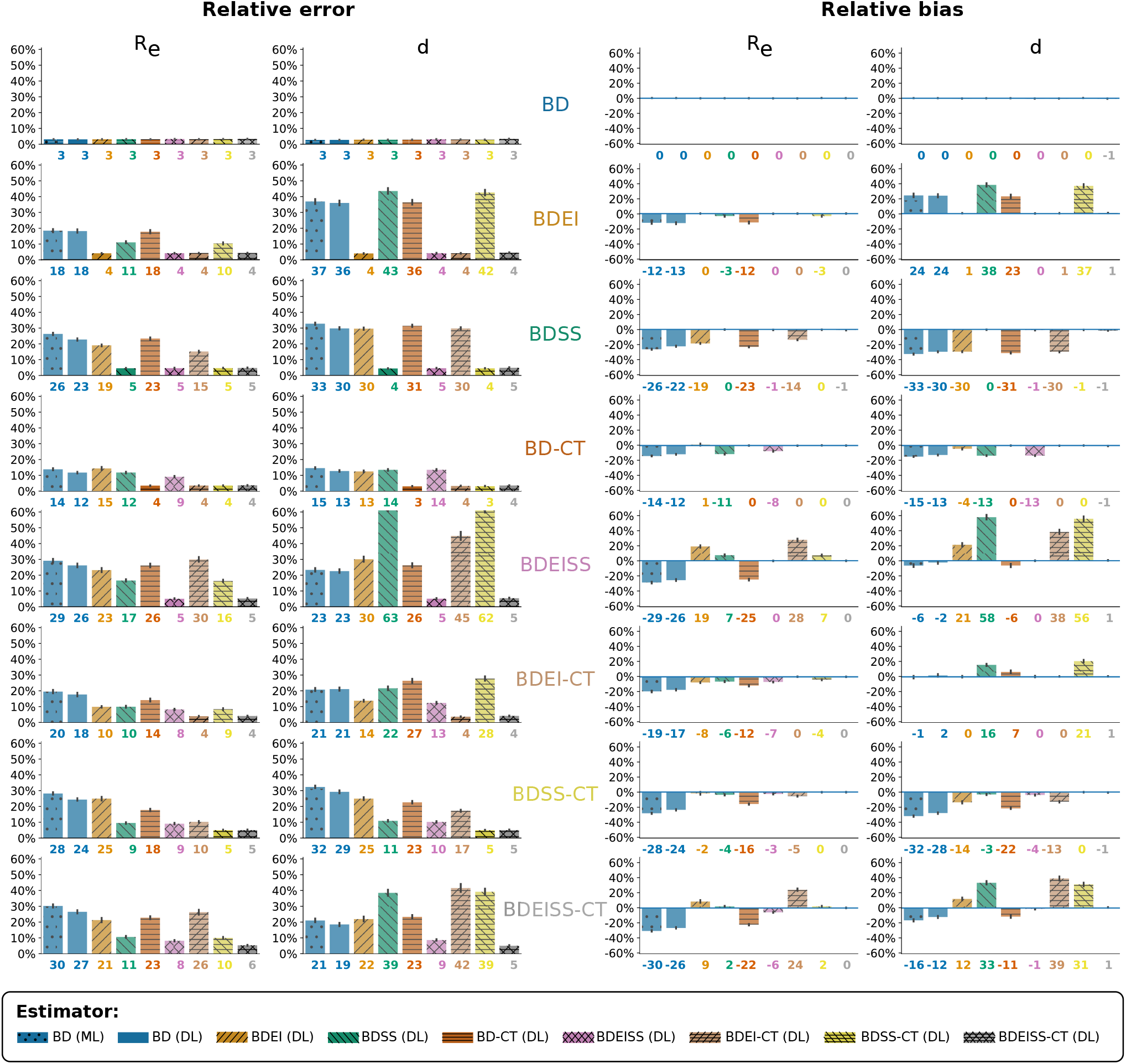
Relative percentage errors (left half) and biases (right half) for effective reproduction number *R*_*e*_ (first and third columns) and average infection time *d* (second and forth columns). The errors and biases were estimated on datasets of 1 000 transmission trees with 2 000-5 000-tips generated under different flavors of the BD(EI)(SS)(−CT) model (corresponding to rows, labels shown in the middle). The bars correspond to different estimators (see the legend at the bottom). (ML) and dotted pattern indicate a maximum-likelihood estimator, while the (DL) indicates deep-learning estimators. The DL estimators were trained on a mix of trees generated under the corresponding model and its nested models (e.g., BDEI and BD trees for the BDEI estimator). Estimators accounting for incubation have forward-slash pattern, those accounting for super-spreading have backward-slash one, and those accounting for contact-tracing have horizontal dashes. Bar heights correspond to the mean relative error/bias on 1 000 trees (also shown below each bar), while the whiskers correspond to 95% confidence intervals. The estimators that do not correspond or generalize the data model (e.g., BDEI estimator for BDSS data) are shown with semi-transparent colors. (In practice, they correspond to large relative error/bias bars, while the matching and opaque ones correspond to low, almost-zero, bars.)

### 3.1 Observed trends

#### DL-based BD estimator performed as well as the maximum-likelihood (ML) one

DL-based BD estimator has errors, biases and CIs compatible with the ML estimator (Figs. 3,4, two left-most blue bars).

**Figure 4:**
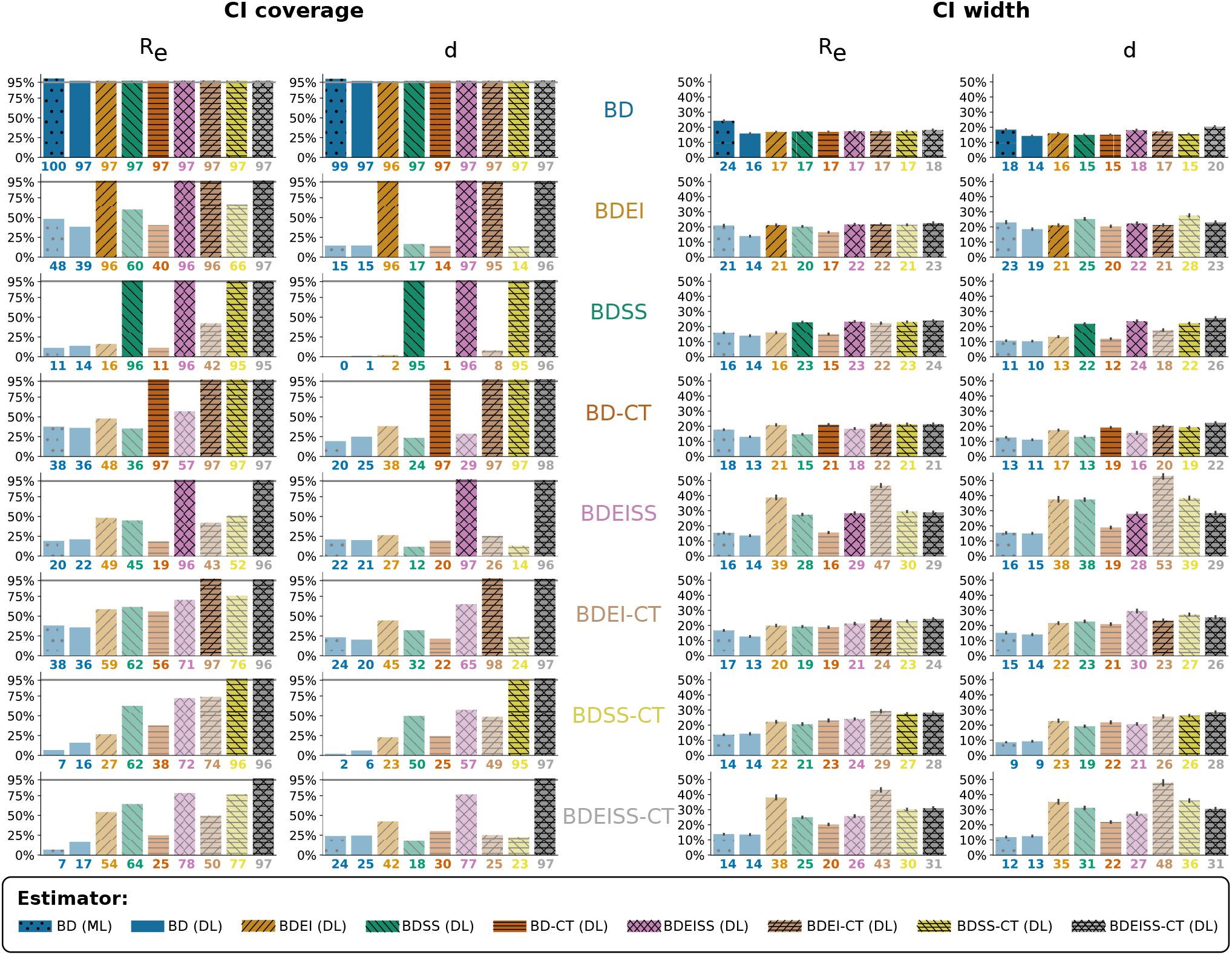
95%-CI coverage (left half, percentage of trees for which the true parameter value is within the estimated interval) and relative width (right half, (*p*_97.5%_ − *p*_2.5%_)*/p*_*true*_ for *p* ∈ {*R, d*}) for effective reproduction number *R*_*e*_ (first and third columns) and average infection time *d* (second and forth columns). The CIs were estimated on datasets of 1 000 transmission trees with 2 000-5 000-tips generated under different flavors of the BD(EI)(SS)(−CT) model (corresponding to rows, labels shown in the middle). The bars correspond to different estimators (see the legend at the bottom). (ML) and dotted pattern indicate a maximum-likelihood estimator, while the (DL) indicates deep-learning estimators. The DL estimators were trained on a mix of trees generated under the corresponding model and its nested models (e.g., BDEI and BD trees for the BDEI estimator). Their CIs were estimated with pinball loss [29]. For the BD (ML) estimator the CIs are approximate: they were obtained from the CIs of the rate parameters (*λ* and *ψ*) as *R*_*e*,97.5%_ = *λ*_97.5%_*/ψ*_2.5%_; *R*_*e*,2.5%_ = *λ*_2.5%_*/ψ*_97.5%_; *d*_97.5%_ = 1*/ψ*_2.5%_; *d*_97.5%_ = 1*/ψ*_2.5%_. The rate parameter CIs were estimated using Wilks’ method [38]. Estimators accounting for incubation have forward-slash pattern, those accounting for super-spreading have backward-slash one, and those accounting for contact-tracing have horizontal dashes. Bar heights correspond to the CI coverage/mean relative width on 1 000 trees (also shown below each bar), while the whiskers for the width correspond to 95% confidence intervals. The estimators that do not correspond or generalize the data model (e.g., BDEI estimator for BDSS data) are shown with semi-transparent colors.

**Using a simple estimator on complex data leads to biases in estimations**, which overall increase with data complexity. For instance the average error for the *R*_*e*_ estimation with the maximum likelihood BD estimator was 3% on the BD data, it increased to 18% on the BDEI data, to 29% on BDEISS data, and finally to 30% on the BDEISS-CT data. The bias also increased, from 0% on the BD data to −12% on the BDEI data, to −29% on the BDEISS data, and finally to −30% on the BDEISS-CT data (Fig. 3, the left-most blue bar). The same trend is observed for the CI coverage: it dropped from 100% on the BD data to only 7% of trees on the BDEISS-CT data (Fig. 4, the left-most blue bar).

**BD estimators tend to underestimate R when complex epidemic aspects are present** (Fig. 3 right panel, two left-most blue bars). This is particularly dangerous from the public health perspective as it suggests that the epidemic is better contained than it really is.

**Accounting for additional epidemic aspects even if they are absent increases CI width but not the error**, as long as the estimator can infer their absence (e.g., output zero for the incubation or super-spreading fractions, *f*_*E*_ and *f*_*S*_, or the CT probability *υ*). For instance, the errors and biases for the estimations of *R*_*e*_ and *d* on the BD data (first rows in Fig. 3 left panel) are within 3% for all the estimators), and the true *R*_*e*_ value is within their CIs for *>* 95% of trees (first rows in Fig. 4 left panel). Tables 3-5 also show that in the absence of incubation, super-spreading, or contact-tracing the estimators for the correct or more general models correctly infer the values of correspondingly *f*_*E*_, *f*_*S*_, or *υ* close to zero, and their CIs include zero in *>* 95% of cases. However, the CIs of more general estimators may be wider than those of model-specific ones (e.g., 12 for BDEISS-CT vs 10 for BDSS estimator, when estimating *f*_*S*_ on the BDSS data, see Table 4). Finally, the most complex estimator, BDEISS-CT, performs well at estimating all the parameters in all the settings (the last bar in Figs. 3,4 and the last column of Tables 3-5).

**Table 4:**
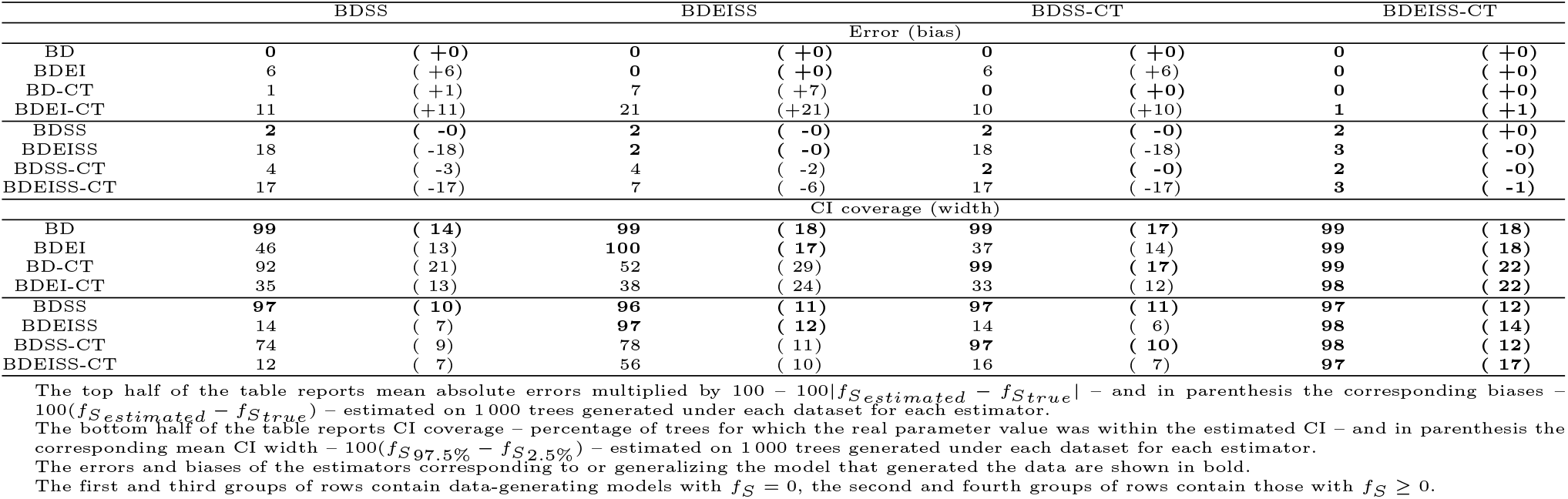
Estimation metrics (errors, biases, CI coverage and width) for the super-spreader fraction *f*_*S*_ for 2 000-5 000-tip transmission trees generated under different models (rows) and different estimators (columns).

#### Accounting for at least some of the epidemic aspects is usually better than not accounting for anything

For instance, the error and bias for *R*_*e*_ on BDSS-CT data were lower for the BDSS estimator and BD-CT estimators (error: 9% and 18%, bias: −4% and −16%) than for the BD ones (24-28% and −24– −28%), while with the correct estimator (BDSS-CT) their values were the lowest (5% and 0%, row BDSS-CT in Fig 3). Moreover, the BDSS and BD-CT CIs included the true *R*_*e*_ value for respectively 62% and 38% of trees, while the BD-estimated CIs only included it in 7-16%. The correct estimator’s CIs (BDSS-CT) included the true *R*_*e*_ value for 96% of trees (row BDSS-CT in Fig 4 left panel).

#### Unaccounted-for epidemic aspects might get confused with something else

In particular, *incubation for super-spreading, and vice versa* (Table 4, rows BDEI and BDEI-CT, columns BDSS and BDSS-CT; Table 3, rows BDSS and BDSS-CT, columns BDEI and BDEI-CT). *Super-spreading and contact-tracing* might be confused one for another as well (Table 5, rows BDSS and BDEISS, columns BD-CT and BDEI-CT; Table 4, rows BD-CT and BDEI-CT, columns BDSS and BDEISS). However, *incubation does not get confused with CT, and vice versa* (Table 5, row BDEI, columns BD-CT and BDSS-CT; Table 3, row BD-CT, columns BDEI and BDEISS).

**Table 5:**
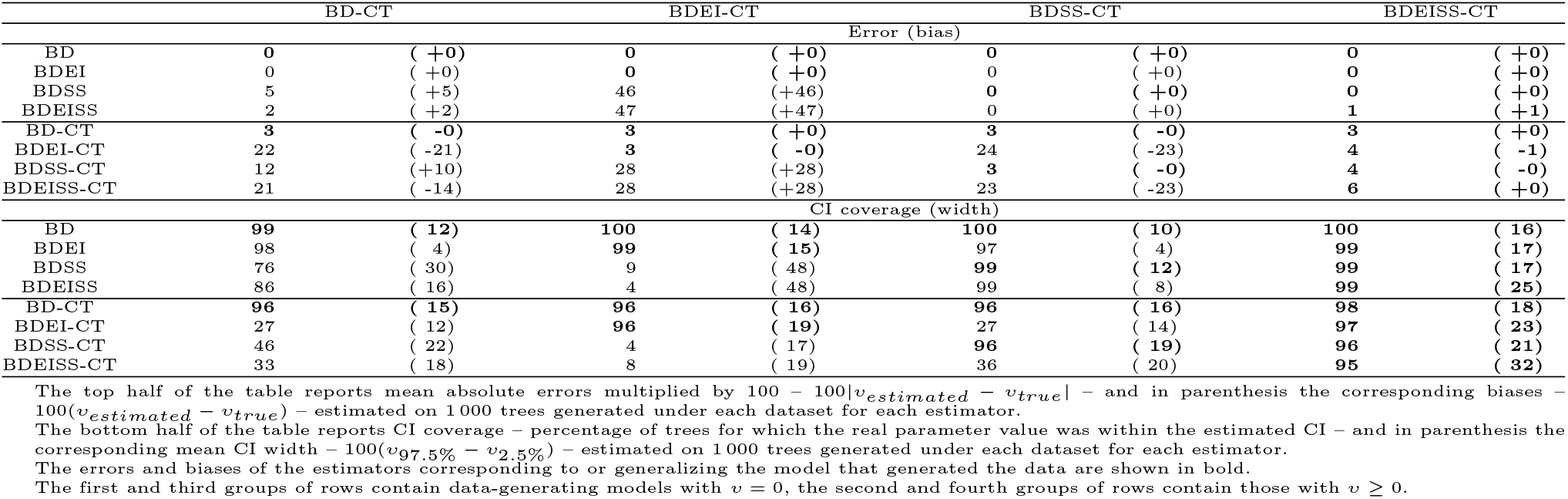
Estimation metrics (errors, biases, CI coverage and width) for the contact-tracing probability *υ* for 2 000-5 000-tip transmission trees generated under different models (rows) and different estimators (columns).

The above confusion trends can also be explained using patterns in the transmission trees. Incubation and super-spreading leave similar internal branch patterns. Incubation leads to length differences between sister branches, as right after the transmission (internal node) the transmitter (one of the child branches) is ready to transmit again or be removed, while the recipient (the other child branch) needs to undergo incubation first. Under super-spreading, when the transmitter and the recipient states are different, the same pattern arises for internal sister branches: the one corresponding to a regular spreader is on average longer. This is due to regular spreaders transmitting less over the same infectious time, hence taking longer between transmissions.(Note, that internal branches represent times between transmissions.) CT, on the contrary, does not change the times between transmissions, hence not leading to such a pattern.

Super-spreading and CT produce similar patterns in tip time differences. Under CT, when an infected individual (the notifier) is removed, their notified contacts are removed soon afterwards. This speeds up the contacts’ removal and therefore shortens the gap between the notifier’s removal time and theirs – a gap that is smaller, on average, than the one between a transmitter and a recipient when no notification takes place. Super-spreading also shortens the removal-time gap between a superspreader transmitter and their first recipients, compared to regular transmitters. But here the mechanism is different: because superspreaders and regular transmitters have the same infectious period, the shorter gap must reflect the timing of transmission itself. Under the BDSS model, superspreaders transmit sooner after infection, which reduces the time between their removal and their recipients’. Incubation, by contrast, has the opposite effect: it lengthens the tip time difference by adding the incubation period (as explained above).

#### Having larger trees reduces uncertainty and increases accuracy with correct models

Indeed, CIs are narrower and errors are lower on larger datasets, when the estimator model matches (or generalizes) the model that generated the data (Tables S3-S16). For instance, the error for *d* with the BDEI(SS)(−CT) estimators on BDEI trees dropped from 9-14% to 4-5%, and the CI width from 49-68% to 19-26%, as the tree sizes increased from 200-500 tips to 2 000-5 000 tips (Tables S4 and S11, row BDEI, columns BDEI, BDEISS, BDEI-CT and BDEISS-CT). However, for mismatched estimators, the errors overall did not decrease, while the CI widths did, e.g., the errors for *d* with the BD-CT estimator on BDEI data stayed around 36% for all tree sizes, though the CI width decreased from 52-55% to 20-25% (Tables S4 and S11, row BDEI, column BD-CT). This is dangerous, as mismatched estimators become more and more certain in their biased predictions.

#### *X*_*C*_ and *X*_*S*_ are the most difficult parameters to estimate

The average errors were more than 10% for all the estimators (Tables S8-S9), and many CIs close or wider than 100% (Tables S15-S16). This is not surprising as there are few traces of these parameters in the tree (i.e., few notified external branches for *X*_*C*_ and few superspreader internal branches for *X*_*S*_). This behavior was also observed in previous studies [36, 39, 20].

#### DL predictors trained on a mix of nested models generalize better

As the BDEISS-CT training data contained *<*1% of BD-like trees (with *f*_*E*_, *f*_*S*_ and *υ* close to zero), it is not surprising that it performed worse on the BD dataset than the BDEISS-CT estimator trained on nested models, with ≥ 12.5% of them containing BD-like data (Tables S3-S16). Strong performance of nested-model-trained estimators suggests that an explicit model selection step is not required. Rather than adopting a two-step approach – in which a model finder first selects the most appropriate model for the input tree, followed by a model-specific estimator to infer the parameters (e.g., as in Voznica *et al*. [36]) – one can instead train a single estimator on data generated from all models simultaneously (provided each model is represented by enough examples). This estimator will then implicitly perform model selection and infer the appropriate parameters directly from the input data (as the mixed BDEISS-CT estimator here).

### Application to SARS-CoV-2 wave 3 epidemic in Hong Kong

For the SARS-CoV-2 tree, BD and BD-CT models were incompatible according to the summary statistics check (see 2.1 for details): median length of internal branches, variance of tip branch lengths and the 25^*th*^ percentile of the sum of branch lengths of 4-chains were more than 5 standard deviations away from the corresponding mean values of the training data. We report the predictions of these estimators for completeness, but the check suggests they should not be trusted. Moreover, as the other estimators pass the check, the mismatch between the BD(−CT) training data and the SARS-CoV-2 tree is most likely due to the presence of complex epidemiological aspects in the latter.

Using the BDEISS-CT estimator for the SARS-CoV-2 wave 3 epidemic in Hong Kong (Fig. 5, Table S16), we obtained the reproduction number *R*_*e*_ = 1.91 (1.5 − 2.8). This point estimate is higher than the ones estimated with all the other models, as well as from a previous study on this data, using the BDSS model and an epidemiological approach from incidence data [39]. This is compatible with the trends obtained on simulated data: our simulations showed that simpler models tend to underestimate *R*_*e*_. We estimated the infection duration *d* = 7 (5 − 11) days, most of which (92 (74 − 95) %) corresponds to incubation. The latter part could be explained by the high CT probability 0.7 (0.4 − 0.8) with a very large (80 (37 − 100) times) increase in removal rate. This means that once notified, the infected person immediately takes care to prevent further transmission (and if notified during the incubation period does not transmit at all). We estimated that there is potentially no super-spreading (*f*_*S*_ = 0 (0 − 0.3)), and even if there is, the difference in transmission rate is not large (*X*_*S*_ = 1 (1 − 7) times). Assuming no super-spreading, the BDEI-CT estimator gives very similar values for all the parameters to BDEISS-CT: *R*_*e*_ = 1.73 (1.5 − 2.6); *d* = 7 (5 − 12) [days] out of which 95 (85 − 98)% is incubation; *υ* = 0.7 (0.5 − 0.8) with 88 (40 − 107) times faster removal after notification.

**Figure 5:**
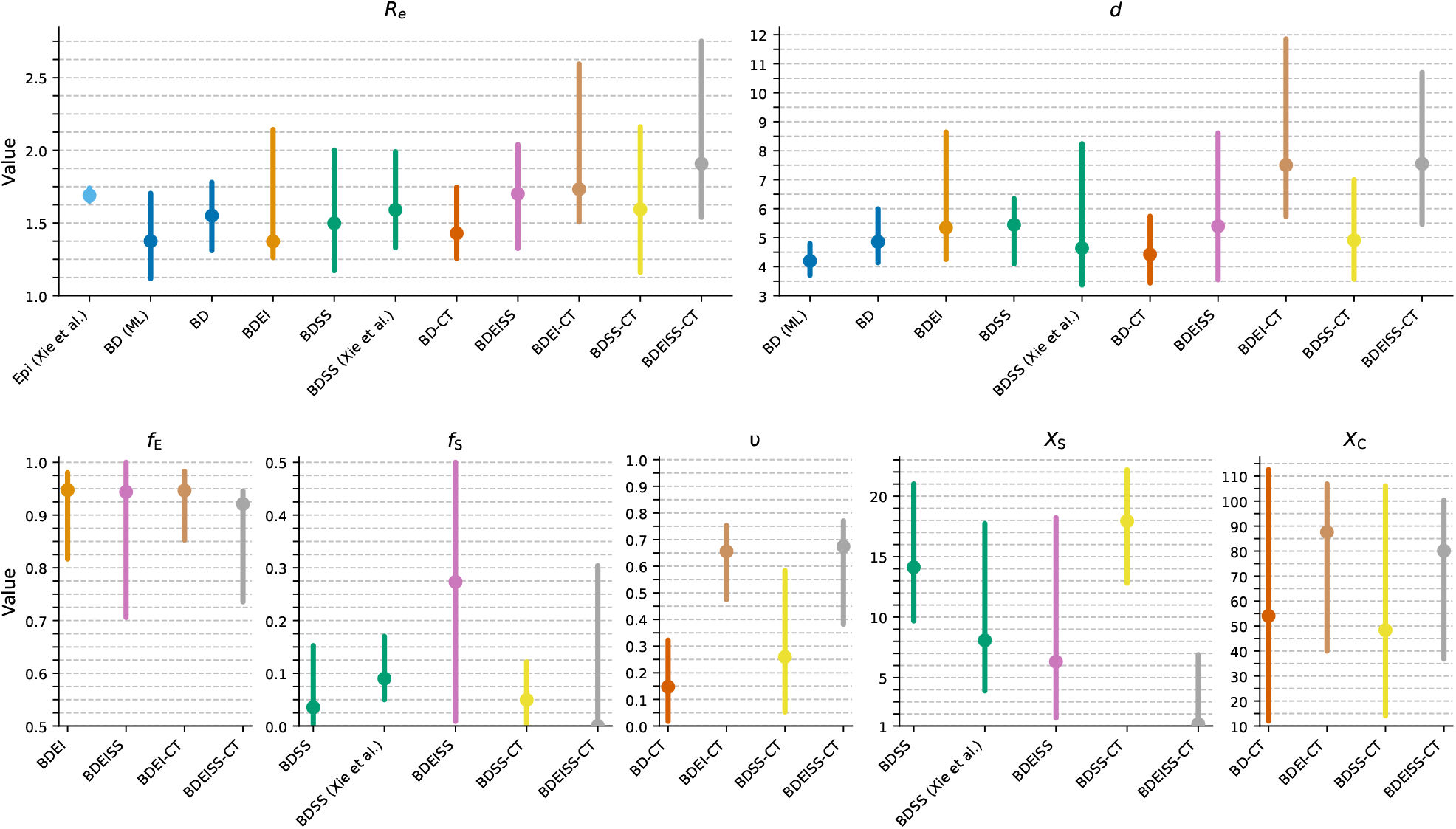
Hong Kong SARS-CoV-2 wave 3 epidemiological parameters and their CIs estimated with different models. Each parameter is represented by a separate plot (parameter name shown in title). The dots correspond to the median estimates, while the whiskers cover the 95% CIs inferred with different estimators. The estimator names are shown on the x-axis. “BD (ML)” is a maximum-likelihood estimator for the BD model [40]. Estimators labeled only with the model name correspond to the DL estimators described in this study. They were trained on nested models for 200-500-tip trees. “BDSS (Xie *et al*.)” is a DL-based inference with the BDSS model, while “Epi (Xie *et al*.)” is an epidemiological inference from incidence data, both described in Xie *et al*. [39].

As we have seen on simulated data, unaccounted contact-tracing might be confused with super-spreading, which explains why our BDEISS estimator and a previous study [39] using a BDSS estimator detected super-spreading in these data. While the *R*_*e*_ estimate by the BDEISS and the BDEI-CT models are close, the underlying mechanisms are very different. In the case of super-spreading (the BDEISS model), the estimated *R*_*e*_ and *d* parameters translate to the infectious period *d*_*inf*_ = 0.3 [days], with an average transmission rate of 5.6 [times per day]. This transmission rate corresponds to the basic transmission rate *λ* = 2.3 [times per day] for regular spreaders *I*, and an increased rate of 14.5 [times per day] for superspreaders (accounting for 27% of the infected population). The infectious time seems to be short, though might be due to the person’s decision to self-isolate or wear a mask soon after the onset of symptoms. In the case of contact tracing (BDEI-CT), the estimated parameters translate to the average infectious period *d*_*inf*_ = 0.4 [days] and a transmission rate *λ* = 4.3 [times per day]. The average infectious period of 0.4 days here means that untraced spreaders *I* typically stay infectious longer than that, while contact-traced ones *I*_*C*_ – shorter (potentially taking care to wear a mask even before the end of incubation). Therefore, with comparable average infectious times and transmission rates, the BDEISS model suggests lower base transmission rate (for *I*) and its increase for superspreaders over homogeneous infectious time, while the BDEI-CT model suggests homogeneous transmission rate over heterogeneous infectious time, with shorter time for contact-traced individuals.

## 4 Discussion

We investigated, using simulated data, the performance of epidemiological parameter estimators of varying complexity in the setting of different epidemiological model misspecifications. On one hand, our results suggest that using simpler estimators (such as BD) on data that features complex epidemic features (e.g., super-spreading) leads to bias in parameter estimation. This bias is dangerous from the public health perspective: For instance using the BD estimator on the data featuring super-spreading, contact tracing or incubation, leads to underestimation of the reproduction number *R*_*e*_ by 10-50%, and hence potential conclusion that the epidemic is better contained that it really is. This results agrees with a previous study of effects of non-accounting for incubation by Park and Koelle [19]. On the other hand, our results suggest that using a more complex estimator on simpler data (e.g., accounting for contact-tracing when it is not present) leads to accurate estimates (e.g., contact tracing probability of zero), however with wider confidence intervals. A similar conclusion was recently achieved for several models in macroevolution domain [1], as well as for nucleotide substitution models for phylogenetic inference [7] in the Bayesian context, provided well-behaved priors are used. However, as our simulations show, to achieve this behavior with complex models in the DL-based context, it is important to have boundary-case data (i.e., corresponding to the nested models) well represented in the training dataset. Moreover, our results show that there is no need for explicit model selection: an estimator trained on the data generated from multiple models performs model selection implicitly and infers the parameters appropriate to the input data.

Our study hence raises awareness of the importance of accounting for as many aspects of the epidemic as possible. Until recently, this strategy was hindered by the mathematical and computational complexity of the models in question. This led to excessive use of the simple BD(SKY) models, and potential underestimates of the key epidemiological parameter values. However, the recent arrival of deep learning to the phylodynamic domain [36, 30, 14, 20] enabled replacing complex likelihood computation with much simpler transmission tree simulation (for training of a deep learning predictor). Deep learning thus opened the way for the design of, and parameter estimation under, more complex models. In particular, the CT-aware models in our study did not have an explicit formula for *R*_*e*_ and *d*, however, these parameter values could be extracted from simulations, and the DL-based predictors managed to accurately estimate them. Estimation of these parameters for CT-aware models would not be possible in the likelihood-based setting.

For our study we trained proof-of-concept deep learning estimators for complex models that did not have a likelihood-based alternative. We used a simple neural network architecture and did not try to achieve the best possible performance for these estimators. Nevertheless even these versions of complex-model estimators outperformed the estimators for simpler models, and especially the maximum likelihood estimator for the BD model. The performance gap might further increase with a better deep learning architecture [20] or larger training dataset size.

We applied our estimators to an empirical data sets (SARS-CoV-2 wave 3 epidemic in Hong-Kong) and obtained biologically reasonable estimates, which were also coherent with the trends observed on simulated data (e.g., lower *R*_*e*_ for simpler models, contact-tracing potentially mistaken for super-spreading).

Our investigation also raises new questions for future studies: Where should one stop when accounting for complex epidemic aspects? And how to ensure all the needed aspects are accounted for? In particular, in the models we considered all the parameters were constant. This is rarely the case in real epidemics, and can become an additional source of bias – or even non-identifyability [16] – in the estimations. Assessing the errors in estimations with BD(EI)(SS)(−CT) models on BD(EI)(SS)(−CT)-SKY data – featuring piecewise-constant changes in rates over time – would be the natural next step. We could detect the mismatch between the training data for the BD and BD-CT estimators and the real SARS-CoV-2 tree by looking at summary statistics. This check is definitely useful, but its performance depends on the choice of summary statistics. For instance, we could not detect a mismatch for the BDSS model, while the presence of incubation is well-established for SARS-CoV-2. Therefore, additional tests will need to be designed in the future.

### 4.1 Conclusion

In conclusion, our results suggest that accounting for as many potential aspects (super-spreading, incubation, contact-tracing) of an epidemic as possible is key even for estimation of epidemiological parameters that do not directly depend on these aspects (e.g., the reproduction number *R*_*e*_). Deep learning helps remove the mathematical difficulty in designing such estimators. If, however, a simpler estimator is used on a potentially more complex epidemic, one needs to keep in mind potential biases in the results (e.g., underestimation of *R*_*e*_). These biases might be present in previous phylodynamic studies that had to use the simpler BD(SKY) model estimator due to its closed-form ODE solution and scalability to large sequence datasets. Simulated data allows to assess potential biases in estimation with such simpler models.

## Data Availability

All code and data produced are available online at https://github.com/modpath/bdeissct

## 5 Competing interests

No competing interest is declared.

## 6 Author contributions statement

A.Z. and R.X. conceived the experiments, A.Z. conducted the experiments, analysed the results and wrote the manuscript. R.X. and O.G. critically reviewed the manuscript, and discussed the project progress.

## 7 Acknowledgments

This work is supported in part by funds from the Marie Sklodowska-Curie Actions (Project No. 101203810, R.X.) and from the French National Research Agency (Project No. ANR-25-CE45-1990 EmPaTHIc, A.Z.). We would like to thank the High-Performance Computational Cluster team of Institut Pasteur.

## Footnotes

1 According to Google scholar (https://scholar.google.com, assessed on August 10, 2026).

## Notes

### Competing Interest Statement

The authors have declared no competing interest.

### Summary of Updates

Added 2 tables, containing model definitions and training parameter ranges; DL architecture was improved for seed stability and median ensemble of 5 independent seed models used instead of a single ones; Results updated slightly according to the new model.

