## Supplementary Material for "Simpler is not always better: Phylodynamic misspecification and deep-learning corrections"

[1] ORCID: 0009-0004-5893-4284 [2] ORCID: 0000-0002-9412-9723 [3] ORCID: 0000-0003-2200-7935

**References** **17**

### S1 MTBD-CT average reproduction number and infection duration calculation and simulation

#### S1.1 Formulas for the MTBD model

A Multi-Type Birth-Death (MTBD) model [9, 6] describes pathogen transmission in a heterogeneous population with $m$ possible states of infected hosts ($1 \leq i \leq m$). It has $2m^2 + m$ parameters:

- $m(m-1)$ host **state change rates** $\mu_{ij} \geq 0$ (from state $i$ to state $j \neq i$, $1 \leq i, j \leq m$), while the total state change rate from a host in state $i$ can be calculated as $\mu_i = \sum\limits_{j=1, j \neq i}^{m} \mu_{ij}$;
- $m^2$ **transmission rates** $\lambda_{ij} \geq 0$ (from host in state $i$ to a newly infected individual in state $j$, $1 \leq i, j \leq m$), while the total transmission rate from a host in state $i$ can be calculated as $\lambda_i = \sum\limits_{j=1}^{m} \lambda_{ij}$;
- $m$ **removal rates** $\psi_i \geq 0$ (at which a host in state $i$ becomes non-infectious, $1 \leq i \leq m$);
- $m$ **sampling probabilities** $0 \leq \rho_i \leq 1$ (with which the pathogen of a removed host in state $i$ may be sampled, $1 \leq i \leq m$).

One can also define the total **exit rate** from state $i$ as $\epsilon_i = \mu_i + \psi_i$, $1 \leq i \leq m$.

Using the above rates we can calculate the **equilibrium frequencies** $\pi_i$ ($1 \leq i \leq m$, $\sum\limits_{1}^{m} \pi_i = 1$) corresponds to the ratios of different states at a given time $t^*$ after these ratios stopped changing (assuming that this may happen). $\pi_i = \frac{N_i(t^*)}{N(t^*)}$, where $N_i(t^*)$ is the number of hosts in state $i$ and $N(t^*)$ is the total number of infected (infectious or not) hosts at time $t^*$. Hence, the derivative of the number of hosts in state $i$ is proportional to the derivative of the total number of infected individuals:

$$dN_i(t^*) = \pi_i dN(t^*). \tag{S1}$$

The number of hosts in state $i$ increases due to state changes from states $j \neq i$ to state $i$ and due to transmissions from states $j$ to state $i$. It decreases due to removal:

$$dN_i(t^*) = \sum_{j=1, j \neq i}^{m} \mu_{ji} N_j(t^*) + \sum_{j=1}^{m} \lambda_{ji} N_j(t^*) - \psi_i N_i(t^*) \tag{S2}$$

State changes do not affect the total number of infected individuals, transmissions increase the total number, and removals decreases it:

$$dN(t^*) = \sum_{i=1}^{m} \lambda_i N_i(t^*) - \sum_{i=1}^{m} \psi_i N_i(t^*). \tag{S3}$$

Combining Eqs. (S1), (S2) and (S3) we can obtain the values of the equilibrium frequencies $\pi_i$.

Model rates and equilibrium frequencies permit us to calculate **state-specific infection durations** (once in this state) $d_i$, by solving the following system of linear equations:

$$d_i = \frac{1}{\epsilon_i} + \sum_{j=1,\, j \neq i}^{m} \frac{\mu_{ij}}{\epsilon_i} d_j \text{ for } 1 \leq i \leq m. \tag{S4}$$

The above equations correspond to the time the person stays in state $i$ ($1/\epsilon_i$) plus the infection duration of the state $j$ to which they transition (weighted by the probability to transition to that state). Using them, one can calculate the **average infection duration** $d$ as a sum of infection durations corresponding to the different recipient states weighted by their equilibrium frequencies. We take only the recipient states as for the non-recipient states the beginning of their infection happens in a different state:

$$d = \frac{\sum\limits_{1 \leq i \leq m,\ \exists 1 \leq j \leq m:\ \lambda_{ji} > 0} \pi_i d_i}{\sum\limits_{1 \leq i \leq m,\ \exists 1 \leq j \leq m:\ \lambda_{ji} > 0} \pi_i}. \tag{S5}$$

To calculate **state-specific reproduction numbers** $R_{ei}$, one needs to solve the following system of linear equations:

$$R_{ei} = \frac{\lambda_i}{\epsilon_i} + \sum_{j=1,\, j\neq i}^{m} \frac{\mu_{ij}}{\epsilon_i} R_{ej} \text{ for } 1 \leq i \leq m. \tag{S6}$$

The above equations correspond to the person transmitting at a rate $\lambda_i$ for the time during which they stay in that state, and then spreading the infection further according to the state $j$ to which they transition (weighted by the probability to transition to that state). One can then calculate the **average reproduction number** $R_e$ as a sum of reproduction numbers corresponding to different recipient states, weighted by their equilibrium frequencies:

$$R_e = \frac{\sum\limits_{1\leq i\leq m,\ \exists 1\leq j\leq m:\ \lambda_{ji}>0} \pi_i R_{ei}}{\sum\limits_{1\leq i\leq m,\ \exists 1\leq j\leq m:\ \lambda_{ji}>0} \pi_i}. \tag{S7}$$

#### S1.2 Formulas for the BDEISS model

The Birth-Death Exposed-Infectious Super-Spreading (BDEISS) model, which we focus on in the main text, is a special case of the MTBD model with $m = 3$ states (exposed $E$, regular spreaders $I$ and superspreaders $S$). It has two non-zero state change rates from state $E$: $\mu_{EI} = (1 - f_S)\mu$ and $\mu_{ES} = f_S\mu$, where the total state change rate from state $E$ is $\mu_E = \mu_{EI} + \mu_{ES} = \mu$. State changes from other states are not possible: $\mu_I = \mu_S = 0$. The BDEISS model has two non-zero transmission rates: from $I$ to $E$ $\lambda_{IE} = \lambda$ and from $S$ to $E$ $\lambda_{SE} = X_S\lambda$, all the other transmission rates being zero: $\lambda_{EE} = \lambda_{EI} = \lambda_{ES} = \lambda_{II} = \lambda_{IS} = \lambda_{SI} = \lambda_{SS} = 0$. There are two equal non-zero removal rates: $\psi_I = \psi_S = \psi$, while the removal rate from the state E is zero: $\psi_E = 0$ (assuming the pathogen needs to incubate in order to become detectable). There are also two equal sampling probabilities $\rho_I = \rho_S = \rho$. (Since $E$ cannot be removed, it also cannot be sampled upon removal.) The total exit rates for this model correspond to $\epsilon_E = \mu$, and $\epsilon_I = \epsilon_S = \psi$.

For this model, the state-specific infection durations (Eq. (S4)) can be calculated as:

$$\begin{cases} d_E &= \frac{1}{\mu} + \frac{1}{\psi} \\ d_I &= \frac{1}{\psi} \\ d_S &= \frac{1}{\psi} \end{cases} \tag{S8}$$

The average duration corresponds to $d_E$, as it is the only recipient state:

$$d = d_{inc} + d_{inf},\ where\ d_{inc} = \frac{1}{\mu},\ d_{inf} = \frac{1}{\psi}. \tag{S9}$$

State-specific reproduction numbers (Eq. (S6)) under the BDEISS model correspond to:

$$\begin{cases} R_{eE} &= (1 - f_S)R_{eI} + f_S R_{eS} \\ R_{eI} &= \frac{\lambda}{\psi} \\ R_{eS} &= \frac{X_S\lambda}{\psi} \end{cases} \tag{S10}$$

Finally, the average reproduction number corresponds to $R_{eE}$:

$$R_e = \Big(1 + (X_S - 1)f_S\Big)\lambda d_{inf} \tag{S11}$$

#### S1.3 Extracting values from simulations for -CT models

Contact-tracing (CT) extension adds $m$ contact-traced states $(1_C, \dots, m_C)$ to an MTBD model with $m$ states $(1, \dots, m)$. It also adds $m(m-1)$ state change rates that mirror the original ones: $\mu_{i_C j_C} = \mu_{ij}$ $(i \neq j)$; $m^2$ transmission rates: $\lambda_{i_C j} = \lambda_{ij}$; $m$ removal rates $\psi_{i_C}$; $m$ sampling probabilities $\rho_{i_C}$ and a contact-tracing probability $\upsilon$. State changes between contact-traced and original states as well as transmissions with a contact-traced recipient are not allowed. State transitions from an original state $i$ to its contact-traced version $i_C$ happen due to the fact that $i$'s contact (the host who infected $i$, or one of the hosts infected by $i$) (a) got removed, and (b) decided to notify $i$ (with a probability $\upsilon$).

The epidemiological process under the MTBD models is memory-less, where the future events depend only on the current infected individual state, and the transmission tree branches are independent. On the contrary, the CT extension adds the dependency between the notifier branch and the contact branches, and keeps the memory of who infected whom. This branch dependency significantly complicates the calculation of equilibrium frequencies, as the transitions to contact-traced states now depend on removal and willing to notify of their contacts, who might be in any state. We did not find a way to derive equations for equilibrium frequencies under the MTBD-CT model. This in turn implied that the formulas for $R_e$ and $d$ were not easily derivable either. Intuitively, we do not know at which point during the infection the notification happens on average, and hence how much contact tracing reduces the average infectious time $d_{inf}$.

Nevertheless, we can extract the values of these parameters from the transmission tree simulations. As we can simulate full transmission trees (including unsampled parts) under given model rate parameters, we can observe the proportion $\xi_{observed}$ of removed hosts among all infected hosts during the epidemic. On average a host is removed before the ones whom they infected. Hence for every host who got removed there are still on average $R_e$ hosts who are infected, and we can estimate $R_e$ as:

$$R_e = \frac{1}{\xi_{observed}}. \tag{S12}$$

For the average infection duration $d$ we have observations of total (till removal) or partial (till the end of the transmission tree, i.e., sampling period) infection time for all the infected hosts. We can estimate $d$ from them with the Kaplan–Meier estimator [5]. However, as our simulations showed, an even more precise way to estimate $d$ for the BDEISS-CT model was to estimate $R_e$ from simulations and then calculate $d$ from Eqs. (S9) and (S11). This is due to the fact that a transmission tree contains more information related to $R_e$ (internal branches corresponding to times between transmissions and external branches, corresponding to times till removal) than to $d$ (only external branches). We checked this approach on trees with known formulas (BDEISS and nested models) and confirmed that $R_e$ and $d$ estimated this way are very close to real values (relative error $< 1\%$, relative bias $< 0.5\%$, Table S1).

Table S1: Accuracy of estimation of the average reproduction number $R_e$ and the average infection duration $d$ from observations on simulated full transmission trees.

| | $R_e$ | | $d$ | |
|---|---|---|---|---|
| BD | 0.6 | (0.3) | 0.6 | (0.3) |
| BDEI | 0.5 | (0.3) | 0.2 | (0.1) |
| BDSS | 0.8 | (0.4) | 0.8 | (0.4) |
| BDEISS | 0.9 | (0.5) | 0.4 | (0.2) |

Rows correspond to the models used for tree generation. Mean relative errors multiplied by 100 (e.g., for the reproduction number: $100|R_{e,observed} - R_{e,true}|/R_{e,true}$) and in parenthesis the corresponding relative biases (e.g., $100(R_{e,observed} - R_{e,true})/R_{e,true}$) are reported for 1 000 test trees generated under each model.

### S2 Summary statistics used for deep learning BD(EI)(SS)(-CT) estimators

The appendix describes the summary statistics used to encode trees for training deep learning BD(EI)(SS)(-CT) parameter estimators. These summary statistics were adapted from Voznica *et al.* [12], who in turn adapted the statistics from Saulnier *et al.* [8]. We normalized these statistics when necessary with respect to the tree size. We also added several contact tracing and incubation-related statistics.

In the following we assume that the statistics are calculated for a tree $\mathscr{T}$ containing $n$ sampled tips. We propose 218 summary statistics to encode such a tree.

#### S2.1 Tree size (1)

We include the tree size measured in the number of tips:

1. number of tips.

#### S2.2 Branch length summary statistic (36)

The following branch length statistics are adapted from Saulnier *et al.* [8]. We first calculate them on the entire tree:

1-3. mean, median and variance of the lengths of the non-root internal nodes;

4-6. mean, median and variance of the lengths of the tips;

7-9. fraction of the corresponding internal and tip node values: mean length of internal nodes by the mean length of tips, and the same for median and variance.

Then we split the total time between the tree start and the last sampled tip into three equal parts, and calculate the same statistics only on branches starting and finishing in the top, middle or bottom parts of the tree:

10-18. same as 1-9 but considering only branches starting and finishing in the top third of the tree;

19-27. same as 1-9 but considering only branches starting and finishing in the middle third of the tree;

28-36. same as 1-9 but considering only branches starting and finishing in the bottom third of the tree.

##### S2.3 Event time summary statistic (10)

The following time-based statistics are normalized by dividing them by the time of the last tip in the tree ($T$).

1-5. normalized mean, minimum, maximum, median and variance of the sampling times of tips.

6-10. normalized mean, minimum, maximum, median and variance of the times of internal nodes.

##### S2.4 4-transmission chain summary statistic (10)

We adapted the transmission chain statistics from Voznica *et al.* [12], defining a $k$-node transmission chain of node $i$ to be the shortest $k$-branch path descending from $i$. We used $k = 4$.

1. number of 4-node transmission chains, divided by the number of internal nodes;

2-6. mean, minimum, maximum, median and variance of branch length sums of all 4-node transmission chains;

7-10. X-th percentile of branch length sums of all 4-node transmission chains, where $X \in \{1, 5, 10, 25\}$.

##### S2.5 Lineage-Through-Time summary statistics (60)

We calculate Lineage-Through-Time (LTT)-based statistics following the ones described by Saulnier *et al.* [8]. However, we normalize them by dividing by the total number of tips $n$. We calculate these statistics for 21 times $t_i$ ($0 \leq i \leq 20$) equally distributed between the tree start ($t_0 = 0$) and the last sampled tip ($t_{20} = T$).

1-20. backward-going time coordinates $T - t_0$, $T - t_1$, ..., $T - t_{19}$ (we do not report $T - t_{20}$ as it is always zero);

21-40. numbers of normalized lineages at times $t_0$, $t_1$, ..., $t_19$.

Additionally, we calculate the following five lineage-related statistics adapted from Saulnier *et al.* [8] four times: (1) on the whole tree ($t_{start} = 0$ and $t_{end} = T$); (2) on the top part of the tree (covering the time interval between $t_{start} = 0$ and $t_{end} = \frac{1}{3}T$); (3) on the middle part of the tree (covering the time interval between $t_{start} = \frac{1}{3}T$ and $t_{end} = \frac{2}{3}T$); and (4) on the bottom part of the tree (covering the time interval between $t_{start} = \frac{2}{3}T$ and $t_{end} = T$):

41-44. $t_{end} - t_{lmax}$, where $t_{lmax}$ is the time when the maximum number of lineages was first achieved;

45-48. maximum number of normalized lineages;

49-52. linear slope between the normalized lineages at times $t_{start}$ and $t_{lmax}$;

53-56. linear slope between the normalized lineages at times $t_{lmax}$ and $t_{end}$;

57-60. ratio between the two linear slopes described above.

#### S2.6 Tree balance summary statistics (10)

We adapted the tree balance summary statistics used in Saulnier *et al.* [8] (they were initially proposed in [7, 2, 1]) and, where applicable, normalized them.

Following Colijn *et al.* [1], we define the node's depth as the number of branches separating it from the tree root: The root has depth 0, its children have depth 1, and so on. We define the tree width at depth $d$ as the number of nodes that have depth $d$.

To normalize balance and depth-related statistics we calculated them on the deepest (and at the same time most unbalanced) tree $\mathscr{T}_{deep}$ with the same number of tips $n$: a ladder tree. To normalize width-related statistics we calculated them on the widest tree $\mathscr{T}_{wide}$ with the same numbers of tips $n$: a balanced tree.

1. Colless value (sum over internal nodes of absolute differences of numbers of tips in their left and right subtrees [2]), divided by the Colless value for $\mathscr{T}_{deep}$ ($\frac{(n-2)(n-1)}{2}$);
2. Sackin value, (i.e., sum (over tips) of the number of internal nodes separating them from the root [7]), divided by the Sackin value for $\mathscr{T}_{deep}$ ($\frac{(n-1)n}{2} - 1$);
3. maximum width, divided it by the maximum width of $\mathscr{T}_{wide}$;
4. maximum node depth, normalized by the maximum depth of $\mathscr{T}_{deep}$ ($n - 1$);
5. maximum width to maximum depth ratio, normalized by this ratio for $\mathscr{T}_{wide}$;
6. maximum width difference between neighbouring depths, divided by this difference for $\mathscr{T}_{wide}$, whose last two layers were reorganized to increase the difference (the tips of the last layer were grouped into one ladderized subtree);
7. fraction of internal nodes that have a tip descendant [1];
8. maximum number of connected internal nodes with a single tip descendant [1], divided it by the number of tips ($n$);
9. fraction of internal nodes that have different number of tips in their smallest and their largest child subtrees [1];
10. mean ratio of min-to-max subtree sizes over all internal nodes [1];

#### S2.7 Topology summary statistics (10)

We propose the following five summary statistics to describe the subtree topologies found in the tree.

1. fraction of tips that are found in cherries;
2. fraction of tips that are found in ladderized triplets;
3. fraction of tips that are found in unresolved triplets;
4. fraction of tips that are found in ladderized quartets;
5. fraction of tips that are found in balanced quartets (with two cherries as children);
6. fraction of tips that are found in fully unresolved quartets (with four children);
7. fraction of tips that are found in partially unresolved quartets with an unresolved triplet and a tip as children;
8. fraction of tips that are found in partially unresolved quartets with a cherry and two tips as three children;
9. fraction of tips that are not part of any subtree of size 2, 3 or 4;
10. fraction of internal nodes that have a sibling internal node.

##### S2.8 Subtree tip time difference summary statistics (80)

We propose the following summary statistics to capture potential contact tracing and incubation patterns. We extract the subtrees of three kinds: cherries, triplets, and internal cherries (i.e., internal nodes with two internal children). For each of them we calculate the average of absolute time differences among all the tip pairs presented in this subtree (e.g., for a triplet containing tips sampled at times $t_1$, $t_2$ and $t_3$, this value would correspond to $\frac{|t_1-t_2|+|t_1-t_3|+|t_2-t_3|}{3}$). We then calculate different metrics on these differences across all subtrees of the same kind.

1-5. mean, minimum, maximum, median, variance of average of tip time differences across all cherries;

6-10. same as above but across all ladderized triplets;

11-15. same as above but across all unresolved triplets;

16-20. same as above but across all fully unresolved quartets;

21-25. same as above but across all internal cherries.

Additionally we calculate the same 25 statistics on reshuffled subtrees. To construct a reshuffled subtree of a certain kind (cherry, triplet, internal cherry) from a real one, we keep one of its (randomly chosen) tips. For each other tip from the real subtree we replace it with a tip (or internal node for an internal cherry), whose time at the beginning of its branch is as close as possible. We ensure than all the tips in the reshuffled subtree are different and that no two tips of the reshuffled subtree belong to the same real subtree of this kind.

25-50. mean, minimum, maximum, median, variance of average of tip time differences across all reshuffled cherries; all reshuffled ladderized triplets; all reshuffled unresolved triplets; all reshuffled fully unresolved quartets; and all reshuffled internal cherries.

We also calculate percentile statistics for them:

51-58. X-th percentile ($X \in 1, 5, 10, 25$) of average of tip time differences across all cherries; and all ladderized triplets;

59-62. X-th percentile ($X \in 75, 90, 95, 99$) of average of tip time differences across all internal cherries;

63-70. X-th percentile ($X \in 1, 5, 10, 25$) of average of tip time differences across all reshuffled cherries; and all reshuffled ladderized triplets;

71-74. X-th percentile ($X \in 75, 90, 95, 99$) of average of tip time differences across all reshuffled internal cherries.

Finally we compare the average tip time differences between real subtrees and their corresponding reshuffled subtrees:

75-76. fraction of cherries/ladderized triplets whose average tip time difference is smaller in real subtrees than in the corresponding reshuffled ones;

77. fraction of internal cherries whose average tip time difference is larger in real subtrees than in the corresponding reshuffled ones;

78-79. p-value of the sign test checking for cherries/reshuffled triplets whether their average tip time differences are indistinguishable in real and reshuffled subtrees (the alternative hypothesis is that some real ones are smaller);

80. p-value of the sign test checking for internal cherries whether their average tip time differences are indistinguishable in real and reshuffled subtrees (the alternative hypothesis is that some real ones are larger).

##### S2.9 Sampling probability (1)

We include the presumed sampling probability $\rho$ as an additional summary statistics, as for the identifiability of the BD(SKY) models and their extensions one of their parameters needs to be fixed [10, 11]. The sampling fraction can often be estimated as the proportion of sampled sequences among the total declared cases.

1. presumed sampling probability.

##### S2.10 Code availability

The tree summary statistic calculator is implemented in Python 3. It uses ETE 3 framework for tree manipulation [4] and NumPy package for array operations [3]. It is available as a command-line program and a Python 3 package via PyPi (treesumstats), and via Docker/Singularity (evolbioinfo/treesumstats). Its source code and the installation and usage documentation are available on GitHub at github.com/modpath/treesumstats.

### S3 Robustness of deep learning estimators

Table S2: Cross-seed convergence stability, measured between the median ensemble and five individual seeds (columns) via mean pairwise difference in the MSSE (Δ MSSE ± standard error) and Two One-Sided Tests (TOST) with an equivalence bound of $\epsilon^2 = 0.02$. Values with $p_{\text{TOST}} < 0.05$ are put in bold.

| estimator | | ensemble vs 1 | | ensemble vs 2 | | ensemble vs 3 | | ensemble vs 4 | | ensemble vs 5 | |
|---|---|---|---|---|---|---|---|---|---|---|---|
| | | Δ MSSE | $p_{TOST}$ | Δ MSSE | $p_{TOST}$ | Δ MSSE | $p_{TOST}$ | Δ MSSE | $p_{TOST}$ | Δ MSSE | $p_{TOST}$ |
| 200–500 tip trees | | | | | | | | | | | |
| BD | pure | -0.010 ± 0.002 | 0.00 | 0.003 ± 0.002 | 0.00 | -0.001 ± 0.002 | 0.00 | -0.000 ± 0.003 | 0.00 | -0.004 ± 0.002 | 0.00 |
| BDEI | pure | -0.002 ± 0.001 | 0.00 | -0.009 ± 0.001 | 0.00 | -0.006 ± 0.001 | 0.00 | 0.003 ± 0.002 | 0.00 | 0.009 ± 0.001 | 0.00 |
| | mixed | -0.006 ± 0.001 | 0.00 | -0.007 ± 0.002 | 0.00 | 0.005 ± 0.002 | 0.00 | -0.001 ± 0.002 | 0.00 | -0.002 ± 0.001 | 0.00 |
| BDSS | pure | 0.015 ± 0.002 | 0.00 | -0.015 ± 0.002 | 0.00 | 0.003 ± 0.002 | 0.00 | -0.006 ± 0.002 | 0.00 | -0.013 ± 0.002 | 0.00 |
| | mixed | 0.000 ± 0.001 | 0.00 | 0.004 ± 0.002 | 0.00 | 0.004 ± 0.002 | 0.00 | -0.013 ± 0.002 | 0.00 | -0.012 ± 0.001 | 0.00 |
| BD-CT | pure | 0.002 ± 0.002 | 0.00 | -0.010 ± 0.002 | 0.00 | 0.002 ± 0.002 | 0.00 | -0.009 ± 0.002 | 0.00 | -0.005 ± 0.002 | 0.00 |
| | mixed | -0.005 ± 0.002 | 0.00 | 0.009 ± 0.002 | 0.00 | -0.009 ± 0.002 | 0.00 | -0.006 ± 0.002 | 0.00 | -0.002 ± 0.002 | 0.00 |
| BDEISS | pure | -0.012 ± 0.001 | 0.00 | -0.004 ± 0.001 | 0.00 | -0.018 ± 0.001 | 0.00 | 0.005 ± 0.001 | 0.00 | 0.007 ± 0.001 | 0.00 |
| | mixed | -0.006 ± 0.001 | 0.00 | -0.004 ± 0.001 | 0.00 | -0.000 ± 0.001 | 0.00 | -0.004 ± 0.001 | 0.00 | -0.002 ± 0.001 | 0.00 |
| BDEI-CT | pure | 0.007 ± 0.001 | 0.00 | -0.001 ± 0.001 | 0.00 | -0.007 ± 0.001 | 0.00 | **-0.021 ± 0.002** | **0.82** | 0.001 ± 0.002 | 0.00 |
| | mixed | 0.012 ± 0.002 | 0.00 | -0.014 ± 0.002 | 0.00 | **-0.019 ± 0.002** | **0.29** | 0.011 ± 0.002 | 0.00 | -0.002 ± 0.002 | 0.00 |
| BDSS-CT | pure | **-0.023 ± 0.002** | **1.00** | 0.002 ± 0.002 | 0.00 | 0.003 ± 0.001 | 0.00 | -0.002 ± 0.001 | 0.00 | 0.000 ± 0.001 | 0.00 |
| | mixed | -0.003 ± 0.001 | 0.00 | -0.009 ± 0.001 | 0.00 | 0.002 ± 0.001 | 0.00 | 0.000 ± 0.001 | 0.00 | -0.005 ± 0.001 | 0.00 |
| BDEISS-CT | pure | -0.010 ± 0.001 | 0.00 | 0.008 ± 0.001 | 0.00 | -0.010 ± 0.001 | 0.00 | -0.005 ± 0.001 | 0.00 | -0.014 ± 0.001 | 0.00 |
| | mixed | -0.004 ± 0.001 | 0.00 | -0.002 ± 0.001 | 0.00 | -0.003 ± 0.001 | 0.00 | -0.005 ± 0.001 | 0.00 | -0.006 ± 0.001 | 0.00 |
| 500–1000 tip trees | | | | | | | | | | | |
| BD | pure | -0.010 ± 0.003 | 0.00 | 0.002 ± 0.002 | 0.00 | -0.007 ± 0.002 | 0.00 | 0.009 ± 0.003 | 0.00 | -0.012 ± 0.003 | 0.00 |
| BDEI | pure | **-0.020 ± 0.002** | **0.56** | 0.000 ± 0.001 | 0.00 | -0.003 ± 0.001 | 0.00 | 0.006 ± 0.002 | 0.00 | 0.001 ± 0.001 | 0.00 |
| | mixed | 0.006 ± 0.002 | 0.00 | **-0.020 ± 0.002** | **0.41** | **0.021 ± 0.002** | **0.79** | -0.013 ± 0.002 | 0.00 | -0.006 ± 0.001 | 0.00 |
| BDSS | pure | 0.009 ± 0.002 | 0.00 | 0.005 ± 0.002 | 0.00 | -0.014 ± 0.002 | 0.00 | -0.018 ± 0.002 | 0.00 | 0.002 ± 0.002 | 0.00 |
| | mixed | -0.007 ± 0.002 | 0.00 | 0.011 ± 0.002 | 0.00 | 0.003 ± 0.001 | 0.00 | **-0.029 ± 0.002** | **1.00** | -0.002 ± 0.002 | 0.00 |
| BD-CT | pure | -0.013 ± 0.002 | 0.00 | -0.005 ± 0.003 | 0.00 | -0.014 ± 0.002 | 0.00 | -0.011 ± 0.002 | 0.00 | 0.006 ± 0.003 | 0.00 |
| | mixed | -0.010 ± 0.002 | 0.00 | -0.007 ± 0.002 | 0.00 | 0.003 ± 0.002 | 0.00 | 0.001 ± 0.002 | 0.00 | -0.011 ± 0.002 | 0.00 |
| BDEISS | pure | -0.009 ± 0.002 | 0.00 | -0.002 ± 0.001 | 0.00 | -0.014 ± 0.001 | 0.00 | 0.008 ± 0.001 | 0.00 | -0.002 ± 0.001 | 0.00 |
| | mixed | -0.010 ± 0.001 | 0.00 | -0.008 ± 0.001 | 0.00 | 0.000 ± 0.001 | 0.00 | -0.003 ± 0.001 | 0.00 | 0.001 ± 0.001 | 0.00 |
| BDEI-CT | pure | 0.004 ± 0.003 | 0.00 | **-0.022 ± 0.002** | **0.97** | -0.010 ± 0.002 | 0.00 | -0.013 ± 0.002 | 0.00 | **0.021 ± 0.003** | **0.82** |
| | mixed | -0.002 ± 0.002 | 0.00 | -0.012 ± 0.002 | 0.00 | -0.011 ± 0.004 | 0.00 | -0.015 ± 0.002 | 0.00 | -0.003 ± 0.003 | 0.00 |
| BDSS-CT | pure | -0.006 ± 0.001 | 0.00 | -0.007 ± 0.001 | 0.00 | -0.009 ± 0.001 | 0.00 | 0.005 ± 0.001 | 0.00 | -0.003 ± 0.001 | 0.00 |
| | mixed | -0.011 ± 0.001 | 0.00 | 0.001 ± 0.001 | 0.00 | 0.003 ± 0.001 | 0.00 | -0.009 ± 0.001 | 0.00 | 0.005 ± 0.002 | 0.00 |
| BDEISS-CT | pure | 0.004 ± 0.001 | 0.00 | -0.016 ± 0.001 | 0.00 | -0.002 ± 0.001 | 0.00 | -0.009 ± 0.001 | 0.00 | -0.007 ± 0.001 | 0.00 |
| | mixed | -0.005 ± 0.001 | 0.00 | -0.000 ± 0.001 | 0.00 | -0.003 ± 0.001 | 0.00 | -0.006 ± 0.001 | 0.00 | -0.003 ± 0.001 | 0.00 |
| 1000–2000 tip trees | | | | | | | | | | | |
| BD | pure | -0.002 ± 0.002 | 0.00 | **-0.026 ± 0.003** | **1.00** | -0.008 ± 0.002 | 0.00 | 0.002 ± 0.002 | 0.00 | 0.014 ± 0.002 | 0.00 |
| BDEI | pure | **0.018 ± 0.003** | **0.14** | **-0.026 ± 0.002** | **1.00** | **0.032 ± 0.003** | **1.00** | -0.017 ± 0.002 | 0.00 | **-0.019 ± 0.002** | **0.27** |
| | mixed | **-0.022 ± 0.002** | **0.92** | **-0.027 ± 0.002** | **1.00** | 0.005 ± 0.002 | 0.00 | **0.024 ± 0.003** | **1.00** | -0.005 ± 0.002 | 0.00 |
| BDSS | pure | 0.004 ± 0.002 | 0.00 | 0.008 ± 0.003 | 0.00 | -0.014 ± 0.002 | 0.00 | -0.015 ± 0.002 | 0.00 | -0.002 ± 0.002 | 0.00 |
| | mixed | -0.015 ± 0.002 | 0.00 | 0.003 ± 0.002 | 0.00 | -0.009 ± 0.002 | 0.00 | -0.016 ± 0.002 | 0.00 | 0.013 ± 0.002 | 0.00 |
| BD-CT | pure | 0.014 ± 0.002 | 0.00 | **-0.025 ± 0.003** | **1.00** | 0.002 ± 0.003 | 0.00 | -0.003 ± 0.004 | 0.00 | **-0.027 ± 0.002** | **1.00** |
| | mixed | -0.007 ± 0.003 | 0.00 | -0.017 ± 0.002 | 0.00 | 0.002 ± 0.004 | 0.00 | -0.007 ± 0.002 | 0.00 | -0.015 ± 0.002 | 0.00 |
| BDEISS | pure | 0.009 ± 0.002 | 0.00 | -0.014 ± 0.002 | 0.00 | -0.003 ± 0.001 | 0.00 | -0.008 ± 0.001 | 0.00 | 0.002 ± 0.001 | 0.00 |
| | mixed | -0.005 ± 0.001 | 0.00 | 0.005 ± 0.001 | 0.00 | -0.000 ± 0.001 | 0.00 | -0.007 ± 0.001 | 0.00 | -0.007 ± 0.001 | 0.00 |
| BDEI-CT | pure | -0.014 ± 0.003 | 0.00 | **0.042 ± 0.003** | **1.00** | **-0.038 ± 0.005** | **1.00** | **-0.042 ± 0.003** | **1.00** | **0.042 ± 0.004** | **1.00** |
| | mixed | -0.008 ± 0.004 | 0.00 | **-0.037 ± 0.004** | **1.00** | **-0.080 ± 0.005** | **1.00** | **0.042 ± 0.004** | **1.00** | **0.028 ± 0.003** | **1.00** |
| BDSS-CT | pure | -0.006 ± 0.001 | 0.00 | -0.014 ± 0.002 | 0.00 | -0.007 ± 0.002 | 0.00 | 0.001 ± 0.002 | 0.00 | 0.009 ± 0.002 | 0.00 |
| | mixed | -0.006 ± 0.001 | 0.00 | -0.006 ± 0.001 | 0.00 | 0.000 ± 0.001 | 0.00 | -0.015 ± 0.002 | 0.00 | 0.004 ± 0.001 | 0.00 |
| BDEISS-CT | pure | -0.006 ± 0.001 | 0.00 | -0.008 ± 0.001 | 0.00 | -0.003 ± 0.001 | 0.00 | -0.018 ± 0.001 | 0.00 | 0.004 ± 0.001 | 0.00 |
| | mixed | -0.003 ± 0.001 | 0.00 | -0.003 ± 0.001 | 0.00 | -0.004 ± 0.001 | 0.00 | -0.002 ± 0.001 | 0.00 | -0.003 ± 0.001 | 0.00 |
| 2000–5000 tip trees | | | | | | | | | | | |
| BD | pure | -0.009 ± 0.008 | 0.00 | **0.031 ± 0.003** | **1.00** | **-0.038 ± 0.004** | **1.00** | 0.005 ± 0.003 | 0.00 | **-0.028 ± 0.003** | **1.00** |
| BDEI | pure | **-0.055 ± 0.003** | **1.00** | **-0.056 ± 0.007** | **1.00** | -0.016 ± 0.003 | 0.00 | **0.039 ± 0.003** | **1.00** | -0.006 ± 0.002 | 0.00 |
| | mixed | 0.004 ± 0.002 | 0.00 | **-0.021 ± 0.003** | **0.74** | 0.011 ± 0.003 | 0.00 | **-0.029 ± 0.003** | **1.00** | 0.001 ± 0.002 | 0.00 |
| BDSS | pure | **-0.029 ± 0.003** | **1.00** | -0.003 ± 0.002 | 0.00 | 0.009 ± 0.003 | 0.00 | -0.011 ± 0.003 | 0.00 | -0.002 ± 0.002 | 0.00 |
| | mixed | 0.001 ± 0.002 | 0.00 | 0.014 ± 0.002 | 0.00 | **-0.038 ± 0.004** | **1.00** | 0.005 ± 0.002 | 0.00 | -0.012 ± 0.002 | 0.00 |
| BD-CT | pure | -0.009 ± 0.004 | 0.00 | 0.014 ± 0.004 | 0.00 | -0.015 ± 0.003 | 0.00 | **-0.058 ± 0.004** | **1.00** | **-0.030 ± 0.004** | **1.00** |
| | mixed | -0.001 ± 0.003 | 0.00 | **-0.021 ± 0.004** | **0.72** | **-0.063 ± 0.006** | **1.00** | -0.005 ± 0.002 | 0.00 | 0.000 ± 0.003 | 0.00 |
| BDEISS | pure | 0.005 ± 0.001 | 0.00 | -0.005 ± 0.001 | 0.00 | 0.007 ± 0.002 | 0.00 | -0.009 ± 0.001 | 0.00 | **-0.023 ± 0.002** | **1.00** |
| | mixed | 0.008 ± 0.002 | 0.00 | **-0.024 ± 0.003** | **0.99** | -0.012 ± 0.002 | 0.00 | -0.002 ± 0.001 | 0.00 | 0.003 ± 0.001 | 0.00 |
| BDEI-CT | pure | **0.020 ± 0.003** | **0.45** | **-0.043 ± 0.003** | **1.00** | **0.023 ± 0.005** | **0.87** | **-0.033 ± 0.004** | **1.00** | **0.062 ± 0.005** | **1.00** |
| | mixed | -0.011 ± 0.004 | 0.00 | **-0.072 ± 0.004** | **1.00** | **0.095 ± 0.005** | **1.00** | **-0.058 ± 0.004** | **1.00** | 0.010 ± 0.005 | 0.00 |
| BDSS-CT | pure | -0.007 ± 0.002 | 0.00 | **-0.024 ± 0.002** | **1.00** | -0.005 ± 0.002 | 0.00 | -0.010 ± 0.002 | 0.00 | 0.001 ± 0.002 | 0.00 |
| | mixed | **0.023 ± 0.002** | **1.00** | -0.013 ± 0.002 | 0.00 | -0.010 ± 0.002 | 0.00 | 0.010 ± 0.001 | 0.00 | **-0.022 ± 0.002** | **1.00** |
| BDEISS-CT | pure | -0.017 ± 0.001 | 0.00 | -0.002 ± 0.001 | 0.00 | 0.001 ± 0.001 | 0.00 | -0.011 ± 0.001 | 0.00 | -0.005 ± 0.001 | 0.00 |
| | mixed | -0.002 ± 0.001 | 0.00 | -0.002 ± 0.001 | 0.00 | -0.004 ± 0.001 | 0.00 | -0.003 ± 0.001 | 0.00 | -0.002 ± 0.000 | 0.00 |

We evaluated DL estimator accuracy and seed stability with a the Mean Squared Standardized Error (MSSE) computed across target parameters. For estimator for epidemiological model $m$ (e.g., BDEI) with $K$ target parameters (e.g., for the BDEI model $K = 3$: $R_e$, $d$, and $f_e$) on tree $i$ the $MSSE_{m,i}$ is defined as:

$$MSSE_{m,i} = \frac{1}{K}\sum_{k=1}^{K}\Big(\frac{\hat{Y}_{m,i,k} - Y_{m,i,k}}{s_{Y,k}}\Big)^2, \tag{S13}$$

where $\hat{Y}_{m,i,k}$ is the estimated value of the $k^{th}$ parameter, $Y_{m,i,k}$ is the true value of this parameter, and $s_{Y,k}$ is the sample standard deviation.

We performed the benchmark across four tree size categories (200–500, 500–1 000, 1 000–2 000, and 2 000–5 000 leaves) across 15 estimation tasks (estimators for all the 8 BD(EI)(SS)(-CT) models flavors, both pure and mixed), each on a test set of $N = 8,000$ examples (composed of 1, 000 test trees generated under each of the 8 BD(EI)(SS)(-CT) models flavors of the corresponding size). For each evaluation, five single-seed DL estimators were benchmarked against a point-wise median ensemble estimator. Practical equivalence between the median ensemble and individual seeds was formally evaluated using Two One-Sided Tests (TOST) with an equivalence bound of $\epsilon^2 = 0.02$ on standardized error ($p_{\text{TOST}} < 0.05$). The results are shown in Table S2.

On small trees (200–500 leaves) complete cross-seed reproducibility was achieved: TOST confirmed practical performance equivalence across all individual seeds and the median ensemble in over 95% of evaluated tasks, with mean difference in MSSE $\Delta < 0.015$ units. As dataset scale increased to 2 000–5 000 leaves, task difficulty scaled selectively: estimators for the BD-CT and BDEI-CT models exhibited increased single-seed optimization sensitivity ($\Delta \approx 0.03 - -0.09$), whereas BDEISS and BDEISS-CT maintained exceptional cross-seed TOST equivalence even on large trees ($p_{\text{TOST}} < 0.0001$, $\Delta \leq 0.0043$). Across all tree sizes and model complexities, the median ensemble consistently filtered out single-seed optimization drift, keeping predictions centered within the optimal seed cluster and confirming its reliability for large-scale phylodynamic inference.

### S4 Supplementary tables

The following tables represent the performance of different BD(EI)(SS)(-CT) estimators on transmission tree data generated under different flavors of the BD(EI)(SS)(-CT) model, representing all the combinations of presence and absence of super-spreading, incubation, and contact-tracing. We looked at how reliably one can estimate different model parameters with (miss-)specified estimators (see Tables S3-S9 for errors and biases, and Tables S10-S16 for confidence intervals (CIs)), how the tree size influences estimations (different tree sizes are reported in each table), and how estimators trained only on the matching model compare to those trained on a collection of matching and its nested models (pure vs mixed in each table). For the reproduction number $R_e$ and the infection duration $d$ we also compared the performance of a maximum-likelihood (ML) based BD estimator to its deep learning (DL) version (BD column in Tables S3 and S4). For other models ML-based estimators were not easily available.

The results are detailed in the Main text, but the general conclusions were that (i) DL performs as well as ML, (ii) mixed estimators generalize better to nested models than pure ones, (iii) misspecification of the epidemiological model leads to bias in estimation even for the parameters that are shared between different models (e.g., $R_e$ and $d$), (iv) estimates under correct models improve with the tree size (lower errors and narrower CIs on larger trees), (v) accounting for additional epidemiological aspects (i.e., more general model, e.g., BDEISS instead of BD) increases CI width but not the errors.

Table S3: Estimation errors for the average reproduction number $R_e$ for transmission trees generated under different models (rows) and different estimators (columns). The estimator type – maximum-likelihood (ML) or deep-learning-based (DL); pure if the estimator was trained on the corresponding model, or mixed if it was trained on the corresponding and its nested models – is specified below its model.

| | BD | | BDEI | | BDSS | | BDEISS | | BD-CT | | BDEI-CT | | BDSS-CT | | BDEISS-CT | |
|---|---|---|---|---|---|---|---|---|---|---|---|---|---|---|---|---|
| | ML | DL (pure) | DL (pure) | DL (mixed) | DL (pure) | DL (mixed) | DL (pure) | DL (mixed) | DL (pure) | DL (mixed) | DL (pure) | DL (mixed) | DL (pure) | DL (mixed) | DL (pure) | DL (mixed) |
| 200-500-tip trees | | | | | | | | | | | | | | | | |
| BD | **10 (2)** | **9 (0)** | **10 (-5)** | **9 (-1)** | **10 (3)** | **9 (1)** | **9 (-1)** | **9 (0)** | **9 (2)** | **9 (0)** | **9 (-2)** | **9 (-1)** | **11 (6)** | **9 (1)** | **9 (1)** | **9 (0)** |
| BDEI | 21 (-11) | 19 (-12) | **10 (0)** | **10 (1)** | 14 (-1) | 13 (-2) | **11 (1)** | **11 (1)** | 19 (-9) | 19 (-12) | **11 (2)** | **11 (1)** | 14 (1) | 13 (-3) | **12 (2)** | **11 (1)** |
| BDSS | 26 (-25) | 25 (-24) | 18 (-16) | 19 (-18) | **11 (-1)** | **12 (-1)** | **13 (-8)** | **12 (-2)** | 24 (-23) | 25 (-24) | 15 (-11) | 17 (-13) | **12 (1)** | **12 (-1)** | **13 (-8)** | **12 (-2)** |
| BDEISS | 29 (-28) | 28 (-27) | 17 (7) | 18 (8) | 18 (4) | 17 (5) | **12 (1)** | **13 (1)** | 27 (-24) | 28 (-26) | 19 (12) | 18 (11) | 17 (3) | 17 (3) | **13 (1)** | **14 (2)** |
| BD-CT | 14 (-12) | 14 (-11) | 15 (-12) | 13 (-8) | 13 (-6) | 13 (-10) | 13 (-9) | 13 (-9) | **10 (1)** | **11 (0)** | **11 (-1)** | **11 (0)** | **12 (4)** | **11 (1)** | **11 (1)** | **11 (0)** |
| BDEI-CT | 21 (-18) | 20 (-17) | 13 (-8) | 13 (-7) | 14 (-4) | 13 (-5) | 13 (-7) | 13 (-6) | 18 (-11) | 19 (-13) | **11 (0)** | **11 (0)** | 14 (0) | 13 (-2) | **11 (0)** | **11 (0)** |
| BDSS-CT | 27 (-27) | 26 (-25) | 20 (-15) | 22 (-18) | 15 (-4) | 15 (-4) | 16 (-5) | 15 (-4) | 21 (-17) | 21 (-17) | 15 (-6) | 16 (-8) | **13 (0)** | **14 (-1)** | **14 (-3)** | **14 (-1)** |
| BDEISS-CT | 29 (-28) | 28 (-27) | 18 (0) | 19 (2) | 15 (0) | 15 (2) | 14 (-3) | 14 (-2) | 24 (-22) | 25 (-23) | 18 (11) | 18 (10) | 15 (1) | 15 (1) | **13 (1)** | **13 (2)** |
| 500-1000-tip trees | | | | | | | | | | | | | | | | |
| BD | **7 (1)** | **6 (0)** | **7 (-4)** | **6 (-1)** | **7 (2)** | **6 (0)** | **7 (0)** | **7 (0)** | **7 (2)** | **6 (0)** | **7 (-1)** | **6 (0)** | **8 (3)** | **6 (1)** | **7 (1)** | **7 (0)** |
| BDEI | 19 (-12) | 18 (-12) | **7 (0)** | **7 (1)** | 12 (-3) | 11 (-3) | **8 (0)** | **8 (1)** | 18 (-10) | 18 (-11) | **8 (2)** | **8 (1)** | 12 (-2) | 11 (-3) | **8 (1)** | **8 (1)** |
| BDSS | 26 (-26) | 25 (-24) | 18 (-17) | 18 (-18) | **8 (-1)** | **8 (-1)** | **10 (-6)** | **8 (-1)** | 23 (-23) | 25 (-24) | 13 (-11) | 17 (-15) | **9 (0)** | **8 (0)** | **10 (-7)** | **9 (-1)** |
| BDEISS | 29 (-28) | 27 (-26) | 16 (8) | 18 (11) | 16 (4) | 16 (4) | **9 (0)** | **10 (1)** | 25 (-23) | 26 (-25) | 22 (18) | 20 (17) | 15 (3) | 16 (5) | **10 (1)** | **10 (1)** |
| BD-CT | 14 (-13) | 13 (-12) | 13 (-7) | 12 (-7) | 12 (-9) | 13 (-11) | 10 (-5) | 12 (-10) | **7 (0)** | **8 (0)** | **8 (0)** | **8 (0)** | **8 (2)** | **8 (1)** | **8 (1)** | **8 (0)** |
| BDEI-CT | 20 (-19) | 19 (-17) | 12 (-9) | 10 (-7) | 12 (-7) | 12 (-7) | 11 (-8) | 10 (-7) | 17 (-12) | 17 (-13) | **8 (0)** | **8 (0)** | 11 (-3) | 11 (-3) | **8 (0)** | **8 (0)** |
| BDSS-CT | 28 (-27) | 26 (-25) | 19 (-16) | 20 (-16) | 12 (-6) | 12 (-6) | 13 (-3) | 12 (-5) | 19 (-16) | 20 (-15) | 12 (-6) | 14 (-9) | **10 (0)** | **10 (0)** | **10 (-2)** | **10 (0)** |
| BDEISS-CT | 30 (-30) | 28 (-27) | 17 (-2) | 18 (3) | 12 (-2) | 12 (-2) | 11 (-5) | 10 (-4) | 23 (-21) | 24 (-23) | 20 (16) | 19 (14) | 12 (0) | 12 (1) | **9 (0)** | **9 (0)** |
| 1000-2000-tip trees | | | | | | | | | | | | | | | | |
| BD | **5 (0)** | **4 (0)** | **5 (-2)** | **5 (0)** | **5 (1)** | **5 (0)** | **5 (-2)** | **5 (-1)** | **5 (1)** | **5 (0)** | **5 (-1)** | **5 (0)** | **6 (2)** | **5 (0)** | **5 (0)** | **5 (0)** |
| BDEI | 18 (-12) | 17 (-12) | **5 (0)** | **6 (0)** | 10 (-2) | 10 (-3) | **6 (0)** | **6 (0)** | 18 (-10) | 18 (-12) | **6 (1)** | **6 (0)** | 11 (-3) | 10 (-3) | **6 (0)** | **6 (0)** |
| BDSS | 26 (-26) | 24 (-24) | 15 (-14) | 18 (-17) | **6 (0)** | **7 (0)** | **8 (-5)** | **7 (-1)** | 24 (-24) | 24 (-24) | 12 (-9) | 16 (-14) | **7 (1)** | **7 (0)** | **9 (-7)** | **7 (-1)** |
| BDEISS | 29 (-29) | 26 (-25) | 17 (10) | 19 (13) | 15 (6) | 16 (6) | **7 (0)** | **7 (0)** | 26 (-25) | 27 (-26) | 28 (26) | 22 (19) | 14 (4) | 16 (5) | **7 (1)** | **8 (0)** |
| BD-CT | 14 (-14) | 12 (-12) | 18 (6) | 12 (-3) | 12 (-10) | 13 (-13) | 10 (-9) | 11 (-10) | **5 (0)** | **5 (0)** | **6 (0)** | **6 (0)** | **6 (1)** | **6 (0)** | **6 (0)** | **6 (0)** |
| BDEI-CT | 20 (-19) | 18 (-17) | 11 (-7) | 10 (-8) | 10 (-7) | 11 (-8) | 11 (-9) | 10 (-8) | 17 (-12) | 17 (-13) | **6 (0)** | **6 (0)** | 10 (-3) | 10 (-4) | **6 (-1)** | **6 (0)** |
| BDSS-CT | 28 (-28) | 24 (-23) | 21 (2) | 19 (-11) | 10 (-7) | 10 (-7) | 12 (-2) | 10 (-5) | 18 (-14) | 18 (-14) | 10 (-5) | 11 (-8) | **7 (0)** | **7 (0)** | **7 (-2)** | **7 (0)** |
| BDEISS-CT | 30 (-30) | 26 (-26) | 16 (-1) | 18 (2) | 10 (-1) | 11 (-1) | 10 (-7) | 9 (-6) | 24 (-22) | 25 (-24) | 24 (21) | 21 (18) | 11 (1) | 11 (1) | **7 (0)** | **7 (0)** |
| 2000-5000-tip trees | | | | | | | | | | | | | | | | |
| BD | **3 (0)** | **3 (0)** | **4 (-2)** | **3 (0)** | **4 (0)** | **3 (0)** | **4 (-1)** | **3 (0)** | **3 (1)** | **3 (0)** | **4 (-1)** | **3 (0)** | **4 (1)** | **3 (0)** | **4 (0)** | **3 (0)** |
| BDEI | 18 (-12) | 18 (-13) | **4 (0)** | **4 (0)** | 10 (-3) | 11 (-3) | **4 (0)** | **4 (0)** | 16 (-7) | 18 (-12) | **4 (1)** | **4 (0)** | 11 (-3) | 10 (-3) | **5 (1)** | **4 (0)** |
| BDSS | 26 (-26) | 23 (-22) | 12 (-8) | 19 (-19) | **4 (0)** | **5 (0)** | **6 (-4)** | **5 (-1)** | 22 (-22) | 23 (-23) | 11 (-9) | 15 (-14) | **5 (0)** | **5 (0)** | **7 (-5)** | **5 (-1)** |
| BDEISS | 29 (-29) | 26 (-26) | 18 (14) | 23 (19) | 15 (7) | 17 (7) | **5 (0)** | **5 (0)** | 23 (-16) | 26 (-25) | 32 (30) | 30 (28) | 15 (6) | 16 (7) | **5 (1)** | **5 (0)** |
| BD-CT | 14 (-14) | 12 (-12) | 22 (15) | 15 (1) | 11 (-10) | 12 (-11) | 7 (-6) | 9 (-8) | **4 (0)** | **4 (0)** | **4 (1)** | **4 (0)** | **4 (0)** | **4 (0)** | **4 (1)** | **4 (0)** |
| BDEI-CT | 20 (-19) | 18 (-17) | 11 (-5) | 10 (-8) | 10 (-8) | 10 (-6) | 9 (-8) | 8 (-7) | 15 (-10) | 14 (-12) | **4 (0)** | **4 (0)** | 9 (-4) | 9 (-4) | **4 (0)** | **4 (0)** |
| BDSS-CT | 28 (-28) | 24 (-24) | 38 (29) | 25 (-2) | 10 (-5) | 9 (-4) | 12 (2) | 9 (-3) | 18 (-13) | 18 (-16) | 9 (-1) | 10 (-5) | **5 (0)** | **5 (0)** | **5 (-1)** | **5 (0)** |
| BDEISS-CT | 30 (-30) | 27 (-26) | 18 (8) | 21 (9) | 9 (0) | 11 (2) | 9 (-7) | 8 (-6) | 21 (-17) | 23 (-22) | 27 (26) | 26 (24) | 11 (3) | 10 (2) | **5 (0)** | **6 (0)** |

Mean absolute percentage errors, $100|R_{e_{estimated}} - R_{e_{true}}|/R_{e_{true}}$, and in parenthesis the corresponding biases, $100(R_{e_{estimated}} - R_{e_{true}})/R_{e_{true}}$, are reported for 1000 trees generated under each dataset for each estimator.
The errors and biases of the estimators corresponding to or generalizing the model that generated the data are shown in bold.

Table S4: Estimation errors for the average infection time $d$ for transmission trees generated under different models (rows) and different estimators (columns). The estimator type – maximum-likelihood (ML) or deep-learning-based (DL); pure if the estimator was trained on the corresponding model, or mixed if it was trained on the corresponding and its nested models – is specified below its model.

| | BD | | BDEI | | BDSS | | BDEISS | | BD-CT | | BDEI-CT | | BDSS-CT | | BDEISS-CT | |
|---|---|---|---|---|---|---|---|---|---|---|---|---|---|---|---|---|
| | ML | DL (pure) | DL (pure) | DL (mixed) | DL (pure) | DL (mixed) | DL (pure) | DL (mixed) | DL (pure) | DL (mixed) | DL (pure) | DL (mixed) | DL (pure) | DL (mixed) | DL (pure) | DL (mixed) |
| 200-500-tip trees | | | | | | | | | | | | | | | | |
| BD | **9 (1)** | **8 (0)** | **27 (-27)** | **9 (-4)** | **9 (5)** | **8 (1)** | **22 (-21)** | **9 (-3)** | **8 (2)** | **8 (0)** | **27 (-27)** | **9 (-4)** | **11 (8)** | **8 (1)** | **21 (-20)** | **9 (-3)** |
| BDEI | 39 (26) | 37 (24) | **9 (-1)** | **13 (5)** | 48 (43) | 45 (40) | **10 (1)** | **13 (6)** | 38 (27) | 37 (24) | **10 (2)** | **13 (6)** | 51 (47) | 44 (40) | **11 (3)** | **14 (6)** |
| BDSS | 32 (-32) | 31 (-31) | 45 (-45) | 37 (-37) | **11 (-1)** | **11 (-1)** | **23 (-22)** | **13 (-4)** | 30 (-29) | 31 (-31) | 40 (-40) | 35 (-35) | **11 (-1)** | **11 (-2)** | **27 (-26)** | **13 (-5)** |
| BDEISS | 25 (-5) | 25 (-4) | 22 (5) | 20 (6) | 58 (51) | 57 (51) | **10 (0)** | **13 (3)** | 24 (-1) | 26 (-3) | 30 (20) | 26 (17) | 55 (47) | 57 (50) | **11 (1)** | **13 (3)** |
| BD-CT | 15 (-13) | 14 (-12) | 24 (-23) | 14 (-12) | 13 (-7) | 14 (-11) | 21 (-20) | 15 (-13) | **9 (0)** | **10 (0)** | **19 (-17)** | **10 (-2)** | **11 (4)** | **10 (1)** | **16 (-14)** | **10 (-1)** |
| BDEI-CT | 23 (1) | 23 (2) | 13 (-9) | 15 (-2) | 28 (21) | 25 (19) | 13 (-6) | 17 (1) | 27 (9) | 27 (8) | **10 (0)** | **12 (3)** | 33 (26) | 30 (23) | **11 (0)** | **13 (3)** |
| BDSS-CT | 31 (-31) | 30 (-29) | 34 (-33) | 31 (-30) | 14 (-4) | 15 (-3) | 20 (-12) | 15 (-5) | 23 (-21) | 24 (-21) | 28 (-23) | 25 (-20) | **12 (0)** | **13 (-1)** | **19 (-15)** | **13 (-3)** |
| BDEISS-CT | 22 (-14) | 22 (-13) | 20 (-2) | 19 (1) | 39 (31) | 40 (32) | 12 (-4) | 15 (1) | 22 (-7) | 23 (-9) | 28 (22) | 26 (20) | 39 (29) | 39 (31) | **10 (0)** | **13 (3)** |
| 500-1000-tip trees | | | | | | | | | | | | | | | | |
| BD | **6 (1)** | **5 (0)** | **22 (-22)** | **6 (-1)** | **6 (2)** | **6 (0)** | **18 (-16)** | **7 (-1)** | **6 (2)** | **6 (0)** | **24 (-24)** | **7 (-2)** | **8 (5)** | **6 (1)** | **19 (-18)** | **7 (-2)** |
| BDEI | 37 (25) | 36 (24) | **7 (0)** | **8 (2)** | 45 (40) | 43 (39) | **7 (1)** | **9 (3)** | 36 (26) | 36 (24) | **7 (1)** | **8 (3)** | 46 (41) | 43 (39) | **8 (2)** | **9 (3)** |
| BDSS | 32 (-32) | 32 (-31) | 41 (-41) | 34 (-34) | **8 (-1)** | **8 (-1)** | **16 (-14)** | **9 (-2)** | 30 (-30) | 31 (-31) | 39 (-39) | 33 (-33) | **8 (-1)** | **8 (-1)** | **21 (-20)** | **9 (-2)** |
| BDEISS | 24 (-6) | 24 (-5) | 23 (8) | 23 (10) | 60 (54) | 59 (54) | **8 (0)** | **9 (2)** | 24 (-2) | 25 (-3) | 36 (28) | 33 (25) | 55 (47) | 60 (54) | **9 (1)** | **10 (2)** |
| BD-CT | 14 (-14) | 14 (-13) | 15 (-12) | 13 (-11) | 13 (-11) | 14 (-12) | 13 (-11) | 14 (-13) | **7 (0)** | **7 (0)** | **15 (-13)** | **7 (-1)** | **8 (2)** | **7 (0)** | **14 (-12)** | **7 (-1)** |
| BDEI-CT | 22 (0) | 22 (1) | 12 (-8) | 15 (-3) | 24 (18) | 23 (17) | 12 (-5) | 15 (0) | 26 (9) | 27 (8) | **7 (0)** | **8 (1)** | 30 (23) | 29 (23) | **8 (-1)** | **9 (2)** |
| BDSS-CT | 32 (-31) | 31 (-30) | 26 (-19) | 28 (-27) | 13 (-4) | 13 (-4) | 17 (-4) | 13 (-5) | 23 (-19) | 23 (-20) | 27 (-21) | 24 (-20) | **9 (0)** | **9 (0)** | **14 (-12)** | **10 (-1)** |
| BDEISS-CT | 22 (-16) | 21 (-14) | 19 (0) | 20 (3) | 38 (30) | 38 (31) | 10 (-4) | 12 (-1) | 21 (-8) | 23 (-9) | 34 (29) | 30 (25) | 38 (28) | 39 (31) | **8 (0)** | **9 (1)** |
| 1000-2000-tip trees | | | | | | | | | | | | | | | | |
| BD | **4 (0)** | **4 (0)** | **16 (-16)** | **4 (-1)** | **5 (1)** | **4 (0)** | **17 (-17)** | **5 (-1)** | **4 (1)** | **4 (0)** | **21 (-21)** | **4 (-1)** | **5 (3)** | **4 (0)** | **19 (-18)** | **5 (-1)** |
| BDEI | 37 (24) | 36 (24) | **5 (0)** | **6 (1)** | 45 (41) | 44 (39) | **6 (0)** | **6 (1)** | 36 (24) | 36 (22) | **5 (0)** | **6 (1)** | 45 (40) | 44 (39) | **6 (1)** | **6 (1)** |
| BDSS | 33 (-33) | 31 (-31) | 31 (-30) | 31 (-31) | **6 (0)** | **6 (0)** | **13 (-11)** | **6 (-1)** | 30 (-30) | 31 (-31) | 37 (-37) | 32 (-32) | **6 (0)** | **6 (0)** | **17 (-16)** | **7 (-1)** |
| BDEISS | 23 (-6) | 23 (-4) | 23 (9) | 25 (14) | 61 (56) | 61 (55) | **6 (0)** | **7 (1)** | 22 (-3) | 26 (-5) | 43 (36) | 36 (27) | 56 (49) | 60 (54) | **7 (0)** | **8 (0)** |
| BD-CT | 15 (-15) | 14 (-13) | 13 (-3) | 12 (-8) | 13 (-12) | 15 (-14) | 12 (-11) | 13 (-13) | **5 (0)** | **5 (0)** | **12 (-10)** | **5 (0)** | **6 (1)** | **5 (0)** | **12 (-12)** | **5 (-1)** |
| BDEI-CT | 21 (0) | 21 (2) | 12 (-6) | 14 (-2) | 22 (17) | 22 (16) | 11 (-6) | 14 (0) | 26 (8) | 27 (6) | **5 (0)** | **6 (1)** | 29 (22) | 28 (21) | **6 (-1)** | **6 (1)** |
| BDSS-CT | 32 (-32) | 29 (-28) | 22 (-2) | 24 (-22) | 12 (-5) | 12 (-5) | 15 (-1) | 12 (-4) | 22 (-20) | 22 (-20) | 23 (-17) | 20 (-15) | **6 (0)** | **7 (0)** | **11 (-9)** | **7 (-1)** |
| BDEISS-CT | 21 (-16) | 20 (-12) | 18 (2) | 20 (6) | 37 (30) | 38 (31) | 9 (-5) | 11 (-1) | 21 (-9) | 24 (-11) | 40 (36) | 35 (31) | 39 (30) | 39 (31) | **6 (0)** | **7 (0)** |
| 2000-5000-tip trees | | | | | | | | | | | | | | | | |
| BD | **3 (0)** | **3 (0)** | **11 (-10)** | **3 (0)** | **3 (1)** | **3 (0)** | **13 (-12)** | **3 (0)** | **3 (1)** | **3 (0)** | **16 (-15)** | **3 (0)** | **4 (2)** | **3 (0)** | **17 (-16)** | **3 (-1)** |
| BDEI | 37 (24) | 36 (24) | **4 (0)** | **4 (1)** | 44 (40) | 43 (38) | **4 (0)** | **4 (0)** | 36 (29) | 36 (23) | **4 (1)** | **4 (1)** | 43 (38) | 42 (37) | **5 (2)** | **4 (1)** |
| BDSS | 33 (-33) | 30 (-30) | 22 (-20) | 30 (-30) | **4 (0)** | **4 (0)** | **10 (-8)** | **5 (-1)** | 30 (-30) | 31 (-31) | 31 (-31) | 30 (-30) | **5 (0)** | **4 (-1)** | **13 (-12)** | **5 (-1)** |
| BDEISS | 23 (-6) | 23 (-2) | 26 (15) | 30 (21) | 62 (57) | 63 (58) | **4 (0)** | **5 (0)** | 21 (3) | 26 (-6) | 52 (45) | 45 (38) | 57 (51) | 62 (56) | **5 (0)** | **5 (1)** |
| BD-CT | 15 (-15) | 13 (-13) | 16 (12) | 13 (-4) | 13 (-12) | 14 (-13) | 7 (-4) | 14 (-13) | **3 (0)** | **3 (0)** | **9 (-7)** | **4 (0)** | **4 (0)** | **3 (0)** | **11 (-10)** | **4 (-1)** |
| BDEI-CT | 21 (-1) | 21 (2) | 12 (-2) | 14 (0) | 22 (17) | 22 (16) | 10 (-5) | 13 (0) | 25 (11) | 27 (7) | **3 (0)** | **4 (0)** | 28 (22) | 28 (21) | **5 (0)** | **4 (1)** |
| BDSS-CT | 32 (-32) | 29 (-28) | 36 (28) | 25 (-14) | 11 (-4) | 11 (-3) | 17 (7) | 10 (-4) | 22 (-19) | 23 (-22) | 19 (-12) | 17 (-13) | **5 (0)** | **5 (0)** | **8 (-6)** | **5 (-1)** |
| BDEISS-CT | 21 (-16) | 19 (-12) | 22 (11) | 22 (12) | 36 (30) | 39 (33) | 9 (-4) | 9 (-1) | 19 (-5) | 23 (-11) | 46 (42) | 42 (39) | 40 (31) | 39 (31) | **5 (0)** | **5 (1)** |

Mean absolute percentage errors, $100|d_{estimated} - d_{true}|/d_{true}$, and in parenthesis the corresponding biases, $100(d_{estimated} - d_{true})/d_{true}$, are reported for 1000 trees generated under each dataset for each estimator.
The errors and biases of the estimators corresponding to or generalizing the model that generated the data are shown in bold.

Table S5: Estimation errors for the incubation fraction $f_E$ for transmission trees generated under different models (rows) and different estimators (columns). The deep-learning (DL) estimator type – pure if the estimator was trained on the corresponding model, or mixed if it was trained on the corresponding and its nested models – is specified below its model.

| | BDEI | | | | BDEISS | | | | BDEI-CT | | | | BDEISS-CT | | | |
|---|---|---|---|---|---|---|---|---|---|---|---|---|---|---|---|---|
| | DL (pure) | | DL (mixed) | | DL (pure) | | DL (mixed) | | DL (pure) | | DL (mixed) | | DL (pure) | | DL (mixed) | |
| 200-500-tip trees | | | | | | | | | | | | | | | | |
| BD | **19** | **(19)** | **2** | **(2)** | **14** | **(14)** | **1** | **(1)** | **21** | **(21)** | **2** | **(2)** | **15** | **(15)** | **2** | **(2)** |
| BDSS | 49 | (49) | 31 | (31) | **8** | **(8)** | **1** | **(1)** | 57 | (57) | 42 | (42) | **11** | **(11)** | **1** | **(1)** |
| BD-CT | 10 | (10) | 0 | (0) | 8 | (8) | 0 | (0) | **15** | **(15)** | **1** | **(1)** | **11** | **(11)** | **1** | **(1)** |
| BDSS-CT | 45 | (45) | 25 | (25) | 6 | (6) | 1 | (1) | 59 | (59) | 46 | (46) | **10** | **(10)** | **1** | **(1)** |
| BDEI | **5** | **(0)** | **7** | **(-4)** | **6** | **(-3)** | **8** | **(-5)** | **5** | **(1)** | **7** | **(-4)** | **7** | **(-3)** | **8** | **(-5)** |
| BDEISS | 22 | (22) | 21 | (20) | **6** | **(0)** | **7** | **(-1)** | 27 | (27) | 26 | (26) | **6** | **(1)** | **7** | **(-1)** |
| BDEI-CT | 10 | (-8) | 16 | (-16) | 16 | (-15) | 23 | (-22) | **5** | **(0)** | **8** | **(-5)** | **9** | **(-7)** | **10** | **(-8)** |
| BDEISS-CT | 18 | (18) | 17 | (14) | 9 | (-5) | 13 | (-11) | 25 | (25) | 24 | (24) | **7** | **(1)** | **9** | **(0)** |
| 500-1000-tip trees | | | | | | | | | | | | | | | | |
| BD | **14** | **(14)** | **1** | **(1)** | **11** | **(11)** | **1** | **(1)** | **18** | **(18)** | **1** | **(1)** | **14** | **(14)** | **1** | **(1)** |
| BDSS | 43 | (43) | 25 | (25) | **4** | **(4)** | **0** | **(0)** | 56 | (56) | 38 | (38) | **7** | **(7)** | **1** | **(1)** |
| BD-CT | 4 | (4) | 0 | (0) | 4 | (4) | 0 | (0) | **11** | **(11)** | **1** | **(1)** | **9** | **(9)** | **1** | **(1)** |
| BDSS-CT | 26 | (26) | 17 | (17) | 3 | (3) | 0 | (0) | 55 | (55) | 46 | (46) | **7** | **(7)** | **1** | **(1)** |
| BDEI | **4** | **(0)** | **5** | **(-2)** | **5** | **(-2)** | **5** | **(-3)** | **4** | **(1)** | **5** | **(-2)** | **5** | **(-2)** | **5** | **(-3)** |
| BDEISS | 23 | (23) | 22 | (21) | **4** | **(0)** | **5** | **(-1)** | 28 | (28) | 27 | (27) | **5** | **(0)** | **5** | **(0)** |
| BDEI-CT | 10 | (-10) | 16 | (-15) | 16 | (-15) | 21 | (-21) | **4** | **(0)** | **5** | **(-3)** | **7** | **(-5)** | **6** | **(-4)** |
| BDEISS-CT | 17 | (16) | 18 | (14) | 9 | (-7) | 12 | (-11) | 26 | (26) | 26 | (25) | **5** | **(1)** | **6** | **(0)** |
| 1000-2000-tip trees | | | | | | | | | | | | | | | | |
| BD | **9** | **(9)** | **0** | **(0)** | **10** | **(10)** | **1** | **(1)** | **15** | **(15)** | **1** | **(1)** | **13** | **(13)** | **1** | **(1)** |
| BDSS | 31 | (31) | 19 | (19) | **2** | **(2)** | **0** | **(0)** | 54 | (54) | 34 | (34) | **4** | **(4)** | **0** | **(0)** |
| BD-CT | 2 | (2) | 0 | (0) | 3 | (3) | 0 | (0) | **8** | **(8)** | **0** | **(0)** | **8** | **(8)** | **1** | **(1)** |
| BDSS-CT | 12 | (12) | 9 | (9) | 2 | (2) | 0 | (0) | 50 | (50) | 34 | (34) | **5** | **(5)** | **0** | **(0)** |
| BDEI | **3** | **(0)** | **3** | **(-1)** | **3** | **(-1)** | **3** | **(-1)** | **3** | **(1)** | **3** | **(-1)** | **4** | **(-1)** | **4** | **(-1)** |
| BDEISS | 22 | (22) | 22 | (21) | **3** | **(0)** | **4** | **(-1)** | 28 | (28) | 27 | (27) | **3** | **(0)** | **4** | **(0)** |
| BDEI-CT | 11 | (-11) | 16 | (-16) | 16 | (-15) | 22 | (-22) | **3** | **(0)** | **3** | **(-1)** | **5** | **(-4)** | **5** | **(-3)** |
| BDEISS-CT | 16 | (15) | 17 | (13) | 9 | (-8) | 13 | (-13) | 26 | (26) | 26 | (25) | **4** | **(1)** | **5** | **(0)** |
| 2000-5000-tip trees | | | | | | | | | | | | | | | | |
| BD | **7** | **(7)** | **0** | **(0)** | **8** | **(8)** | **0** | **(0)** | **11** | **(11)** | **0** | **(0)** | **11** | **(11)** | **1** | **(1)** |
| BDSS | 22 | (22) | 14 | (14) | **2** | **(2)** | **0** | **(0)** | 49 | (49) | 24 | (24) | **3** | **(3)** | **0** | **(0)** |
| BD-CT | 1 | (1) | 0 | (0) | 1 | (1) | 1 | (1) | **6** | **(6)** | **0** | **(0)** | **7** | **(7)** | **1** | **(1)** |
| BDSS-CT | 5 | (5) | 4 | (4) | 1 | (1) | 0 | (0) | 44 | (44) | 24 | (24) | **4** | **(4)** | **0** | **(0)** |
| BDEI | **2** | **(0)** | **2** | **(-1)** | **2** | **(-1)** | **2** | **(-1)** | **2** | **(0)** | **2** | **(-1)** | **3** | **(-1)** | **3** | **(-1)** |
| BDEISS | 22 | (22) | 21 | (20) | **2** | **(0)** | **3** | **(0)** | 28 | (28) | 26 | (26) | **3** | **(0)** | **3** | **(0)** |
| BDEI-CT | 13 | (-13) | 18 | (-18) | 16 | (-16) | 24 | (-24) | **2** | **(0)** | **2** | **(-1)** | **4** | **(-2)** | **3** | **(-2)** |
| BDEISS-CT | 15 | (13) | 16 | (11) | 9 | (-9) | 15 | (-15) | 26 | (26) | 25 | (25) | **3** | **(0)** | **4** | **(0)** |

Mean absolute errors multiplied by 100, $100|f_{E\,estimated} - f_{E\,true}|$, and in parenthesis the corresponding biases, $100(f_{E\,estimated} - f_{E\,true})$, are reported for 1000 trees generated under each dataset for each estimator.
The errors and biases of the estimators corresponding to or generalizing the model that generated the data are shown in bold.
The upper group of rows contains data-generating models with $f_E = 0$, the bottom group of rows contains those with $f_E \geq 0$.

Table S6: Estimation errors for the superspreader fraction $f_S$ for transmission trees generated under different models (rows) and different estimators (columns). The deep-learning (DL) estimator type – pure if the estimator was trained on the corresponding model, or mixed if it was trained on the corresponding and its nested models – is specified below its model.

| | BDSS | | | | BDEISS | | | | BDSS-CT | | | | BDEISS-CT | | | |
|---|---|---|---|---|---|---|---|---|---|---|---|---|---|---|---|---|
| | DL (pure) | | DL (mixed) | | DL (pure) | | DL (mixed) | | DL (pure) | | DL (mixed) | | DL (pure) | | DL (mixed) | |
| 200-500-tip trees | | | | | | | | | | | | | | | | |
| BD | **22** | **(22)** | **1** | **(1)** | **18** | **(18)** | **1** | **(1)** | **24** | **(24)** | **1** | **(1)** | **20** | **(20)** | **1** | **(1)** |
| BDEI | 19 | (19) | 7 | (7) | **21** | **(21)** | **1** | **(1)** | 18 | (18) | 6 | (6) | **22** | **(22)** | **1** | **(1)** |
| BD-CT | 23 | (23) | 2 | (2) | 22 | (22) | 3 | (3) | **22** | **(22)** | **1** | **(1)** | **22** | **(22)** | **1** | **(1)** |
| BDEI-CT | 21 | (21) | 10 | (10) | 31 | (31) | 12 | (12) | 20 | (20) | 10 | (10) | **26** | **(26)** | **2** | **(2)** |
| BDSS | **5** | **(-1)** | **6** | **(-2)** | **7** | **(1)** | **7** | **(-2)** | **5** | **(0)** | **7** | **(-2)** | **7** | **(2)** | **7** | **(-2)** |
| BDEISS | 17 | (-17) | 17 | (-17) | **5** | **(-1)** | **6** | **(-2)** | 17 | (-17) | 18 | (-17) | **5** | **(-1)** | **7** | **(-4)** |
| BDSS-CT | 5 | (-3) | 6 | (-3) | 6 | (-3) | 6 | (-3) | **5** | **(-1)** | **5** | **(-2)** | **6** | **(0)** | **6** | **(-2)** |
| BDEISS-CT | 17 | (-17) | 17 | (-17) | 6 | (-4) | 8 | (-5) | 16 | (-16) | 17 | (-16) | **5** | **(-1)** | **8** | **(-5)** |
| 500-1000-tip trees | | | | | | | | | | | | | | | | |
| BD | **20** | **(20)** | **0** | **(0)** | **16** | **(16)** | **1** | **(1)** | **21** | **(21)** | **0** | **(0)** | **19** | **(19)** | **0** | **(0)** |
| BDEI | 17 | (17) | 7 | (7) | **20** | **(20)** | **0** | **(0)** | 16 | (16) | 6 | (6) | **21** | **(21)** | **0** | **(0)** |
| BD-CT | 22 | (22) | 2 | (2) | 21 | (21) | 3 | (3) | **19** | **(19)** | **0** | **(0)** | **20** | **(20)** | **0** | **(0)** |
| BDEI-CT | 20 | (20) | 10 | (10) | 33 | (33) | 15 | (15) | 19 | (19) | 10 | (10) | **24** | **(24)** | **2** | **(2)** |
| BDSS | **3** | **(0)** | **4** | **(-1)** | **5** | **(0)** | **4** | **(-1)** | **4** | **(0)** | **4** | **(-1)** | **5** | **(1)** | **4** | **(-1)** |
| BDEISS | 18 | (-17) | 18 | (-18) | **4** | **(-1)** | **4** | **(-1)** | 17 | (-17) | 18 | (-18) | **4** | **(-1)** | **5** | **(-2)** |
| BDSS-CT | 4 | (-3) | 4 | (-3) | 6 | (-4) | 4 | (-2) | **3** | **(-1)** | **4** | **(-1)** | **4** | **(-1)** | **4** | **(-1)** |
| BDEISS-CT | 17 | (-17) | 17 | (-17) | 6 | (-4) | 6 | (-5) | 16 | (-16) | 17 | (-17) | **4** | **(-1)** | **6** | **(-2)** |
| 1000-2000-tip trees | | | | | | | | | | | | | | | | |
| BD | **19** | **(19)** | **0** | **(0)** | **13** | **(13)** | **0** | **(0)** | **20** | **(20)** | **0** | **(0)** | **17** | **(17)** | **0** | **(0)** |
| BDEI | 16 | (16) | 7 | (7) | **19** | **(19)** | **0** | **(0)** | 16 | (16) | 7 | (7) | **22** | **(22)** | **0** | **(0)** |
| BD-CT | 22 | (22) | 1 | (1) | 20 | (20) | 3 | (3) | **18** | **(18)** | **0** | **(0)** | **18** | **(18)** | **0** | **(0)** |
| BDEI-CT | 20 | (20) | 11 | (11) | 34 | (34) | 19 | (19) | 18 | (18) | 10 | (10) | **24** | **(24)** | **1** | **(1)** |
| BDSS | **3** | **(0)** | **3** | **(0)** | **3** | **(0)** | **3** | **(0)** | **3** | **(0)** | **3** | **(0)** | **4** | **(1)** | **3** | **(0)** |
| BDEISS | 18 | (-17) | 18 | (-18) | **3** | **(0)** | **3** | **(-1)** | 18 | (-17) | 18 | (-18) | **3** | **(0)** | **3** | **(-1)** |
| BDSS-CT | 4 | (-3) | 4 | (-3) | 6 | (-6) | 4 | (-2) | **2** | **(0)** | **3** | **(-1)** | **3** | **(-1)** | **3** | **(-1)** |
| BDEISS-CT | 17 | (-17) | 17 | (-17) | 6 | (-5) | 7 | (-6) | 16 | (-16) | 17 | (-17) | **3** | **(0)** | **4** | **(-1)** |
| 2000-5000-tip trees | | | | | | | | | | | | | | | | |
| BD | **18** | **(18)** | **0** | **(0)** | **11** | **(11)** | **0** | **(0)** | **19** | **(19)** | **0** | **(0)** | **13** | **(13)** | **0** | **(0)** |
| BDEI | 14 | (14) | 6 | (6) | **19** | **(19)** | **0** | **(0)** | 15 | (15) | 6 | (6) | **20** | **(20)** | **0** | **(0)** |
| BD-CT | 25 | (25) | 1 | (1) | 20 | (20) | 7 | (7) | **18** | **(18)** | **0** | **(0)** | **16** | **(16)** | **0** | **(0)** |
| BDEI-CT | 20 | (20) | 11 | (11) | 36 | (36) | 21 | (21) | 18 | (18) | 10 | (10) | **22** | **(22)** | **1** | **(1)** |
| BDSS | **2** | **(0)** | **2** | **(0)** | **3** | **(0)** | **2** | **(0)** | **2** | **(0)** | **2** | **(0)** | **3** | **(0)** | **2** | **(0)** |
| BDEISS | 18 | (-18) | 18 | (-18) | **2** | **(0)** | **2** | **(0)** | 18 | (-18) | 18 | (-18) | **2** | **(0)** | **3** | **(0)** |
| BDSS-CT | 3 | (-3) | 4 | (-3) | 7 | (-6) | 4 | (-2) | **2** | **(0)** | **2** | **(0)** | **3** | **(-1)** | **2** | **(0)** |
| BDEISS-CT | 17 | (-17) | 17 | (-17) | 6 | (-5) | 7 | (-6) | 17 | (-16) | 17 | (-17) | **2** | **(0)** | **3** | **(-1)** |

Mean absolute errors multiplied by 100, $100|f_{S\,estimated} - f_{S\,true}|$, and in parenthesis the corresponding biases, $100(f_{S\,estimated} - f_{S\,true})$, are reported for 1000 trees generated under each dataset for each estimator.
The errors and biases of the estimators corresponding to or generalizing the model that generated the data are shown in bold.
The upper group of rows contains data-generating models with $f_S = 0$, the bottom group of rows contains those with $f_S \geq 0$.

Table S7: Estimation errors for the contact-tracing probability $\upsilon$ for transmission trees generated under different models (rows) and different estimators (columns). The deep-learning (DL) estimator type – pure if the estimator was trained on the corresponding model, or mixed if it was trained on the corresponding and its nested models – is specified below its model.

| | BD-CT | | BDEI-CT | | BDSS-CT | | BDEISS-CT | |
|---|---|---|---|---|---|---|---|---|
| | DL (pure) | DL (mixed) | DL (pure) | DL (mixed) | DL (pure) | DL (mixed) | DL (pure) | DL (mixed) |
| **200-500-tip trees** | | | | | | | | |
| BD | **11 ( 11)** | **0 ( 0)** | **14 ( 14)** | **1 ( 1)** | **11 ( 11)** | **0 ( 0)** | **12 ( 12)** | **0 ( 0)** |
| BDEI | 7 ( 7) | 0 ( 0) | **14 ( 14)** | **1 ( 1)** | 7 ( 7) | 0 ( 0) | **13 ( 13)** | **1 ( 1)** |
| BDSS | 23 ( 23) | 4 ( 4) | 49 ( 49) | 25 ( 25) | **13 ( 13)** | **1 ( 1)** | **13 ( 13)** | **1 ( 1)** |
| BDEISS | 18 ( 18) | 2 ( 2) | 47 ( 47) | 32 ( 32) | 15 ( 15) | 1 ( 1) | **18 ( 18)** | **3 ( 3)** |
| BD-CT | **7 ( 0)** | **8 ( -1)** | **8 ( 3)** | **8 ( -1)** | **7 ( -2)** | **8 ( -2)** | **8 ( -2)** | **8 ( -1)** |
| BDEI-CT | 21 (-20) | 24 (-22) | **7 ( 0)** | **8 ( -3)** | 22 (-21) | 25 (-24) | **10 ( -6)** | **10 ( -6)** |
| BDSS-CT | 12 ( 9) | 13 ( 7) | 24 ( 24) | 24 ( 23) | **7 ( 0)** | **9 ( -2)** | **8 ( 0)** | **9 ( -1)** |
| BDEISS-CT | 20 (-13) | 22 (-17) | 22 ( 22) | 21 ( 20) | 19 (-16) | 25 (-23) | **10 ( 1)** | **13 ( -2)** |
| **500-1000-tip trees** | | | | | | | | |
| BD | **9 ( 9)** | **0 ( 0)** | **12 ( 12)** | **0 ( 0)** | **10 ( 10)** | **0 ( 0)** | **11 ( 11)** | **0 ( 0)** |
| BDEI | 6 ( 6) | 0 ( 0) | **11 ( 11)** | **1 ( 1)** | 5 ( 5) | 0 ( 0) | **12 ( 12)** | **1 ( 1)** |
| BDSS | 26 ( 26) | 4 ( 4) | 59 ( 59) | 34 ( 34) | **10 ( 10)** | **0 ( 0)** | **10 ( 10)** | **0 ( 0)** |
| BDEISS | 21 ( 21) | 2 ( 2) | 52 ( 52) | 37 ( 37) | 13 ( 13) | 1 ( 1) | **16 ( 16)** | **2 ( 2)** |
| BD-CT | **5 ( 0)** | **5 ( -1)** | **6 ( 2)** | **5 ( -1)** | **5 ( -1)** | **5 ( -1)** | **6 ( -2)** | **6 ( 0)** |
| BDEI-CT | 21 (-20) | 23 (-22) | **5 ( 0)** | **6 ( -2)** | 23 (-22) | 24 (-23) | **8 ( -4)** | **8 ( -3)** |
| BDSS-CT | 12 ( 10) | 12 ( 10) | 26 ( 26) | 27 ( 27) | **6 ( 0)** | **6 ( -1)** | **7 ( 0)** | **6 ( -1)** |
| BDEISS-CT | 20 (-13) | 22 (-16) | 25 ( 25) | 24 ( 24) | 20 (-18) | 24 (-23) | **8 ( 1)** | **10 ( -1)** |
| **1000-2000-tip trees** | | | | | | | | |
| BD | **8 ( 8)** | **0 ( 0)** | **11 ( 11)** | **0 ( 0)** | **9 ( 9)** | **0 ( 0)** | **11 ( 11)** | **0 ( 0)** |
| BDEI | 5 ( 5) | 0 ( 0) | **10 ( 10)** | **0 ( 0)** | 4 ( 4) | 0 ( 0) | **10 ( 10)** | **0 ( 0)** |
| BDSS | 27 ( 27) | 5 ( 5) | 67 ( 67) | 42 ( 42) | **10 ( 10)** | **0 ( 0)** | **9 ( 9)** | **0 ( 0)** |
| BDEISS | 21 ( 21) | 2 ( 2) | 56 ( 56) | 46 ( 46) | 13 ( 13) | 0 ( 0) | **13 ( 13)** | **1 ( 1)** |
| BD-CT | **4 ( 0)** | **4 ( 0)** | **5 ( 1)** | **4 ( 0)** | **4 ( -1)** | **4 ( -1)** | **5 ( -2)** | **4 ( -1)** |
| BDEI-CT | 21 (-21) | 21 (-21) | **4 ( 0)** | **4 ( -1)** | 23 (-22) | 24 (-23) | **6 ( -3)** | **6 ( -2)** |
| BDSS-CT | 11 ( 10) | 13 ( 12) | 28 ( 28) | 29 ( 29) | **4 ( 0)** | **5 ( -1)** | **5 ( 0)** | **5 ( -1)** |
| BDEISS-CT | 20 (-14) | 20 (-14) | 26 ( 26) | 27 ( 27) | 21 (-20) | 23 (-22) | **6 ( 1)** | **8 ( 0)** |
| **2000-5000-tip trees** | | | | | | | | |
| BD | **8 ( 8)** | **0 ( 0)** | **12 ( 12)** | **0 ( 0)** | **9 ( 9)** | **0 ( 0)** | **12 ( 12)** | **0 ( 0)** |
| BDEI | 7 ( 7) | 0 ( 0) | **10 ( 10)** | **0 ( 0)** | 4 ( 4) | 0 ( 0) | **11 ( 11)** | **0 ( 0)** |
| BDSS | 31 ( 31) | 5 ( 5) | 75 ( 75) | 46 ( 46) | **9 ( 9)** | **0 ( 0)** | **8 ( 8)** | **0 ( 0)** |
| BDEISS | 24 ( 24) | 2 ( 2) | 63 ( 63) | 47 ( 47) | 12 ( 12) | 0 ( 0) | **12 ( 12)** | **1 ( 1)** |
| BD-CT | **3 ( 0)** | **3 ( 0)** | **3 ( 1)** | **3 ( 0)** | **3 ( 0)** | **3 ( 0)** | **4 ( -2)** | **3 ( 0)** |
| BDEI-CT | 21 (-20) | 22 (-21) | **3 ( 0)** | **3 ( 0)** | 23 (-22) | 24 (-23) | **4 ( -2)** | **4 ( -1)** |
| BDSS-CT | 12 ( 11) | 12 ( 10) | 27 ( 27) | 28 ( 28) | **3 ( 0)** | **3 ( 0)** | **4 ( -1)** | **4 ( 0)** |
| BDEISS-CT | 19 (-12) | 21 (-14) | 28 ( 28) | 28 ( 28) | 22 (-21) | 23 (-23) | **5 ( 1)** | **6 ( 0)** |

Mean absolute errors multiplied by 100, $100|\upsilon_{estimated} - \upsilon_{true}|$, and in parenthesis the corresponding biases, $100(\upsilon_{estimated} - \upsilon_{true})$, are reported for 1000 trees generated under each dataset for each estimator.
The errors and biases of the estimators corresponding to or generalizing the model that generated the data are shown in bold.
The upper group of rows contains data-generating models with $\upsilon = 0$, the bottom group of rows contains those with $\upsilon \geq 0$.

Table S8: Estimation errors for the superspreading transmission increase $X_S$ for transmission trees generated under different models (rows) and different estimators (columns). The deep-learning (DL) estimator type – pure if the estimator was trained on the corresponding model, or mixed if it was trained on the corresponding and its nested models – is specified below its model.

| | BDSS | | BDEISS | | BDSS-CT | | BDEISS-CT | |
|---|---|---|---|---|---|---|---|---|
| | DL (pure) | DL (mixed) | DL (pure) | DL (mixed) | DL (pure) | DL (mixed) | DL (pure) | DL (mixed) |
| **200-500-tip trees** | | | | | | | | |
| BDSS | **28 ( 2)** | **30 ( 0)** | **31 ( -7)** | **30 ( -1)** | **29 ( 3)** | **30 ( 0)** | **32 ( -2)** | **32 ( -2)** |
| BDEISS | 47 ( 33) | 48 ( 32) | **25 ( 3)** | **27 ( 4)** | 51 ( 36) | 48 ( 32) | **27 ( 3)** | **28 ( 1)** |
| BDSS-CT | 24 ( -1) | 25 ( 0) | 27 ( 0) | 26 ( 4) | **24 ( 4)** | **26 ( 3)** | **27 ( 6)** | **27 ( 2)** |
| BDEISS-CT | 53 ( 38) | 52 ( 37) | 31 ( 14) | 33 ( 15) | 54 ( 41) | 53 ( 38) | **26 ( 3)** | **29 ( -1)** |
| **500-1000-tip trees** | | | | | | | | |
| BDSS | **22 ( 1)** | **23 ( 1)** | **24 (-10)** | **23 ( 0)** | **24 ( 4)** | **24 ( 1)** | **26 ( -5)** | **24 ( -1)** |
| BDEISS | 52 ( 38) | 51 ( 38) | **21 ( 4)** | **22 ( 4)** | 53 ( 41) | 51 ( 38) | **22 ( 3)** | **23 ( 3)** |
| BDSS-CT | 20 ( -5) | 21 ( -5) | 23 ( -4) | 22 ( -1) | **20 ( 3)** | **21 ( 2)** | **23 ( 3)** | **22 ( 2)** |
| BDEISS-CT | 58 ( 44) | 57 ( 42) | 28 ( 13) | 30 ( 15) | 58 ( 46) | 58 ( 45) | **21 ( 2)** | **24 ( 1)** |
| **1000-2000-tip trees** | | | | | | | | |
| BDSS | **19 ( 2)** | **19 ( 2)** | **22 (-11)** | **20 ( 1)** | **20 ( 3)** | **20 ( 2)** | **22 ( -8)** | **21 ( 0)** |
| BDEISS | 55 ( 44) | 56 ( 45) | **16 ( 2)** | **18 ( 4)** | 55 ( 43) | 52 ( 41) | **18 ( 4)** | **18 ( 4)** |
| BDSS-CT | 18 ( -7) | 18 ( -6) | 21 ( -7) | 19 ( -5) | **15 ( 2)** | **16 ( 1)** | **18 ( 1)** | **17 ( 2)** |
| BDEISS-CT | 60 ( 47) | 59 ( 47) | 25 ( 12) | 28 ( 14) | 61 ( 50) | 58 ( 47) | **17 ( 2)** | **20 ( 1)** |
| **2000-5000-tip trees** | | | | | | | | |
| BDSS | **14 ( 0)** | **14 ( 0)** | **18 (-11)** | **15 ( -1)** | **15 ( 1)** | **15 ( 0)** | **19 (-10)** | **16 ( -1)** |
| BDEISS | 59 ( 48) | 62 ( 52) | **12 ( 2)** | **14 ( 4)** | 57 ( 46) | 58 ( 47) | **14 ( 3)** | **15 ( 3)** |
| BDSS-CT | 16 ( -9) | 16 ( -7) | 21 ( -8) | 18 ( -6) | **12 ( 2)** | **12 ( 2)** | **15 ( 1)** | **13 ( 2)** |
| BDEISS-CT | 63 ( 49) | 64 ( 52) | 24 ( 12) | 27 ( 14) | 63 ( 53) | 63 ( 53) | **13 ( 2)** | **15 ( 0)** |

Mean absolute percentage errors, $100|X_{S_{estimated}} - X_{S_{true}}|/X_{S_{true}}$, and in parenthesis the corresponding biases, $100(X_{S_{estimated}} - X_{S_{true}})/X_{S_{true}}$, are reported for 1000 trees generated under each dataset for each estimator.
The errors and biases of the estimators corresponding to or generalizing the model that generated the data are shown in bold.

Table S9: Estimation errors for the contact-traced removal speed up $X_C$ for transmission trees generated under different models (rows) and different estimators (columns). The deep-learning (DL) estimator type – pure if the estimator was trained on the corresponding model, or mixed if it was trained on the corresponding and its nested models – is specified below its model.

| | BD-CT | | BDEI-CT | | BDSS-CT | | BDEISS-CT | |
|---|---|---|---|---|---|---|---|---|
| | DL (pure) | DL (mixed) | DL (pure) | DL (mixed) | DL (pure) | DL (mixed) | DL (pure) | DL (mixed) |
| 200-500-tip trees | | | | | | | | |
| BD-CT | **27 ( 7)** | **29 ( 1)** | **32 ( 12)** | **30 ( 3)** | **28 ( 10)** | **30 ( 2)** | **34 ( 16)** | **32 ( 4)** |
| BDEI-CT | 45 ( 12) | 46 (-16) | **36 ( 8)** | **40 ( 0)** | 48 ( 19) | 48 (-14) | **37 ( 10)** | **41 ( -2)** |
| BDSS-CT | 31 ( -6) | 33 ( -8) | 55 ( 36) | 53 ( 31) | **30 ( 6)** | **32 ( 0)** | **37 ( 14)** | **35 ( 4)** |
| BDEISS-CT | 45 ( -5) | 49 (-32) | 49 ( 26) | 51 ( 25) | 45 ( 3) | 54 (-37) | **39 ( 10)** | **45 ( -7)** |
| 500-1000-tip trees | | | | | | | | |
| BD-CT | **21 ( 4)** | **22 ( 2)** | **25 ( 7)** | **23 ( 2)** | **22 ( 7)** | **23 ( 2)** | **26 ( 10)** | **24 ( 4)** |
| BDEI-CT | 46 ( 22) | 42 ( -7) | **28 ( 5)** | **31 ( 0)** | 49 ( 28) | 46 ( -2) | **29 ( 6)** | **32 ( -1)** |
| BDSS-CT | 27 ( -7) | 28 ( -8) | 52 ( 36) | 53 ( 36) | **22 ( 3)** | **24 ( -1)** | **28 ( 7)** | **26 ( 1)** |
| BDEISS-CT | 48 ( 7) | 44 (-30) | 43 ( 22) | 44 ( 23) | 47 ( 14) | 47 (-29) | **30 ( 4)** | **37 ( -6)** |
| 1000-2000-tip trees | | | | | | | | |
| BD-CT | **16 ( 4)** | **17 ( 2)** | **19 ( 2)** | **18 ( 2)** | **16 ( 5)** | **18 ( 2)** | **20 ( 5)** | **19 ( 3)** |
| BDEI-CT | 54 ( 36) | 44 ( 10) | **21 ( 2)** | **24 ( 1)** | 58 ( 41) | 53 ( 22) | **23 ( 4)** | **26 ( 1)** |
| BDSS-CT | 25 (-10) | 27 ( -8) | 51 ( 36) | 56 ( 41) | **17 ( 1)** | **19 ( -2)** | **22 ( 2)** | **20 ( -1)** |
| BDEISS-CT | 51 ( 18) | 44 (-20) | 40 ( 21) | 40 ( 22) | 49 ( 23) | 44 (-10) | **22 ( 1)** | **27 ( -6)** |
| 2000-5000-tip trees | | | | | | | | |
| BD-CT | **10 ( 2)** | **11 ( 1)** | **13 ( 0)** | **12 ( 0)** | **10 ( 1)** | **12 ( 1)** | **14 ( 0)** | **12 ( 0)** |
| BDEI-CT | 55 ( 40) | 47 ( 21) | **16 ( 3)** | **18 ( 1)** | 66 ( 53) | 63 ( 37) | **17 ( 3)** | **19 ( 1)** |
| BDSS-CT | 23 ( -9) | 23 (-12) | 56 ( 42) | 49 ( 34) | **12 ( 1)** | **13 ( 0)** | **16 ( -1)** | **14 ( -2)** |
| BDEISS-CT | 57 ( 24) | 46 ( -1) | 35 ( 18) | 37 ( 22) | 65 ( 45) | 53 ( 12) | **19 ( 1)** | **21 ( -5)** |

Mean absolute percentage errors, $100|X_{C\,estimated} - X_{C\,true}|/X_{C\,true}$, and in parenthesis the corresponding biases, $100(X_{C\,estimated} - X_{C\,true})/X_{C\,true}$, are reported for 1000 trees generated under each dataset for each estimator.
The errors and biases of the estimators corresponding to or generalizing the model that generated the data are shown in bold.

Table S10: CI coverage for the average reproduction number $R_e$ for transmission trees generated under different models (rows) and different estimators (columns). The estimator type – maximum-likelihood (ML) or deep-learning-based (DL); pure if the estimator was trained on the corresponding model, or mixed if it was trained on the corresponding and its nested models – is specified below its model.

| | BD | | BDEI | | BDSS | | BDEISS | | BD-CT | | BDEI-CT | | BDSS-CT | | BDEISS-CT | |
|---|---|---|---|---|---|---|---|---|---|---|---|---|---|---|---|---|
| | ML | DL (pure) | DL (pure) | DL (mixed) | DL (pure) | DL (mixed) | DL (pure) | DL (mixed) | DL (pure) | DL (mixed) | DL (pure) | DL (mixed) | DL (pure) | DL (mixed) | DL (pure) | DL (mixed) |
| 200-500-tip trees | | | | | | | | | | | | | | | | |
| BD | **100 (78)** | **94 (44)** | **93 (44)** | **94 (45)** | **95 (51)** | **96 (46)** | **97 (51)** | **96 (47)** | **94 (48)** | **95 (45)** | **95 (50)** | **95 (46)** | **93 (54)** | **96 (48)** | **98 (57)** | **96 (49)** |
| BDEI | 77 (65) | 64 (39) | **96 (52)** | **96 (54)** | 88 (53) | 87 (50) | **96 (57)** | **97 (55)** | 68 (44) | 66 (41) | **95 (55)** | **96 (55)** | 86 (55) | 88 (51) | **95 (59)** | **96 (56)** |
| BDSS | 58 (51) | 41 (35) | 72 (47) | 65 (44) | **95 (55)** | **94 (56)** | **88 (55)** | **94 (56)** | 52 (41) | 44 (38) | 84 (54) | 78 (52) | **95 (59)** | **95 (57)** | **90 (58)** | **94 (58)** |
| BDEISS | 52 (49) | 43 (35) | 86 (59) | 85 (62) | 84 (58) | 84 (60) | **95 (62)** | **96 (65)** | 49 (40) | 45 (36) | 89 (69) | 90 (71) | 85 (60) | 86 (61) | **96 (65)** | **96 (67)** |
| BD-CT | 0 ( 0) | 72 (37) | 74 (41) | 82 (42) | 83 (46) | 78 (42) | 84 (47) | 82 (45) | **96 (54)** | **97 (55)** | **97 (57)** | **96 (56)** | **96 (61)** | **96 (58)** | **97 (62)** | **96 (59)** |
| BDEI-CT | 70 (53) | 58 (34) | 84 (47) | 87 (48) | 87 (51) | 86 (49) | 88 (51) | 89 (51) | 69 (43) | 67 (42) | **95 (56)** | **95 (59)** | 90 (57) | 90 (56) | **95 (60)** | **96 (60)** |
| BDSS-CT | 48 (44) | 34 (31) | 64 (45) | 58 (41) | 85 (52) | 86 (55) | 84 (56) | 89 (58) | 65 (49) | 64 (49) | 85 (55) | 86 (59) | **95 (64)** | **95 (67)** | **94 (64)** | **96 (67)** |
| BDEISS-CT | 49 (44) | 33 (31) | 82 (61) | 80 (64) | 88 (61) | 88 (63) | 91 (62) | 93 (66) | 55 (41) | 49 (38) | 89 (72) | 91 (76) | 89 (64) | 90 (66) | **95 (68)** | **95 (72)** |
| 500-1000-tip trees | | | | | | | | | | | | | | | | |
| BD | **100 (51)** | **95 (31)** | **95 (35)** | **95 (32)** | **95 (36)** | **95 (33)** | **96 (39)** | **96 (34)** | **94 (34)** | **95 (32)** | **96 (37)** | **95 (33)** | **94 (38)** | **96 (34)** | **98 (43)** | **96 (35)** |
| BDEI | 66 (43) | 55 (27) | **96 (37)** | **96 (39)** | 80 (36) | 82 (37) | **96 (42)** | **96 (41)** | 55 (30) | 57 (30) | **96 (41)** | **96 (40)** | 80 (36) | 82 (36) | **95 (44)** | **96 (41)** |
| BDSS | 35 (33) | 25 (25) | 58 (35) | 46 (32) | **95 (40)** | **95 (41)** | **91 (42)** | **95 (43)** | 33 (29) | 26 (27) | 78 (40) | 65 (39) | **94 (43)** | **95 (42)** | **91 (45)** | **95 (43)** |
| BDEISS | 38 (33) | 32 (25) | 76 (43) | 77 (49) | 75 (43) | 77 (46) | **96 (46)** | **96 (50)** | 38 (28) | 36 (27) | 75 (61) | 78 (62) | 80 (45) | 77 (47) | **96 (50)** | **96 (51)** |
| BD-CT | 0 ( 0) | 60 (25) | 67 (34) | 68 (29) | 72 (32) | 63 (28) | 86 (37) | 70 (30) | **96 (39)** | **96 (40)** | **96 (43)** | **96 (41)** | **97 (43)** | **96 (41)** | **96 (45)** | **96 (41)** |
| BDEI-CT | 60 (35) | 49 (23) | 74 (33) | 82 (35) | 77 (35) | 79 (36) | 83 (36) | 84 (37) | 63 (31) | 64 (32) | **95 (41)** | **95 (43)** | 82 (38) | 86 (39) | **95 (43)** | **95 (44)** |
| BDSS-CT | 28 (29) | 22 (22) | 60 (45) | 38 (30) | 79 (37) | 81 (39) | 78 (40) | 84 (42) | 52 (36) | 55 (38) | 85 (44) | 81 (43) | **95 (47)** | **95 (49)** | **93 (48)** | **95 (50)** |
| BDEISS-CT | 28 (29) | 20 (21) | 68 (45) | 72 (51) | 84 (42) | 87 (46) | 87 (43) | 91 (47) | 42 (30) | 40 (31) | 76 (60) | 82 (63) | 85 (45) | 88 (50) | **96 (49)** | **96 (54)** |
| 1000-2000-tip trees | | | | | | | | | | | | | | | | |
| BD | **100 (36)** | **96 (23)** | **94 (26)** | **96 (24)** | **97 (27)** | **96 (24)** | **98 (31)** | **96 (25)** | **96 (26)** | **96 (24)** | **98 (30)** | **96 (24)** | **95 (28)** | **96 (25)** | **98 (34)** | **97 (26)** |
| BDEI | 60 (31) | 52 (20) | **96 (28)** | **95 (30)** | 79 (28) | 79 (30) | **95 (32)** | **96 (30)** | 48 (25) | 51 (23) | **96 (31)** | **96 (30)** | 72 (27) | 75 (27) | **96 (35)** | **97 (31)** |
| BDSS | 21 (24) | 19 (19) | 56 (29) | 35 (23) | **96 (31)** | **96 (31)** | **90 (33)** | **95 (32)** | 23 (22) | 18 (21) | 74 (34) | 56 (32) | **95 (33)** | **95 (32)** | **91 (36)** | **96 (33)** |
| BDEISS | 28 (23) | 27 (20) | 64 (34) | 65 (42) | 65 (33) | 67 (37) | **96 (35)** | **96 (39)** | 28 (24) | 26 (22) | 54 (55) | 60 (55) | 72 (36) | 67 (36) | **97 (39)** | **96 (39)** |
| BD-CT | 0 ( 0) | 49 (18) | 54 (32) | 58 (24) | 52 (22) | 44 (20) | 73 (27) | 58 (22) | **96 (29)** | **96 (30)** | **97 (34)** | **97 (30)** | **96 (31)** | **96 (30)** | **97 (34)** | **97 (31)** |
| BDEI-CT | 49 (25) | 44 (18) | 65 (26) | 69 (25) | 72 (26) | 73 (27) | 71 (27) | 74 (28) | 56 (24) | 58 (23) | **95 (30)** | **96 (32)** | 78 (28) | 80 (30) | **95 (32)** | **96 (33)** |
| BDSS-CT | 15 (20) | 22 (19) | 52 (41) | 40 (27) | 70 (27) | 75 (29) | 67 (30) | 78 (31) | 48 (30) | 51 (32) | 84 (37) | 82 (38) | **96 (35)** | **97 (38)** | **96 (36)** | **96 (38)** |
| BDEISS-CT | 16 (21) | 20 (19) | 61 (34) | 60 (39) | 80 (33) | 80 (35) | 79 (31) | 85 (35) | 31 (23) | 31 (24) | 61 (51) | 64 (53) | 82 (35) | 82 (37) | **96 (37)** | **97 (41)** |
| 2000-5000-tip trees | | | | | | | | | | | | | | | | |
| BD | **100 (24)** | **97 (16)** | **94 (19)** | **97 (17)** | **96 (19)** | **97 (17)** | **97 (23)** | **97 (17)** | **98 (20)** | **97 (17)** | **97 (22)** | **97 (17)** | **94 (21)** | **97 (17)** | **99 (27)** | **97 (18)** |
| BDEI | 48 (21) | 39 (14) | **96 (20)** | **96 (21)** | 64 (21) | 60 (20) | **95 (22)** | **96 (22)** | 45 (18) | 40 (17) | **96 (22)** | **96 (22)** | 55 (18) | 66 (21) | **96 (26)** | **97 (23)** |
| BDSS | 11 (16) | 14 (14) | 51 (18) | 16 (16) | **95 (22)** | **96 (23)** | **89 (25)** | **96 (23)** | 18 (19) | 11 (15) | 66 (27) | 42 (22) | **95 (25)** | **95 (23)** | **89 (29)** | **95 (24)** |
| BDEISS | 20 (16) | 22 (14) | 45 (29) | 49 (39) | 50 (25) | 45 (28) | **96 (26)** | **96 (29)** | 28 (19) | 19 (16) | 35 (44) | 43 (47) | 55 (28) | 52 (30) | **97 (29)** | **96 (29)** |
| BD-CT | 38 (18) | 36 (13) | 41 (23) | 48 (21) | 42 (16) | 36 (15) | 83 (26) | 57 (18) | **97 (20)** | **97 (21)** | **98 (26)** | **97 (22)** | **97 (22)** | **97 (21)** | **98 (27)** | **97 (21)** |
| BDEI-CT | 38 (17) | 36 (13) | 57 (22) | 59 (20) | 58 (19) | 62 (19) | 64 (22) | 71 (21) | 52 (17) | 56 (19) | **96 (22)** | **97 (24)** | 71 (20) | 76 (23) | **96 (24)** | **96 (24)** |
| BDSS-CT | 7 (14) | 16 (14) | 23 (27) | 27 (22) | 62 (20) | 62 (21) | 65 (28) | 72 (24) | 45 (25) | 38 (23) | 85 (36) | 74 (29) | **96 (25)** | **96 (27)** | **96 (29)** | **96 (28)** |
| BDEISS-CT | 7 (14) | 17 (14) | 49 (31) | 54 (38) | 67 (23) | 64 (25) | 70 (23) | 78 (26) | 30 (18) | 25 (20) | 45 (40) | 50 (43) | 69 (27) | 77 (30) | **95 (27)** | **97 (31)** |

CI coverage (percentage of trees for which the real parameter value was within the estimated CI), and in parenthesis the corresponding mean relative CI width, $100(R_{e\,97.5\%} - R_{e\,2.5\%})/R_{e\,true}$, are reported for 1000 trees generated under each dataset for each estimator.
The values of the estimators corresponding to or generalizing the model that generated the data are shown in bold.

Table S11: CI coverage for the average infection time $d$ for transmission trees generated under different models (rows) and different estimators (columns). The estimator type – maximum-likelihood (ML) or deep-learning-based (DL); pure if the estimator was trained on the corresponding model, or mixed if it was trained on the corresponding and its nested models – is specified below its model.

| | BD | | BDEI | | BDSS | | BDEISS | | BD-CT | | BDEI-CT | | BDSS-CT | | BDEISS-CT | |
|---|---|---|---|---|---|---|---|---|---|---|---|---|---|---|---|---|
| | ML | DL (pure) | DL (pure) | DL (mixed) | DL (pure) | DL (mixed) | DL (pure) | DL (mixed) | DL (pure) | DL (mixed) | DL (pure) | DL (mixed) | DL (pure) | DL (mixed) | DL (pure) | DL (mixed) |
| 200-500-tip trees | | | | | | | | | | | | | | | | |
| BD | **99 (58)** | **95 (39)** | **51 (42)** | **96 (50)** | **95 (49)** | **96 (42)** | **72 (51)** | **97 (55)** | **95 (43)** | **96 (40)** | **44 (39)** | **97 (53)** | **93 (52)** | **96 (44)** | **70 (49)** | **98 (58)** |
| BDEI | 40 (71) | 35 (50) | **96 (49)** | **95 (62)** | 32 (71) | 33 (67) | **96 (56)** | **94 (68)** | 37 (55) | 36 (52) | **95 (51)** | **95 (65)** | 29 (74) | 35 (69) | **94 (58)** | **95 (68)** |
| BDSS | 24 (34) | 18 (29) | 9 (29) | 17 (34) | **94 (52)** | **94 (53)** | **74 (56)** | **94 (59)** | 28 (35) | 21 (31) | 15 (30) | 28 (40) | **95 (55)** | **94 (53)** | **63 (52)** | **94 (62)** |
| BDEISS | 59 (49) | 53 (42) | 70 (61) | 78 (66) | 28 (77) | 29 (78) | **96 (57)** | **96 (71)** | 61 (49) | 51 (43) | 62 (79) | 74 (82) | 31 (80) | 32 (82) | **96 (62)** | **96 (72)** |
| BD-CT | 0 ( 0) | 66 (33) | 57 (42) | 74 (36) | 81 (44) | 72 (38) | 72 (50) | 71 (40) | **96 (49)** | **96 (50)** | **73 (47)** | **97 (55)** | **96 (58)** | **96 (53)** | **88 (57)** | **98 (60)** |
| BDEI-CT | 60 (48) | 53 (40) | 84 (44) | 86 (52) | 57 (63) | 60 (60) | 87 (53) | 88 (67) | 51 (51) | 50 (50) | **95 (50)** | **94 (61)** | 53 (70) | 57 (68) | **94 (56)** | **95 (67)** |
| BDSS-CT | 22 (28) | 24 (27) | 30 (37) | 27 (32) | 79 (47) | 79 (49) | 74 (56) | 78 (51) | 55 (43) | 53 (42) | 44 (39) | 60 (50) | **95 (59)** | **95 (61)** | **82 (57)** | **96 (65)** |
| BDEISS-CT | 56 (38) | 52 (35) | 71 (60) | 78 (63) | 42 (69) | 42 (70) | 90 (54) | 92 (72) | 62 (46) | 58 (43) | 64 (79) | 72 (83) | 49 (77) | 50 (79) | **95 (59)** | **96 (72)** |
| 500-1000-tip trees | | | | | | | | | | | | | | | | |
| BD | **99 (39)** | **95 (28)** | **60 (40)** | **94 (35)** | **95 (34)** | **96 (29)** | **74 (45)** | **97 (39)** | **94 (30)** | **95 (28)** | **41 (34)** | **96 (37)** | **93 (37)** | **96 (30)** | **65 (42)** | **97 (42)** |
| BDEI | 28 (47) | 23 (35) | **96 (35)** | **96 (43)** | 25 (50) | 24 (48) | **96 (41)** | **95 (46)** | 28 (40) | 25 (37) | **96 (38)** | **95 (44)** | 18 (50) | 25 (49) | **95 (43)** | **95 (46)** |
| BDSS | 8 (22) | 6 (20) | 8 (28) | 8 (27) | **95 (38)** | **95 (39)** | **83 (50)** | **95 (43)** | 12 (24) | 7 (22) | 11 (23) | 17 (33) | **94 (40)** | **95 (40)** | **70 (47)** | **95 (45)** |
| BDEISS | 43 (33) | 38 (30) | 53 (46) | 63 (55) | 20 (58) | 21 (60) | **96 (42)** | **96 (52)** | 44 (35) | 40 (33) | 40 (68) | 54 (71) | 24 (62) | 22 (63) | **96 (46)** | **95 (52)** |
| BD-CT | 0 ( 0) | 48 (22) | 66 (37) | 60 (26) | 61 (30) | 56 (26) | 86 (45) | 55 (26) | **95 (35)** | **96 (36)** | **76 (39)** | **96 (39)** | **96 (41)** | **96 (37)** | **86 (46)** | **96 (43)** |
| BDEI-CT | 44 (32) | 37 (27) | 74 (33) | 74 (40) | 50 (43) | 49 (42) | 78 (39) | 82 (50) | 37 (36) | 36 (37) | **96 (36)** | **94 (44)** | 41 (48) | 42 (49) | **96 (43)** | **95 (47)** |
| BDSS-CT | 10 (19) | 13 (19) | 46 (42) | 24 (28) | 69 (34) | 70 (35) | 75 (48) | 71 (38) | 40 (31) | 41 (33) | 31 (28) | 45 (37) | **95 (44)** | **95 (46)** | **86 (48)** | **95 (49)** |
| BDEISS-CT | 42 (25) | 39 (23) | 58 (45) | 66 (54) | 30 (50) | 34 (54) | 85 (38) | 90 (51) | 48 (33) | 46 (34) | 42 (64) | 54 (68) | 36 (56) | 38 (60) | **97 (44)** | **96 (53)** |
| 1000-2000-tip trees | | | | | | | | | | | | | | | | |
| BD | **99 (27)** | **96 (20)** | **70 (37)** | **97 (24)** | **96 (24)** | **96 (21)** | **69 (37)** | **96 (26)** | **96 (23)** | **96 (21)** | **50 (32)** | **96 (25)** | **95 (28)** | **97 (22)** | **55 (34)** | **97 (29)** |
| BDEI | 20 (34) | 19 (27) | **96 (26)** | **95 (31)** | 17 (37) | 18 (36) | **95 (31)** | **96 (32)** | 22 (31) | 19 (28) | **95 (29)** | **95 (31)** | 14 (38) | 16 (35) | **95 (34)** | **94 (33)** |
| BDSS | 2 (16) | 3 (16) | 24 (31) | 6 (20) | **95 (29)** | **95 (30)** | **85 (42)** | **96 (33)** | 10 (20) | 4 (17) | 9 (22) | 12 (25) | **94 (32)** | **95 (31)** | **72 (40)** | **96 (34)** |
| BDEISS | 32 (23) | 30 (24) | 42 (37) | 45 (45) | 15 (44) | 16 (49) | **96 (32)** | **96 (39)** | 42 (30) | 30 (24) | 28 (61) | 38 (60) | 17 (50) | 17 (51) | **96 (35)** | **95 (40)** |
| BD-CT | 0 ( 0) | 39 (17) | 64 (31) | 61 (25) | 46 (21) | 34 (18) | 84 (40) | 39 (18) | **96 (26)** | **96 (27)** | **82 (34)** | **96 (29)** | **96 (30)** | **96 (28)** | **82 (36)** | **96 (31)** |
| BDEI-CT | 33 (22) | 29 (21) | 67 (27) | 64 (31) | 38 (31) | 40 (31) | 69 (30) | 70 (36) | 30 (28) | 29 (28) | **96 (27)** | **96 (32)** | 30 (36) | 32 (36) | **95 (32)** | **96 (35)** |
| BDSS-CT | 5 (13) | 10 (16) | 53 (41) | 34 (30) | 58 (25) | 61 (26) | 70 (41) | 65 (29) | 37 (26) | 36 (27) | 37 (29) | 49 (35) | **95 (33)** | **96 (36)** | **88 (39)** | **96 (38)** |
| BDEISS-CT | 33 (18) | 36 (21) | 52 (36) | 53 (42) | 21 (38) | 24 (40) | 76 (29) | 84 (38) | 46 (28) | 36 (26) | 30 (56) | 36 (57) | 27 (44) | 30 (47) | **97 (33)** | **98 (41)** |
| 2000-5000-tip trees | | | | | | | | | | | | | | | | |
| BD | **99 (18)** | **97 (14)** | **77 (32)** | **96 (16)** | **96 (18)** | **97 (15)** | **67 (30)** | **97 (18)** | **97 (17)** | **97 (15)** | **59 (29)** | **97 (17)** | **95 (20)** | **97 (15)** | **50 (29)** | **97 (20)** |
| BDEI | 15 (23) | 15 (19) | **96 (19)** | **96 (21)** | 14 (25) | 17 (25) | **95 (22)** | **97 (22)** | 16 (25) | 14 (20) | **97 (22)** | **95 (21)** | 10 (27) | 14 (28) | **93 (26)** | **96 (23)** |
| BDSS | 0 (11) | 1 (10) | 37 (28) | 2 (13) | **96 (21)** | **95 (22)** | **85 (34)** | **96 (24)** | 5 (16) | 1 (12) | 16 (23) | 8 (18) | **96 (24)** | **95 (22)** | **76 (35)** | **96 (26)** |
| BDEISS | 22 (16) | 21 (15) | 27 (31) | 27 (38) | 11 (34) | 12 (38) | **96 (23)** | **97 (28)** | 37 (26) | 20 (19) | 24 (54) | 26 (53) | 12 (40) | 14 (39) | **98 (27)** | **96 (29)** |
| BD-CT | 20 (13) | 25 (11) | 60 (39) | 38 (17) | 32 (15) | 24 (13) | 94 (37) | 29 (16) | **97 (18)** | **97 (19)** | **86 (28)** | **97 (20)** | **97 (22)** | **97 (19)** | **80 (29)** | **98 (22)** |
| BDEI-CT | 24 (15) | 20 (14) | 54 (23) | 45 (22) | 28 (21) | 32 (23) | 62 (23) | 65 (30) | 23 (21) | 22 (21) | **98 (20)** | **98 (23)** | 21 (27) | 24 (27) | **96 (25)** | **97 (26)** |
| BDSS-CT | 2 ( 9) | 6 ( 9) | 41 (55) | 23 (23) | 49 (18) | 50 (19) | 69 (38) | 57 (21) | 28 (21) | 25 (22) | 41 (27) | 49 (26) | **97 (24)** | **95 (26)** | **90 (33)** | **97 (28)** |
| BDEISS-CT | 24 (12) | 25 (13) | 38 (30) | 42 (35) | 16 (27) | 18 (31) | 64 (21) | 77 (27) | 35 (21) | 30 (22) | 22 (47) | 25 (48) | 20 (34) | 23 (36) | **96 (25)** | **97 (31)** |

CI coverage (percentage of trees for which the real parameter value was within the estimated CI), and in parenthesis the corresponding mean relative CI width, $100(d_{97.5\%} - d_{2.5\%})/d_{true}$, are reported for 1000 trees generated under each dataset for each estimator.
The values of the estimators corresponding to or generalizing the model that generated the data are shown in bold.

Table S12: CI coverage for the incubation fraction $f_E$ for transmission trees generated under different models (rows) and different estimators (columns). The deep-learning (DL) estimator type – pure if the estimator was trained on the corresponding model, or mixed if it was trained on the corresponding and its nested models – is specified below its model.

| | BDEI | | BDEISS | | BDEI-CT | | BDEISS-CT | |
|---|---|---|---|---|---|---|---|---|
| | DL (pure) | DL (mixed) | DL (pure) | DL (mixed) | DL (pure) | DL (mixed) | DL (pure) | DL (mixed) |
| 200-500-tip trees | | | | | | | | |
| BD | **61 (34)** | **99 (22)** | **66 (32)** | **99 (23)** | **39 (33)** | **99 (24)** | **62 (34)** | **99 (24)** |
| BDSS | 10 (36) | 44 (39) | **87 (28)** | **99 (14)** | 1 (30) | 36 (42) | **82 (35)** | **99 (20)** |
| BD-CT | 91 (34) | 99 (15) | 89 (29) | 100 (16) | **69 (32)** | **99 (20)** | **81 (32)** | **100 (21)** |
| BDSS-CT | 32 (42) | 50 (36) | 83 (24) | 99 (11) | 3 (30) | 19 (40) | **83 (33)** | **99 (23)** |
| BDEI | **96 (24)** | **95 (28)** | **95 (29)** | **94 (30)** | **95 (25)** | **96 (29)** | **94 (31)** | **96 (33)** |
| BDEISS | 21 (18) | 34 (22) | **96 (29)** | **95 (33)** | 14 (18) | 27 (22) | **96 (32)** | **97 (35)** |
| BDEI-CT | 72 (27) | 59 (28) | 55 (31) | 55 (34) | **96 (25)** | **94 (31)** | **90 (35)** | **93 (39)** |
| BDEISS-CT | 40 (19) | 49 (23) | 83 (30) | 74 (35) | 14 (16) | 32 (21) | **97 (34)** | **97 (40)** |
| 500-1000-tip trees | | | | | | | | |
| BD | **66 (28)** | **99 (12)** | **72 (25)** | **99 (14)** | **38 (27)** | **98 (14)** | **56 (27)** | **98 (15)** |
| BDSS | 16 (31) | 40 (28) | **94 (16)** | **99 ( 6)** | 0 (25) | 31 (29) | **87 (21)** | **99 ( 9)** |
| BD-CT | 92 (21) | 96 ( 6) | 95 (20) | 95 ( 8) | **74 (25)** | **99 (11)** | **83 (25)** | **98 (13)** |
| BDSS-CT | 53 (36) | 47 (20) | 83 (14) | 96 ( 5) | 4 (25) | 12 (27) | **85 (21)** | **99 (12)** |
| BDEI | **95 (17)** | **94 (20)** | **94 (22)** | **95 (23)** | **96 (19)** | **95 (21)** | **94 (24)** | **95 (24)** |
| BDEISS | 13 (13) | 27 (17) | **96 (22)** | **95 (25)** | 8 (14) | 18 (18) | **96 (24)** | **96 (26)** |
| BDEI-CT | 55 (20) | 48 (21) | 42 (24) | 44 (25) | **95 (18)** | **95 (23)** | **91 (28)** | **95 (28)** |
| BDEISS-CT | 32 (15) | 38 (17) | 71 (22) | 63 (25) | 7 (12) | 21 (16) | **97 (26)** | **96 (31)** |
| 1000-2000-tip trees | | | | | | | | |
| BD | **77 (22)** | **99 ( 6)** | **70 (20)** | **99 ( 7)** | **50 (23)** | **98 ( 7)** | **50 (22)** | **98 ( 8)** |
| BDSS | 34 (30) | 34 (17) | **96 ( 9)** | **100 ( 3)** | 1 (19) | 32 (21) | **90 (15)** | **100 ( 4)** |
| BD-CT | 78 (18) | 97 ( 4) | 97 (13) | 96 ( 5) | **82 (19)** | **99 ( 6)** | **78 (20)** | **99 ( 8)** |
| BDSS-CT | 78 (31) | 43 (12) | 89 ( 7) | 92 ( 3) | 6 (20) | 11 (18) | **86 (15)** | **99 ( 7)** |
| BDEI | **96 (13)** | **97 (15)** | **95 (17)** | **96 (16)** | **95 (14)** | **96 (15)** | **95 (18)** | **96 (18)** |
| BDEISS | 12 (11) | 16 (12) | **95 (16)** | **95 (19)** | 6 (10) | 12 (14) | **96 (19)** | **96 (21)** |
| BDEI-CT | 43 (16) | 35 (15) | 33 (19) | 32 (18) | **95 (13)** | **95 (16)** | **91 (22)** | **96 (22)** |
| BDEISS-CT | 31 (13) | 26 (12) | 59 (17) | 47 (19) | 6 ( 8) | 11 (11) | **96 (21)** | **95 (26)** |
| 2000-5000-tip trees | | | | | | | | |
| BD | **79 (15)** | **99 ( 3)** | **64 (15)** | **99 ( 4)** | **54 (18)** | **99 ( 4)** | **48 (19)** | **99 ( 5)** |
| BDSS | 45 (21) | 45 ( 9) | **96 ( 6)** | **100 ( 2)** | 3 (19) | 14 (11) | **91 (10)** | **99 ( 3)** |
| BD-CT | 99 ( 5) | 93 ( 2) | 98 ( 8) | 48 ( 2) | **86 (14)** | **99 ( 3)** | **78 (16)** | **98 ( 5)** |
| BDSS-CT | 88 (15) | 72 ( 6) | 96 ( 3) | 95 ( 2) | 8 (20) | 5 ( 9) | **88 (11)** | **100 ( 5)** |
| BDEI | **95 ( 9)** | **96 (10)** | **95 (12)** | **96 (12)** | **97 (11)** | **97 (11)** | **94 (15)** | **96 (13)** |
| BDEISS | 10 ( 8) | 14 ( 9) | **95 (12)** | **96 (14)** | 6 (10) | 10 (11) | **97 (14)** | **97 (15)** |
| BDEI-CT | 24 (13) | 21 (10) | 20 (13) | 22 (12) | **97 (10)** | **97 (13)** | **94 (16)** | **96 (16)** |
| BDEISS-CT | 26 (10) | 23 ( 9) | 41 (12) | 34 (13) | 4 ( 7) | 8 ( 8) | **96 (16)** | **96 (19)** |

CI coverage (percentage of trees for which the real parameter value was within the estimated CI), and in parenthesis the corresponding mean CI width, $100(f_{E\,97.5\%} - f_{E\,2.5\%})$, are reported for 1000 trees generated under each dataset for each estimator.
The values of the estimators corresponding to or generalizing the model that generated the data are shown in bold.
The upper group of rows contains data-generating models with $f_E = 0$, the bottom group of rows contains those with $f_E \geq 0$.

Table S13: CI coverage for the superspreader fraction $f_S$ for transmission trees generated under different models (rows) and different estimators (columns). The deep-learning (DL) estimator type – pure if the estimator was trained on the corresponding model, or mixed if it was trained on the corresponding and its nested models – is specified below its model.

| | BDSS | | | | BDEISS | | | | BDSS-CT | | | | BDEISS-CT | | | |
|---|---|---|---|---|---|---|---|---|---|---|---|---|---|---|---|---|
| | DL (pure) | | DL (mixed) | | DL (pure) | | DL (mixed) | | DL (pure) | | DL (mixed) | | DL (pure) | | DL (mixed) | |
| 200-500-tip trees | | | | | | | | | | | | | | | | |
| BD | **72** | **(48)** | **99** | **(30)** | **86** | **(48)** | **100** | **(30)** | **69** | **(48)** | **99** | **(30)** | **79** | **(47)** | **99** | **(31)** |
| BDEI | 35 | (36) | 62 | (29) | **76** | **(48)** | **99** | **(30)** | 34 | (37) | 64 | (30) | **79** | **(48)** | **99** | **(31)** |
| BD-CT | 54 | (47) | 97 | (33) | 55 | (45) | 95 | (35) | **71** | **(48)** | **99** | **(31)** | **53** | **(47)** | **99** | **(33)** |
| BDEI-CT | 25 | (34) | 50 | (29) | 36 | (42) | 69 | (37) | 31 | (35) | 51 | (28) | **66** | **(47)** | **97** | **(35)** |
| BDSS | **94** | **(24)** | **95** | **(27)** | **94** | **(31)** | **96** | **(29)** | **95** | **(26)** | **96** | **(29)** | **92** | **(33)** | **96** | **(34)** |
| BDEISS | 25 | (13) | 28 | (14) | **96** | **(25)** | **95** | **(29)** | 30 | (14) | 30 | (15) | **95** | **(28)** | **96** | **(34)** |
| BDSS-CT | 92 | (22) | 93 | (25) | 89 | (27) | 93 | (26) | **96** | **(23)** | **96** | **(27)** | **94** | **(29)** | **98** | **(32)** |
| BDEISS-CT | 25 | (13) | 29 | (14) | 89 | (24) | 88 | (27) | 28 | (14) | 31 | (15) | **97** | **(28)** | **96** | **(36)** |
| 500-1000-tip trees | | | | | | | | | | | | | | | | |
| BD | **77** | **(48)** | **100** | **(23)** | **88** | **(47)** | **99** | **(25)** | **77** | **(48)** | **99** | **(24)** | **81** | **(47)** | **99** | **(25)** |
| BDEI | 33 | (32) | 52 | (22) | **76** | **(48)** | **99** | **(23)** | 35 | (32) | 49 | (23) | **80** | **(48)** | **99** | **(24)** |
| BD-CT | 52 | (46) | 96 | (27) | 53 | (43) | 91 | (30) | **81** | **(48)** | **99** | **(24)** | **66** | **(46)** | **99** | **(27)** |
| BDEI-CT | 23 | (30) | 42 | (22) | 25 | (38) | 56 | (32) | 33 | (31) | 44 | (21) | **70** | **(47)** | **97** | **(29)** |
| BDSS | **96** | **(18)** | **97** | **(19)** | **95** | **(24)** | **97** | **(21)** | **96** | **(19)** | **97** | **(20)** | **94** | **(26)** | **97** | **(23)** |
| BDEISS | 17 | ( 9) | 20 | (10) | **96** | **(19)** | **96** | **(22)** | 20 | (10) | 21 | (10) | **96** | **(22)** | **97** | **(24)** |
| BDSS-CT | 89 | (16) | 89 | (18) | 80 | (19) | 90 | (20) | **96** | **(17)** | **97** | **(19)** | **96** | **(23)** | **97** | **(22)** |
| BDEISS-CT | 17 | ( 9) | 20 | (10) | 81 | (18) | 82 | (20) | 21 | (10) | 23 | (10) | **97** | **(22)** | **97** | **(28)** |
| 1000-2000-tip trees | | | | | | | | | | | | | | | | |
| BD | **84** | **(48)** | **100** | **(18)** | **93** | **(45)** | **100** | **(21)** | **84** | **(48)** | **100** | **(20)** | **86** | **(46)** | **99** | **(22)** |
| BDEI | 31 | (28) | 47 | (18) | **80** | **(47)** | **99** | **(19)** | 34 | (29) | 42 | (17) | **78** | **(47)** | **99** | **(21)** |
| BD-CT | 58 | (46) | 95 | (22) | 50 | (40) | 90 | (26) | **83** | **(47)** | **100** | **(20)** | **74** | **(45)** | **99** | **(24)** |
| BDEI-CT | 23 | (26) | 38 | (17) | 21 | (34) | 47 | (27) | 30 | (28) | 40 | (16) | **71** | **(47)** | **98** | **(26)** |
| BDSS | **96** | **(13)** | **96** | **(14)** | **95** | **(17)** | **97** | **(14)** | **96** | **(14)** | **97** | **(15)** | **97** | **(20)** | **98** | **(16)** |
| BDEISS | 13 | ( 8) | 14 | ( 8) | **97** | **(15)** | **96** | **(17)** | 15 | ( 7) | 15 | ( 8) | **97** | **(17)** | **97** | **(18)** |
| BDSS-CT | 81 | (12) | 82 | (13) | 57 | (13) | 86 | (15) | **96** | **(13)** | **96** | **(14)** | **95** | **(17)** | **97** | **(16)** |
| BDEISS-CT | 14 | ( 8) | 14 | ( 8) | 71 | (13) | 69 | (15) | 17 | ( 8) | 18 | ( 8) | **97** | **(17)** | **97** | **(22)** |
| 2000-5000-tip trees | | | | | | | | | | | | | | | | |
| BD | **86** | **(47)** | **99** | **(14)** | **95** | **(42)** | **99** | **(18)** | **85** | **(48)** | **99** | **(17)** | **97** | **(43)** | **99** | **(18)** |
| BDEI | 26 | (24) | 46 | (13) | **83** | **(48)** | **100** | **(17)** | 30 | (26) | 37 | (14) | **86** | **(47)** | **99** | **(18)** |
| BD-CT | 50 | (44) | 92 | (21) | 49 | (36) | 52 | (29) | **84** | **(48)** | **99** | **(17)** | **84** | **(42)** | **99** | **(22)** |
| BDEI-CT | 18 | (22) | 35 | (13) | 17 | (30) | 38 | (24) | 30 | (26) | 33 | (12) | **78** | **(47)** | **98** | **(22)** |
| BDSS | **96** | **( 9)** | **97** | **(10)** | **95** | **(13)** | **96** | **(11)** | **96** | **(10)** | **97** | **(11)** | **94** | **(16)** | **97** | **(12)** |
| BDEISS | 10 | ( 5) | 14 | ( 7) | **96** | **(11)** | **97** | **(12)** | 13 | ( 7) | 14 | ( 6) | **97** | **(13)** | **98** | **(14)** |
| BDSS-CT | 68 | ( 8) | 74 | ( 9) | 52 | (11) | 78 | (11) | **97** | **( 9)** | **97** | **(10)** | **94** | **(13)** | **98** | **(12)** |
| BDEISS-CT | 9 | ( 6) | 12 | ( 7) | 59 | (10) | 56 | (10) | 14 | ( 6) | 16 | ( 7) | **95** | **(13)** | **97** | **(17)** |

CI coverage (percentage of trees for which the real parameter value was within the estimated CI), and in parenthesis the corresponding mean CI width, $100(f_{S97.5\%} - f_{S2.5\%})$, are reported for 1000 trees generated under each dataset for each estimator.
The values of the estimators corresponding to or generalizing the model that generated the data are shown in bold.
The upper group of rows contains data-generating models with $f_S = 0$, the bottom group of rows contains those with $f_S \geq 0$.

Table S14: CI coverage for the contact-tracing probability $\upsilon$ for transmission trees generated under different models (rows) and different estimators (columns). The deep-learning (DL) estimator type – pure if the estimator was trained on the corresponding model, or mixed if it was trained on the corresponding and its nested models – is specified below its model.

| | BD-CT | | | | BDEI-CT | | | | BDSS-CT | | | | BDEISS-CT | | | |
|---|---|---|---|---|---|---|---|---|---|---|---|---|---|---|---|---|
| | DL (pure) | | DL (mixed) | | DL (pure) | | DL (mixed) | | DL (pure) | | DL (mixed) | | DL (pure) | | DL (mixed) | |
| 200-500-tip trees | | | | | | | | | | | | | | | | |
| BD | **74** | **(65)** | **100** | **(25)** | **80** | **(66)** | **100** | **(29)** | **72** | **(64)** | **99** | **(28)** | **77** | **(65)** | **99** | **(30)** |
| BDEI | 80 | (59) | 100 | (15) | **76** | **(65)** | **99** | **(30)** | 79 | (60) | 100 | (18) | **76** | **(64)** | **99** | **(31)** |
| BDSS | 53 | (67) | 94 | (39) | 14 | (57) | 55 | (56) | **78** | **(66)** | **99** | **(32)** | **85** | **(64)** | **98** | **(34)** |
| BDEISS | 64 | (64) | 96 | (27) | 8 | (51) | 35 | (51) | 76 | (66) | 98 | (30) | **76** | **(67)** | **99** | **(42)** |
| BD-CT | **96** | **(34)** | **96** | **(35)** | **93** | **(36)** | **97** | **(37)** | **96** | **(36)** | **96** | **(36)** | **96** | **(41)** | **96** | **(38)** |
| BDEI-CT | 61 | (37) | 53 | (32) | **97** | **(39)** | **96** | **(41)** | 59 | (37) | 52 | (34) | **95** | **(48)** | **96** | **(48)** |
| BDSS-CT | 76 | (35) | 77 | (36) | 24 | (22) | 43 | (30) | **94** | **(37)** | **94** | **(39)** | **94** | **(43)** | **95** | **(43)** |
| BDEISS-CT | 69 | (43) | 58 | (35) | 37 | (28) | 49 | (32) | 72 | (45) | 67 | (41) | **96** | **(50)** | **96** | **(54)** |
| 500-1000-tip trees | | | | | | | | | | | | | | | | |
| BD | **86** | **(64)** | **100** | **(18)** | **86** | **(67)** | **99** | **(22)** | **86** | **(63)** | **99** | **(19)** | **85** | **(64)** | **99** | **(22)** |
| BDEI | 74 | (53) | 99 | ( 8) | **84** | **(64)** | **98** | **(23)** | 89 | (55) | 99 | (10) | **84** | **(63)** | **99** | **(25)** |
| BDSS | 46 | (67) | 89 | (34) | 6 | (54) | 36 | (56) | **89** | **(64)** | **99** | **(23)** | **89** | **(62)** | **99** | **(25)** |
| BDEISS | 51 | (61) | 92 | (22) | 5 | (46) | 20 | (50) | 78 | (65) | 99 | (19) | **79** | **(64)** | **98** | **(35)** |
| BD-CT | **95** | **(26)** | **96** | **(27)** | **92** | **(29)** | **96** | **(28)** | **96** | **(28)** | **96** | **(28)** | **96** | **(33)** | **96** | **(29)** |
| BDEI-CT | 46 | (25) | 42 | (24) | **96** | **(30)** | **96** | **(32)** | 46 | (27) | 41 | (24) | **94** | **(39)** | **96** | **(38)** |
| BDSS-CT | 61 | (28) | 66 | (32) | 12 | (17) | 18 | (24) | **95** | **(29)** | **96** | **(32)** | **96** | **(36)** | **96** | **(34)** |
| BDEISS-CT | 56 | (32) | 50 | (30) | 19 | (20) | 28 | (27) | 59 | (35) | 54 | (32) | **96** | **(42)** | **96** | **(47)** |
| 1000-2000-tip trees | | | | | | | | | | | | | | | | |
| BD | **89** | **(63)** | **100** | **(14)** | **91** | **(67)** | **100** | **(16)** | **86** | **(62)** | **100** | **(15)** | **88** | **(61)** | **100** | **(19)** |
| BDEI | 76 | (49) | 99 | ( 5) | **89** | **(63)** | **98** | **(18)** | 90 | (54) | 100 | ( 6) | **88** | **(62)** | **99** | **(21)** |
| BDSS | 32 | (66) | 81 | (33) | 2 | (53) | 24 | (54) | **89** | **(65)** | **99** | **(18)** | **94** | **(59)** | **99** | **(21)** |
| BDEISS | 41 | (58) | 88 | (20) | 2 | (44) | 10 | (50) | 76 | (65) | 99 | (13) | **83** | **(63)** | **98** | **(30)** |
| BD-CT | **96** | **(20)** | **96** | **(21)** | **94** | **(24)** | **97** | **(22)** | **96** | **(22)** | **97** | **(22)** | **94** | **(25)** | **97** | **(23)** |
| BDEI-CT | 36 | (17) | 32 | (17) | **96** | **(22)** | **97** | **(25)** | 36 | (18) | 32 | (19) | **95** | **(30)** | **97** | **(30)** |
| BDSS-CT | 50 | (23) | 48 | (25) | 7 | (13) | 9 | (23) | **95** | **(22)** | **97** | **(25)** | **96** | **(27)** | **96** | **(27)** |
| BDEISS-CT | 45 | (23) | 43 | (23) | 7 | (15) | 13 | (22) | 47 | (25) | 43 | (26) | **96** | **(34)** | **95** | **(39)** |
| 2000-5000-tip trees | | | | | | | | | | | | | | | | |
| BD | **90** | **(63)** | **99** | **(12)** | **88** | **(69)** | **100** | **(14)** | **91** | **(62)** | **100** | **(10)** | **90** | **(62)** | **100** | **(16)** |
| BDEI | 55 | (49) | 98 | ( 4) | **91** | **(64)** | **99** | **(15)** | 88 | (49) | 97 | ( 4) | **92** | **(62)** | **99** | **(17)** |
| BDSS | 18 | (72) | 76 | (30) | 0 | (51) | 9 | (48) | **93** | **(64)** | **99** | **(12)** | **95** | **(58)** | **99** | **(17)** |
| BDEISS | 33 | (63) | 86 | (16) | 0 | (44) | 4 | (48) | 74 | (64) | 99 | ( 8) | **89** | **(61)** | **99** | **(25)** |
| BD-CT | **96** | **(14)** | **96** | **(15)** | **94** | **(18)** | **96** | **(16)** | **97** | **(16)** | **96** | **(16)** | **92** | **(21)** | **98** | **(18)** |
| BDEI-CT | 29 | (13) | 27 | (12) | **95** | **(16)** | **96** | **(19)** | 30 | (13) | 27 | (14) | **96** | **(23)** | **97** | **(23)** |
| BDSS-CT | 34 | (19) | 46 | (22) | 2 | (12) | 4 | (17) | **95** | **(16)** | **96** | **(19)** | **95** | **(22)** | **96** | **(21)** |
| BDEISS-CT | 37 | (20) | 33 | (18) | 4 | (12) | 8 | (19) | 39 | (19) | 36 | (20) | **96** | **(26)** | **95** | **(32)** |

CI coverage (percentage of trees for which the real parameter value was within the estimated CI), and in parenthesis the corresponding mean CI width, $100(\upsilon_{97.5\%} - \upsilon_{2.5\%})$, are reported for 1000 trees generated under each dataset for each estimator.
The values of the estimators corresponding to or generalizing the model that generated the data are shown in bold.
The upper group of rows contains data-generating models with $\upsilon = 0$, the bottom group of rows contains those with $\upsilon \geq 0$.

Table S15: CI coverage for the superspreading transmission increase $X_S$ for transmission trees generated under different models (rows) and different estimators (columns). The deep-learning (DL) estimator type – pure if the estimator was trained on the corresponding model, or mixed if it was trained on the corresponding and its nested models – is specified below its model.

| | BDSS | | BDEISS | | BDSS-CT | | BDEISS-CT | |
|---|---|---|---|---|---|---|---|---|
| | DL (pure) | DL (mixed) | DL (pure) | DL (mixed) | DL (pure) | DL (mixed) | DL (pure) | DL (mixed) |
| 200-500-tip trees | | | | | | | | |
| BDSS | **96 (125)** | **95 (130)** | **94 (131)** | **95 (132)** | **95 (128)** | **95 (131)** | **94 (137)** | **95 (134)** |
| BDEISS | 60 (90) | 65 (99) | **96 (120)** | **96 (128)** | 61 (92) | 66 (101) | **96 (129)** | **96 (135)** |
| BDSS-CT | 95 (116) | 96 (124) | 95 (126) | 96 (128) | **96 (119)** | **97 (128)** | **97 (131)** | **97 (136)** |
| BDEISS-CT | 59 (90) | 64 (100) | 90 (118) | 89 (125) | 59 (92) | 65 (102) | **97 (134)** | **96 (144)** |
| 500-1000-tip trees | | | | | | | | |
| BDSS | **96 (104)** | **96 (109)** | **93 (107)** | **95 (114)** | **95 (108)** | **96 (114)** | **93 (116)** | **95 (116)** |
| BDEISS | 50 (72) | 53 (76) | **96 (97)** | **95 (108)** | 51 (74) | 58 (86) | **95 (107)** | **94 (116)** |
| BDSS-CT | 94 (92) | 94 (98) | 91 (95) | 95 (107) | **97 (97)** | **96 (107)** | **95 (107)** | **97 (114)** |
| BDEISS-CT | 48 (71) | 52 (77) | 84 (94) | 85 (104) | 49 (73) | 54 (87) | **97 (108)** | **96 (126)** |
| 1000-2000-tip trees | | | | | | | | |
| BDSS | **96 (87)** | **96 (93)** | **92 (92)** | **96 (97)** | **96 (93)** | **96 (96)** | **92 (100)** | **94 (99)** |
| BDEISS | 42 (58) | 47 (72) | **96 (81)** | **96 (91)** | 42 (61) | 48 (73) | **95 (89)** | **96 (99)** |
| BDSS-CT | 90 (71) | 92 (79) | 87 (80) | 93 (90) | **97 (79)** | **97 (88)** | **95 (90)** | **97 (95)** |
| BDEISS-CT | 38 (54) | 44 (69) | 79 (78) | 78 (85) | 41 (56) | 46 (74) | **96 (89)** | **96 (109)** |
| 2000-5000-tip trees | | | | | | | | |
| BDSS | **95 (65)** | **95 (71)** | **90 (72)** | **95 (75)** | **95 (74)** | **95 (74)** | **90 (84)** | **95 (79)** |
| BDEISS | 31 (41) | 33 (54) | **96 (63)** | **96 (72)** | 35 (51) | 39 (62) | **95 (73)** | **96 (78)** |
| BDSS-CT | 78 (49) | 84 (56) | 70 (58) | 80 (63) | **96 (61)** | **97 (68)** | **93 (72)** | **97 (73)** |
| BDEISS-CT | 28 (36) | 34 (52) | 69 (59) | 67 (62) | 34 (46) | 39 (62) | **96 (71)** | **96 (86)** |

CI coverage (percentage of trees for which the real parameter value was within the estimated CI), and in parenthesis the corresponding mean relative CI width, $100(X_{S97.5\%} - X_{S2.5\%})/X_{S\,true}$, are reported for 1000 trees generated under each dataset for each estimator.
The values of the estimators corresponding to or generalizing the model that generated the data are shown in bold.

Table S16: CI coverage for the contact-traced removal speed up $X_C$ for transmission trees generated under different models (rows) and different estimators (columns). The deep-learning (DL) estimator type – pure if the estimator was trained on the corresponding model, or mixed if it was trained on the corresponding and its nested models – is specified below its model.

| | BD-CT | | BDEI-CT | | BDSS-CT | | BDEISS-CT | |
|---|---|---|---|---|---|---|---|---|
| | DL (pure) | DL (mixed) | DL (pure) | DL (mixed) | DL (pure) | DL (mixed) | DL (pure) | DL (mixed) |
| 200-500-tip trees | | | | | | | | |
| BD-CT | **95 (143)** | **95 (144)** | **95 (168)** | **96 (155)** | **96 (151)** | **96 (150)** | **97 (177)** | **96 (162)** |
| BDEI-CT | 94 (176) | 93 (175) | **95 (181)** | **94 (178)** | 95 (181) | 91 (176) | **96 (191)** | **95 (183)** |
| BDSS-CT | 91 (142) | 89 (142) | 88 (181) | 85 (167) | **96 (159)** | **94 (166)** | **94 (193)** | **95 (179)** |
| BDEISS-CT | 90 (186) | 86 (165) | 92 (182) | 89 (175) | 92 (196) | 87 (173) | **96 (196)** | **95 (190)** |
| 500-1000-tip trees | | | | | | | | |
| BD-CT | **96 (110)** | **95 (114)** | **94 (125)** | **96 (123)** | **97 (119)** | **96 (120)** | **96 (153)** | **96 (131)** |
| BDEI-CT | 89 (150) | 90 (158) | **95 (140)** | **96 (149)** | 90 (155) | 87 (158) | **96 (166)** | **95 (156)** |
| BDSS-CT | 85 (105) | 88 (112) | 79 (150) | 71 (141) | **96 (121)** | **95 (129)** | **95 (165)** | **95 (146)** |
| BDEISS-CT | 87 (161) | 85 (155) | 85 (147) | 86 (157) | 90 (174) | 88 (160) | **96 (173)** | **96 (172)** |
| 1000-2000-tip trees | | | | | | | | |
| BD-CT | **96 (87)** | **95 (90)** | **93 (101)** | **96 (97)** | **96 (93)** | **95 (96)** | **95 (114)** | **95 (108)** |
| BDEI-CT | 76 (134) | 80 (143) | **96 (113)** | **95 (126)** | 76 (131) | 78 (151) | **96 (127)** | **95 (135)** |
| BDSS-CT | 79 (83) | 80 (87) | 70 (139) | 62 (118) | **96 (93)** | **97 (104)** | **95 (120)** | **96 (116)** |
| BDEISS-CT | 76 (144) | 79 (143) | 79 (126) | 84 (135) | 85 (156) | 84 (149) | **97 (137)** | **96 (150)** |
| 2000-5000-tip trees | | | | | | | | |
| BD-CT | **97 (61)** | **96 (66)** | **95 (74)** | **97 (71)** | **96 (66)** | **96 (69)** | **96 (88)** | **96 (77)** |
| BDEI-CT | 61 (121) | 63 (130) | **95 (86)** | **96 (98)** | 59 (129) | 59 (135) | **96 (98)** | **96 (104)** |
| BDSS-CT | 72 (66) | 77 (69) | 61 (128) | 62 (111) | **96 (67)** | **95 (80)** | **97 (95)** | **97 (89)** |
| BDEISS-CT | 59 (139) | 63 (134) | 75 (104) | 79 (117) | 65 (152) | 66 (144) | **97 (108)** | **97 (124)** |

CI coverage (percentage of trees for which the real parameter value was within the estimated CI), and in parenthesis the corresponding mean relative CI width, $100(X_{C97.5\%} - X_{C2.5\%})/X_{C\,true}$, are reported for 1000 trees generated under each dataset for each estimator.
The values of the estimators corresponding to or generalizing the model that generated the data are shown in bold.

Table S17: Hong Kong SARS-CoV-2 wave 3 epidemiological parameters and their 95% CIs (columns) estimated with different models (rows).

| | $R_e$ | $d$ [days] | $f_E$ | $f_S$ | $X_S$ | $\upsilon$ | $X_C$ |
|---|---|---|---|---|---|---|---|
| BD (ML) | 1.38 (1.12 - 1.70) | 4.20 (3.71 - 4.79) | | | | | |
| BD | 1.55 (1.31 - 1.78) | 4.86 (4.14 - 5.99) | | | | | |
| BDEI | 1.37 (1.26 - 2.14) | 5.35 (4.25 - 8.64) | 0.95 (0.82 - 0.98) | | | | |
| BDSS | 1.50 (1.17 - 2.00) | 5.45 (4.10 - 6.35) | | 0.04 (0.00 - 0.15) | 14.12 (9.69 - 21.01) | | |
| BD-CT | 1.43 (1.26 - 1.75) | 4.42 (3.44 - 5.74) | | | | 0.15 (0.02 - 0.32) | 54.05 (11.96 - 112.59) |
| BDEISS | 1.70 (1.33 - 2.04) | 5.40 (3.55 - 8.61) | 0.94 (0.71 - 1.00) | 0.27 (0.01 - 0.50) | 6.32 (1.66 - 18.22) | | |
| BDEI-CT | 1.73 (1.51 - 2.59) | 7.50 (5.73 - 11.85) | 0.95 (0.85 - 0.98) | | | 0.66 (0.47 - 0.75) | 87.61 (39.99 - 106.86) |
| BDSS-CT | 1.59 (1.16 - 2.16) | 4.91 (3.57 - 7.00) | | 0.05 (0.00 - 0.12) | 17.94 (12.81 - 22.17) | 0.26 (0.05 - 0.58) | 48.35 (14.12 - 106.09) |
| BDEISS-CT | 1.91 (1.54 - 2.75) | 7.55 (5.46 - 10.70) | 0.92 (0.74 - 0.95) | 0.00 (0.00 - 0.30) | 1.16 (1.00 - 6.89) | 0.67 (0.38 - 0.77) | 80.11 (36.80 - 100.39) |
| BDSS (Xie *et al.* [13]) | 1.59 (1.33 - 1.99) | 4.64 (3.37 - 8.24) | | 0.09 (0.05 - 0.17) | 8.08 (3.91 - 17.73) | | |
| Epi (Xie *et al.* [13]) | 1.69 (1.65 - 1.74) | | | | | | |

BD (ML) is a maximum-likelihood estimator for the BD model [14].
The second group of estimators (below BD (ML) and above those from Xie *et al.* [13]) are the DL-based estimators described in this study.
BDSS (Xie *et al.* [13]) is a DL-based estimator inference described in Xie *et al.* [13].
Epi (Xie *et al.* [13]) is an epidemiological inference using a combination of line-listed incidence data to estimate $R$ described in Xie *et al.* [13].

[13] Ruopeng Xie, Dillon C Adam, Shu Hu, Benjamin J Cowling, Olivier Gascuel, Anna Zhukova, and Vijaykrishna Dhanasekaran. Integrating contact tracing data to enhance outbreak phylodynamic inference: a deep learning approach. *Molecular Biology and Evolution*, page msae232, November 2024.

[14] Anna Zhukova and Olivier Gascuel. Accounting for contact tracing in epidemiological birth-death models. *PLOS Computational Biology*, 21(5):e1012461, May 2025.
